# Assessing the nationwide impact of COVID-19 mitigation policies on the transmission rate of SARS-CoV-2 in Brazil

**DOI:** 10.1101/2020.06.26.20140780

**Authors:** Daniel C. P. Jorge, Moreno S. Rodrigues, Mateus S. Silva, Luciana L. Cardim, Nívea B. da Silva, Ismael H. Silveira, Vivian A. F. Silva, Felipe A. C. Pereira, Arthur R. de Azevedo, Alan A. S. Amad, Suani T.R. Pinho, Roberto F. S. Andrade, Pablo I. P. Ramos, Juliane F. Oliveira

## Abstract

COVID-19 is now identified in almost all countries in the world, with poorer regions being particularly more disadvantaged to efficiently mitigate the impacts of the pandemic. In the absence of efficient therapeutics or vaccines, control strategies are currently based on non-pharmaceutical interventions, comprising changes in population behavior and governmental interventions, among which the prohibition of mass gatherings, closure of non-essential establishments, quarantine and movement restrictions. In this work we analyzed the effects of 707 published governmental interventions, and population adherence thereof, on the dynamics of COVID-19 cases across all 27 Brazilian states, with emphasis on state capitals and remaining inland cities. A generalized SEIR (Susceptible, Exposed, Infected and Removed) model with a time-varying transmission rate (TR), that considers transmission by asymptomatic individuals, is presented. We analyze the effect of both the extent of enforced measures across Brazilian states and population movement on the changes in the TR and effective reproduction number. The social mobility reduction index, a measure of population movement, together with the stringency index, adapted to incorporate the degree of restrictions imposed by governmental regulations, were used in conjunction to quantify and compare the effects of varying degrees of policy strictness across Brazilian states. Our results show that population adherence to social distance recommendations plays an important role for the effectiveness of interventions and represents a major challenge to the control of COVID-19 in low- and middle-income countries.

## 1. Introduction

COVID-19, a disease caused by the SARS-CoV-2 coronavirus, emerged in December 2019 in China and was recognized as a pandemic by the World Health Organization on March 11, 2020 [1]. At that moment, Brazil had already confirmed 53 cases. On March 20, with 972 confirmed cases, the Brazilian Ministry of Health recognized community transmission of COVID-19 throughout the national territory, 24 days after the first confirmed case of COVID-19 was identified [2]. Brazil is a country with 209.5 million individuals and stark socioeconomic disparities throughout its territory. It is the largest country in South America and the fifth largest nation in the world. Accordingly, the many challenges imposed by the COVID-19 pandemic are unprecedented in this country.

The political-administrative organization of Brazil comprises three spheres of governance: The Union (federal government), the 27 states (including the Federal District, where the capital city, Brasilia, is located) and 5,570 municipalities. To reduce the transmission of SARS-CoV-2, federal, state and city governments implemented a series of interventions by means of government decrees [3]. This included recommendations to identify and isolate confirmed cases and contacts; to restrict unnecessary movements; to practice social distancing; to increase hygiene awareness; to follow respiratory etiquette; to wear masks in public, among others. In the absence of more intensive mitigation policies implemented by the federal government (such as lock-downs and movement restrictions), most measures were adopted by local governments (state/municipalities) [3]. However, adherence to these policies varied greatly throughout the country, and while some regions enacted more strict controls, others have been more lax.

The COVID-19 crisis has called attention to the importance of mathematical modeling to inform governmental policies and in quantifying the effects of the pandemic at multiple levels [4]. Models have been adapted to address questions such as the role of asymptomatic individuals to SARS-CoV-2 transmission chains [5], to assess the effectiveness of testing and quarantine strategies in informing economic reopening [6] and epidemic suppression by partial lockdown strategies [7], to quantify the needs for hospital beds under various social distancing scenarios [8, 9, 10], among others, eg. [11, 12, 13, 14]. By drawing on a generalized SEIR model that simulates the dynamics of viral spread in a population entirely susceptible to the new virus, it is possible to define and estimate the transmission rate (TR) at which an infected individual will transmit the disease to a susceptible person [15]. Therefore, the higher this rate, the greater the number of new cases in a region over time. Downward changes on the TR are expected with the implementation of mitigation policies such as non-pharmaceutical interventions (NPI), the only option to limit the spread of SARS-CoV-2 in the absence of effective therapies or vaccination with sufficient population coverage.

In this work, we comparatively analyze the evolution of the COVID-19 transmission rate and effective reproduction number across all 27 Brazilian states, with emphasis on state capitals and remaining inland cities, establishing links with measures of governmental restrictions (NPIs) implemented in each region together with the human behavior response, particularly the adherence to recommendations of social distancing. The varying degree of enforced policies across the country offers an opportunity to study the impacts of interventions, including their breadth and timing, on the TR of SARS-CoV-2 throughout Brazilian states. These findings can be extrapolated to similar settings in other low- and middle-income countries to drive improvements in mitigation policies against subsequent waves of SARS-CoV-2 and other potentially pandemic pathogens.

## 2. Methodology

### Data sources

The number of confirmed cases of COVID-19 for each Brazilian municipality, up to May 22, 2020, was obtained from the Ministry of Health of Brazil and are publicly available [16, 17]. Since the capitals of each state have different dynamics and largely concentrated COVID-19 cases in the initial wave of the epidemic (an assumption informed by previous works [18, 19]), we analyzed the transmission dynamics for each state in the following way: 1) The entire state, 2) the state capital alone; and 3) the remaining state municipalities grouped as a single place by summing up their reported cases. Throughout the text we refer to the latter as inland cities, although strictly not all of these are distant from the shore.

To evaluate state-wide enforced governmental measures, we collated government decrees and resolutions scattered throughout various state government gazettes and other official repositories up to May 22, 2020, since each state uses different platforms to communicate their legislation. We annotated the type of measure enforced, the implementation date, the duration and whether it was valid to the whole state or limited to specific regions (see Supplementary Material).

In addition to state level decrees we also collated COVID-19-related federal regulations, which affect equally all states [20]. However, federal-level regulations did not interfere on measures enforced by state or municipalities, and most of them were later repealed. Indeed, on April 8, 2020 the Brazilian Federal Supreme Court ruled in favor of the decentralization of operations aimed at controlling COVID-19 in the country, empowering the actions of state and municipal governments [21]. The evaluation of municipal-level regulations was not performed due to the large number of Brazilian cities, a total of 5,570, each having their own system for publications of legislation, with variable degrees of update frequency and usually providing unstructured data and burdensome access options, which would make their manual processing unpractical. For this reason, in this work we only considered regulations issued by the 27 state governments, which totaled 707 decrees.

As a proxy of the population adherence to recommendations of social distancing, we used information from InLoco [22], a Brazilian technology start-up that developed an index of social mobility, which seeks to help in fight the pandemic in Brazil. Data for the index construction is obtained from the unidentified, aggregated geo-movement patterns extracted from 60 million mobile devices throughout the country. The index ranges from 0 to 100% and measures the proportion of devices from a given municipality that remained within a 450 meter radius from the location identified as home by the device. The higher the index, the greatest the population adherence to social distancing recommendations. The data is available at [22]. Examples of other works that used the Social mobility reduction index (SMRI) can be found in [23, 24].

### Stringency Index

To comparatively evaluate the governmental measures implemented by the Brazilian states, we constructed a stringency index, denoted by *ι*, similarly to that implemented in [25]. To score each employed policy we adapted the methodology to the Brazilian context by taking into account the specific measures established by the different state governments and described in the Data Sources section.

Measures were classified into two categories: Ordinal and cumulative. Ordinal class of measures, denoted by *O*, are those in which there is a clear order on the intensity of the restriction, so that there are less possibilities of reclassification. For instance, a decree prohibiting agglomerations of more than 100 people, followed by a second decree restricting to 500 people, belong to the ordinal category, where the first is more intense than the second. Cumulative class of measures, denoted by *C*, are those with no clear order of intensity, allowing for a wide range of possibilities to classify the restriction. For instance, closure of malls and prohibition to accessing parks and beaches have no clear order to which of these measures is more intense and may lead to subjective classification.

Every class of measures is a collection of sub-measures depending on how each government decided to response and enforce measures. In this work we have seven classes of measures, of which four are ordered and three are cumulative, further described in Supplementary Table 1. Cancellation of public events (*O*_1_) presents six sub-measures. Similarly, health etiquette policies (*C*_1_) has two sub-measures.

The evaluation of the ordered class of measures varies from 0 (when no measure is applied) to *j* (when the most stringent sub-measure is applied), where the index *j* varies from 0 to 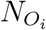, the number of sub-measures of the ordinal class of measures *O*_*i*_ (we similarly denote 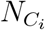, as the number of sub-measures for the cumulative class). For the cumulative class of measures, we evaluate each sub-measure as 0 if the sub-measure was not applied or 1 otherwise, so that the value of the class will be the sum of points of the sub-measures 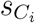. Additionally, to take into account whether the measure was enforced for the whole state or limited to a particular region, each class has a target *G*. If the measure is ordinal, then we consider 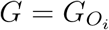, taking either the value 0, if the most stringent sub-measure is applied for specific areas of the state, or 1, if it is enforced for the whole state. If the measure is cumulative, and since it will be a sum of the sub-measures, then 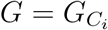 is the sum of targets for each sub-measure 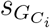, which again is either 0 if the sub-measure is applied to specific areas or 1 if it is applied to the whole state without exceptions. A summary of the seven measures, as well as their number of sub-measures and targets are presented in Table 1.

**Table 1:**
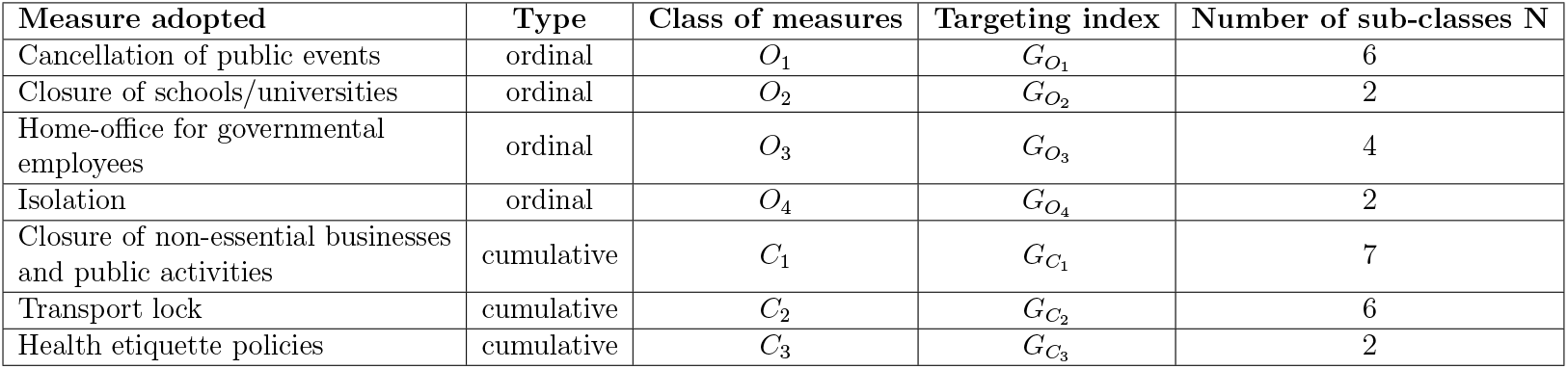
Classification of governmental responses to COVID-19 in Brazil (state-wide).

| Measure adopted | Type | Class of measures | Targeting index | Number of sub-classes N |
| --- | --- | --- | --- | --- |
| Cancellation of public events | ordinal | $O_1$ | $G_{O_1}$ | 6 |
| Closure of schools/universities | ordinal | $O_2$ | $G_{O_2}$ | 2 |
| Home-office for governmental employees | ordinal | $O_3$ | $G_{O_3}$ | 4 |
| Isolation | ordinal | $O_4$ | $G_{O_4}$ | 2 |
| Closure of non-essential businesses and public activities | cumulative | $C_1$ | $G_{C_1}$ | 7 |
| Transport lock | cumulative | $C_2$ | $G_{C_2}$ | 6 |
| Health etiquette policies | cumulative | $C_3$ | $G_{C_3}$ | 2 |

The index for the ordinal classes is given by

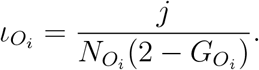

Thus, the value of the stringency 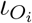 is the value of the most stringent measured applied, *j*, normalized by the number of sub-classes, 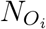, multiplied by the term 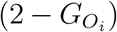, which is 1 when the target is for the whole state or 2 otherwise (that is, the stringency 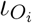 is divided by two).

For the cumulative classes the index is defined by

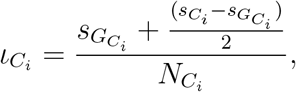

where the sum 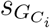 reflects the measures that were applied to the whole state, and the term 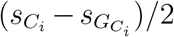 evaluates the measures that were not applied for the whole state. Lastly, the whole expression is normalized by the number of sub-classes 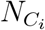.

Therefore, the total state index *ι*, for a given day, will be taken as the average of the value of the classes 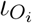 and 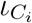, yielding

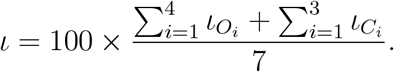

Examples of these estimates are given in Supplementary Note 1.

### The mathematical model

We generalize the usual SEIR model by separating the infectious compartment into two: Asymptomatic/non-detected and symptomatic cases, denoted by *I*_*a*_ and *I*_*s*_, respectively, following our previous modeling strategy [8]. It is reasonable to consider that the compartment *I*_*a*_ encompasses many of the sub-notified cases since the majority of asymptomatic and mild cases are not accounted for in the official data, in spite of their important role to the SARS-CoV-2 transmission chain [5]. The system of differential equations reads:

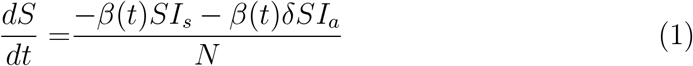

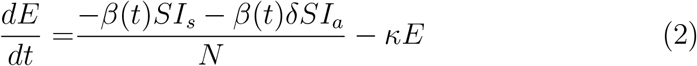

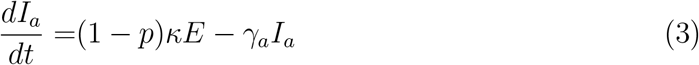

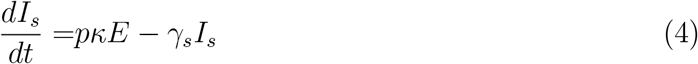

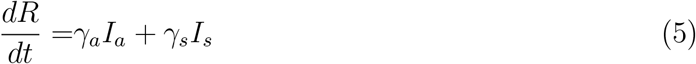

The epidemiological parameters presented in Equations (1 -5) are described in Table 2, along with their estimates (or search ranges, when fitted), which were informed by previous works.

**Table 2:** Key epidemiological parameters used in the SEIR model, with their respective value (when fixed) or the search intervals used for parameter estimations, informed by the literature.

| Parameter | Description | Search interval | Fixed | Reference |
| --- | --- | --- | --- | --- |
| $\beta$ | Transmission rate | [0,2] | - | [5, 26, 27, 28] |
| $t_i$ | Time of when transmission rate change | Defined in the Result section | - | [27, 28] |
| $\delta$ | Asymptomatic infectivity | [0,0.7] | - | [5, 29, 30, 9, 31, 32] |
| $p$ | Proportion of latent (E) that proceed to $I_s$ | - | 0.2 | [5, 31, 33] |
| $\kappa$ | Mean exposed period, or incubation time | - | 1/4 | [5, 30, 34, 35, 36] |
| $\gamma_a$ | Mean asymptomatic period | - | 1/3.5 | [5] |
| $\gamma_s$ | Mean symptomatic period | - | 1/4 | [5, 26, 37] |

As described in [8], the parameter *δ*, a factor associated with the infectivity of asymptomatic/non-detected infections, plays an important role on the viral transmission dynamics. As described by equation 1, it influences the non-linear term, and thus cannot be neglected from the system, unless it is assumed that the infectivity of asymptomatic individuals is equivalent to that of symptomatic cases, ie. *δ* = 1. However, several studies suggest *δ <* 1 [5, 29, 32], which guided our modeling choice that defined the search interval for this parameter (Table 2).

Additionally, to account for variations in the TR over time, we assume that the TR *β* is a function given by:

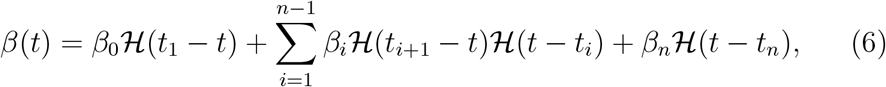

where {*t*_1_, *t*_2_, …, *t*_*n*_} represent a set of points in time defining the change on the TR, *β*_*i*_ are TRs in the interval [*t*_*i*_, *t*_*i*+1_], and ℋ(*x*) = 1 if *x >* 0, ℋ(*x*) = 0 if *x <* 0 and ℋ(*x*) = 1*/*2 if *x* = 0.

The key novelty here is that the influence of governmental measures on social mobility and human behavior can dictate changes on the TR (*β*_*i*_’s) on inferred points *t*_*i*_’s. To do so, we assume that the *t*_*i*_’s change in an interval search based on variations in stringency and social mobility, described previously. Additionally, to evaluate if one or more changes to the transmission rate was required, we assessed the value obtained using the Bayesian information criterion (BIC) as well as by visual evaluation of the fitted results. The visual evaluation is important since the calculation of the BIC may yield small variations when the number of parameters is increased due to the inherent noise in the data.

The time of changing transmission rate and the rate itself, as well as the *δ* parameter are estimated using the Particle Swarm Optimization (PSO) metaheuristic. Under the PSO framework, we used maximum likelihood estimation to optimize the model to the series of daily confirmed cases for each state, capital cities and remaining inland cities up to May 22, 2020. PSO was implemented using `pyswarms` library version 1.1.0 for Python 3 (http://python.org) [38], and was executed with 150 particles through 500 iterations with cognitive parameter 0.1, social parameter 0.3, inertia parameter 0.9, evaluating five closest neighbors through Euclidean (or L2) distance metric. In addition to the point estimates obtained by the PSO method, percentile confidence intervals were also estimated for these parameters. The intervals were constructed using the weighted non-parametric bootstrap method, considering 100 replicates of the original series of new cases. Similar work to evaluate the impact of non-pharmaceutical measures on the TR and dynamic spread of the disease are presented in [27, 28, 39]. The parameters *p, κ, γ*_*a*_, *γ*_*s*_ were informed by the literature and kept fixed. The remaining estimated model parameters and search intervals were informed both by the literature and the analyses presented in this work, and are presented in Table 2.

### Basic and effective reproduction number

Based on the obtained parameter values we also evaluated the basic reproductive number *ℛ*_0_ and the effective reproductive number *ℛ*(*t*), also denoted by *ℛ*_*t*_, as described in [8]. The basic reproductive number *ℛ*_0_ characterizes the disease spread during the very early time interval after it has been introduced, thus it is evaluated on the first transmission rate estimated by the model. Its expression is given by (see [8, 40, 41]):

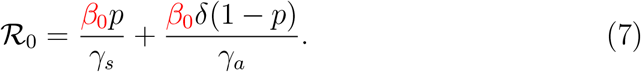

We also obtain the time series of the effective reproductive number *ℛ* (*t*), which indicate the current trend of the epidemic. Details of this calculation can be found in Supplementary Note 5 of our previous work [8]. By writing the infection-age structured epidemic model of our system of equations (1) to (5) [40, 42], we obtain:

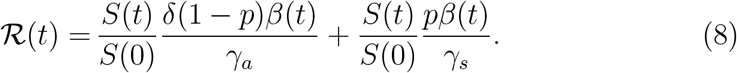

Equation (8) would be similar to the general form given by *ℛ* (*t*) = *ℛ*_0_*S*(*t*) only for a portion of the evaluated period, since we consider changes on time in the transmission rate *β* and the *ℛ*_0_ expression only takes into account the transmission rate before the first change, that is, for *t < t*_1_.

Additionally, equation (8) can be also given by:

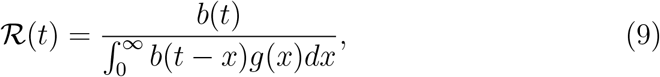

where the numerator *b* represents daily number of new cases and the denominator corresponds to the convolution on *b* and *g*, that is the disease probability distribution function for the time interval between the infection of an individual and its secondary cases. The function *g* is derived from the differential equations of the model and receives contributions from the three compartments *E, I*_*a*_, *I*_*s*_ that impact the evaluation of *ℛ*_0_ and *ℛ* (*t*), as presented in [8]. The advantage of equation (9) is that it allow us to evaluate the effective reproduction number directly on the available data and *g*, a probability distribution function that can take different shapes and makes our result more flexible to compare directly with the data, being *δ* the only estimated necessary parameter used to feed equation (9).

Finally, since the data are reported as daily case counts, to calculate expression (9), we discretize the generation time distribution *g*, such that 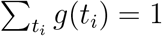, see [42]. Thus,

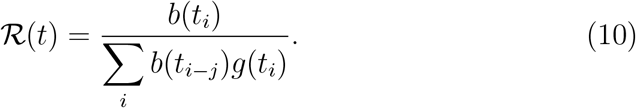

### Visualization app

To facilitate visualizing the results of the analysis performed in this work, a web-based application was developed using R/Shiny (http://shiny.rstudio.com). The application can be found at https://modelingtaskforce.shinyapps.io/transmission_rate_app/.

### Data availability

Codes used to produce the results presented herein and related datasets are available as supplementary material and in a public GitHub repository [43]. Case series data, stringency and SMRI series, model fits and details of estimates can be found in the on-line web page app at https://modelingtaskforce.shinyapps.io/transmission_rate_app/.

### Ethics statement

This study was conducted with publicly available data from the COVID-19 epidemic, published by the Ministry of Health of Brazil or third parties. Therefore, no approval by an ethics committee was required, according to Resolutions 466/2012 and 510/2016 (article 1, sections III and V) from the National Health Council (CNS), Brazil.

## 3. Results

### The entry of SARS-CoV-2 in the Brazilian states

Brazil had a total of 334,555 registered COVID-19 cases up to May 22, 2020. The first confirmed case of COVID-19 in Brazil occurred in the state of São Paulo, southeastern region, on February 26, 2020. Within an interval of seven to 12 days (from March 4 to March 9) the states^10^ of ES, DF, BA, AL, RJ and MG had confirmed cases of COVID-19. From March 10 to March 25 the disease spread to the remaining 20 Brazilian states (Figure 1A).

**Figure 1:**
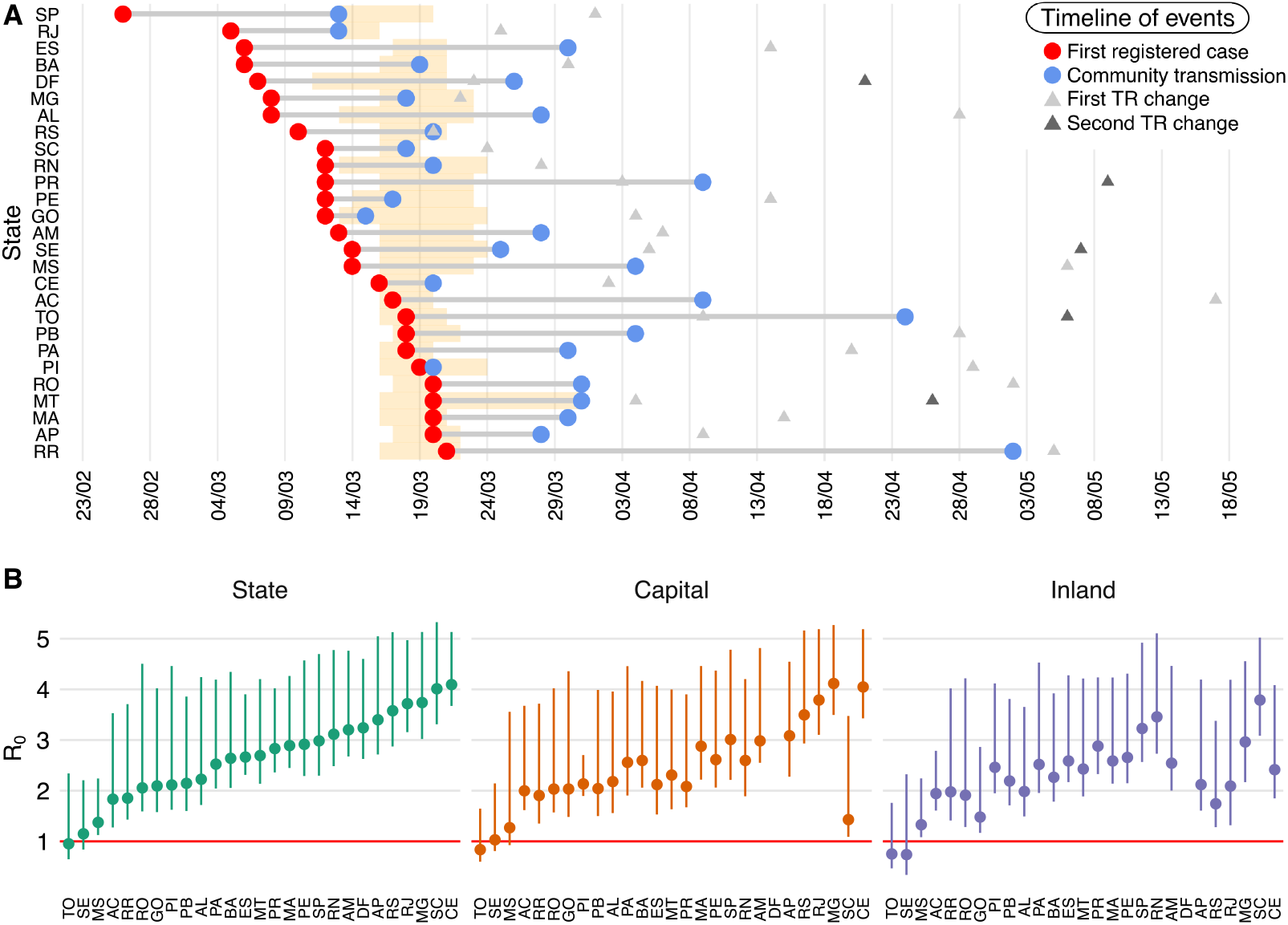
A) A timeline of events associated with the spread of SARS-CoV-2 across the 27 Brazilian states. The dates of first registered cases are shown (red dots), followed by the moment that community transmission was declared in each state (blue dots). The dates when the TR were observed to change (*β*_0_ →*β*_1_ and, when applicable, *β*_1_ →*β*_2_, are shown as triangles). The orange segments indicate the interval between the initial NPIs enforced by states and the first observed peak in stringency. Dates refer to the year 2020. B) Estimates of *R*_0_ (and corresponding 95% confidence intervals) for each state, their capitals and remaining inland cities. Data for the Federal District (DF), the smallest Brazilian federal unit and the only one that has no municipalities, is only shown at the state-level.

By the moment that state authorities were unable to relate confirmed cases through chains of transmission, SARS-CoV-2 community transmission began to be declared in each region (Figure 1A; Supplementary Table 2). Supplementary Figure 1 shows the temporal variation from the first reported case to the declaration of community transmission in the state. As depicted in Supplementary Figure 1, 40.7% (11/27) of the states took between 0 and 10 days to declare community transmission, 51.9% (14/27) took between 11 and 30 days, while only two states (Tocantins and Roraima), corresponding to 7.5%, took more than 30 days to declare community transmission. This scenario is further complicated by the delays in reporting cases. Although each state declared community transmission at some point, there are uncertainties around these dates due to delays in both the registration of cases and the communication from the local level (municipality) to state authorities, as exemplified in the states of Paraná and Roraima [44, 45].

In the supplemental web-page (see Data availability section), we present the incidence of COVID-19 for each state, capitals and inland cities. As shown, in the early stage of the pandemic in Brazil, the North region was the most affected, accounting for the highest incidence, followed by CE and MA in the Northeastern region. Once the entry of the virus was confirmed within each state, the capitals were the most affected cities initially [19, 18], emerging as the epicenter of the epidemic in each state (see Supplementary Figures 2 and 3). Subsequently, SARS-CoV-2 disseminated throughout the inland cities with a different speed, as shown in the top panel plots of Supplementary Figures 2 and 3, where the incidence of COVID-19 is reported for the Brazilian states, capitals and inland cities.

### Governmental measures and population adherence

Next, we evaluated the timing of governmental interventions on the number of cases, the breadth of these interventions as measured by the stringency index, and their effects on influencing people’s behavior, particularly adhe-sion to social distancing recommendations.

A total of 707 regulations published by the 27 Brazilian state governments were annotated according to the methods described to construct the stringency index. The first measures enforcement of the state governments were implemented between March 11 and March 24, with exception of MT that present a larger time window interval, as shown in Figure 1A and Supplementary Table 2. The states of TO, RR, PI, PA, MT, AP, RO, MA, PB and AC adopted measures even before the first registered case. In contrast, some regions which were first affected by COVID-19 were among those that delayed the implementation of measures to contain viral spread. For instance, São Paulo adopted measures only 2 weeks after the confirmation of the first case, on March 13, the same day of community transmission declaration in the state. A similar scenario occurred in Rio de Janeiro, where the first restriction measures were only implemented in parallel to the declaration of community transmission.

Among the measures classified in Table 1, strict quarantine measures (*O*_4_), restrictions on public transportation (*C*_3_) and mandatory use of mask (*C*_3_) were the most weakly implemented measures enforced by the states. Such measures contributed to an average level of 14% in stringency in the studied period, and were adopted only partially in AP, BA, CE, MA and MT. Only an average of 16.9% of public transportation measures were applied in states. In the collected decrees, SP, DF and MS did not enact restrictions on public transport and PA, PB, PI, RN, GO and ES had a very low average of 4% in such measures. On the other hand, the mandatory use of masks contributed, in average, to 33% of the index, with only one state (MS) failing to enact such measures (Supplementary Table 3).

To assess the dynamics of implementation, continuance and lifting of NPIs by Brazilian states, Figure 2 shows the variation of the stringency index over time for each region relative to the number of confirmed COVID-19 deaths per 100,000 inhabitants. All states increased stringency after an incidence of 10 cases per 100k inhabitants. In addition, the states of AP, AM, RR, CE and PE displayed the highest stringency index in the last period evaluated. Of note, those states were close to reaching full occupancy of their health care systems, with more than 75% of ICU occupation [46]. These results evidenced that heightening the level of measures only when the number of cases and hospitalizations is already mounting represents a flawed strategy unable to avert the impacts of a surge of COVID-19 in healthcare demands. Variable adherence to social isolation recommendations was seen across the states, with values of the SMRI close to 30% at the beginning of March, followed by a peak to around 60% at the end of that month. This was observed in both state capitals and inland cities. The evolution of the stringency index for each state, as well as the SMRI for capitals and inland cities is presented in the lower panel plots of Supplementary Figure 2 an 3, as well as in the supplementary web-page. Used in conjunction, both measures offer a quantitative evaluation of the degree of policies enforced by Brazilian state governments, as well as their effectiveness in reducing the circulation of people, which lead to decreased contacts. Our results showed that, once the SMRI reached its maximum, it was followed by a decreasing trend even with the maintenance of measures by state governments.

**Figure 2:**
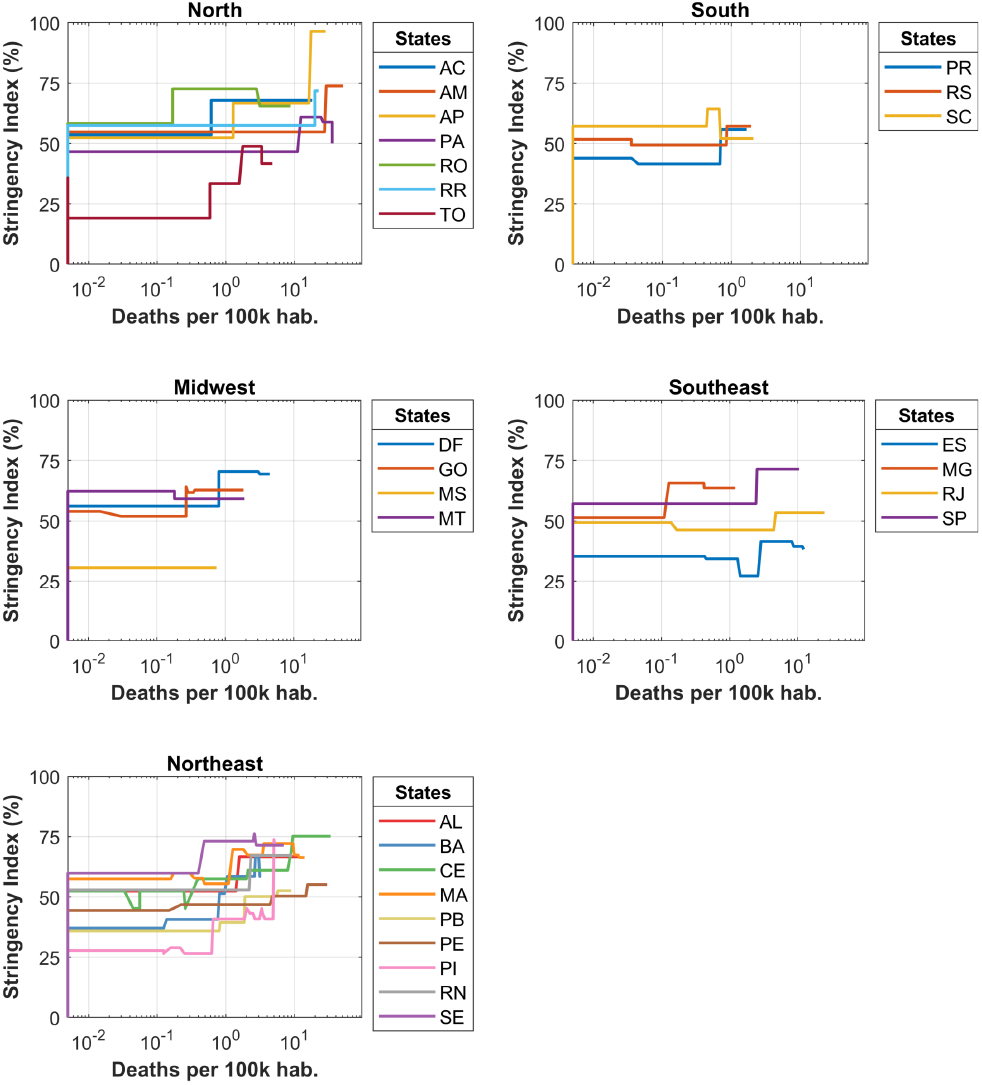
Evolution of the governmental measures adopted for each Brazilian state with respect to COVID-19 death incidence. The figure shows the variation of the stringency index over time for each state relative to the number of confirmed death per 100,000 inhabitants The number of confirmed deaths per 100,000 population is shown in logarithmic scale on the x-axis.

With respect to the breadth and intervention period of governmental measures, our results led to the identification of three stringency index pat-terns: 1) Increase-and-decrease (ID), where the stringency index increases initially, but is followed by the lifting of measures leading to its reduction (such as Santa Catarina in Figure 3a); 2) Increase-and-steady (IS), where stringency measures reach a peak that remains constant over time (depicted by São Paulo in Figure 3b); 3) Increase-and-increase (II), where the stringency index increases successively, probably a mechanism to cope with the accelerated growth of the epidemic in some regions (illustrated by Amapá in Figure 3c). Seven states, all located in the North (AC, AP and AM) and Northeast (BA, CE, PE and PI), followed the II pattern, while seven states, distributed throughout the Midwest (GO, MT), South (RS, SC), Southeast (ES) and North (RO, TO) regions conform to the ID pattern (Supplementary Figure 2 and 3). The remaining 13 states (AL, DF, MA, MG, MS, PA, PB, PR, RJ, RN, RR, SE and SP), distributed in all regions, followed the IS pattern. In addition, our results indicated that the reduction of the SMRI was smaller in states that followed both IS and II patterns (median reduction of −7.55% and −5.88%, respectively), compared to states that relaxed their measures according to the ID pattern (median reduction of −9.03%) (Figure 3d).

**Figure 3:**
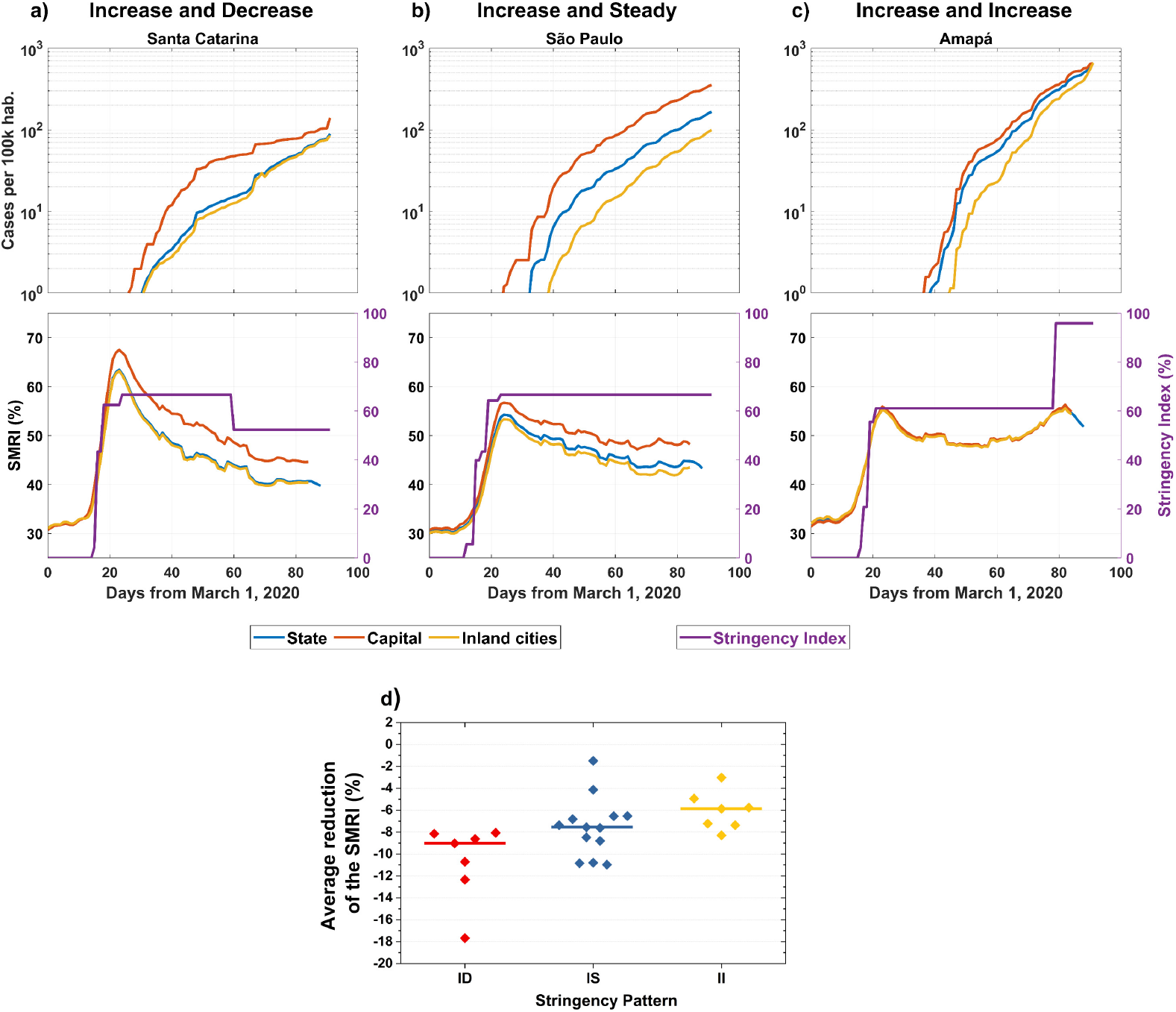
Illustrative examples of a general pattern observed for the behavior of stringency measures over time in Brazil. Upper panels show COVID-19 incidence and bottom panels exhibit the social mobility reduction and the stringency indexes over time for a) Santa Catarina (increase-and-decrease, ID), b) São Paulo (increase-and-steady, IS) and c) Amapá (increase-and-increase, II). The social mobility index is considered separately for capitals, inland cities and the whole state. Plots for the remaining Brazilian states are shown in Supplementary Figure 2 and 3. The average reduction in the SMRI according to the stringency pattern for all states is shown in panel d. For each category, the median is shown as a solid horizontal bar.

Of note, even states that promoted relaxation of policies, such as those that followed an ID pattern, maintained an average SMRI in the last week of May corresponding to 40.1 ± 2.0%, a value higher than that corresponding to the pre-pandemic SMRI in the first week of February, corresponding to a value of 28.7 ± 1.4%.

### Varying transmission rates of SARS-CoV-2 in Brazil

Lastly, we sought to comparatively evaluate the effects of governmental measures and population adherence to social distancing recommendations in the TR of SARS-CoV-2 throughout Brazilian states.

As shown earlier, states reported initial cases of COVID-19 and announced community transmission at different moments. Then, all regions reached a peak of measures implemented and corresponding SMRI on March, 2020. By evaluating these metrics in all of the 81 regions analyzed (state, capitals, inland cities), we defined the search interval for a possible first change in the transmission rate from March 13, to May 15, to include the date of the first measure enforced and the last announcement of community transmission (see Figure 1a). When more than one change in the TR took place, we defined the search interval to the last month of the studied period, i.e. from April 22 to May 22, 2020. More than two points of time changes were not considered in the time interval of study.

In Supplementary Table 4, we detail the number of days elapsed from the date of first implemented measure to the estimated date of TR change, and the variation from the first to the second (*β*_0_, *β*_1_) TR obtained by the SEIR model. Our results show that 26% of the states (BA, RN, DF, MG, RJ, RS and SC) presented a noticeable change in the TR up to 15 days after the implementation of the first NPI, 40.7% between 16 and 30 days, 14.8% (PA, PB, PE, PI) between 31 and 45 days and the remaining 18.5% (AC, RO, RR, AL and MS) of the states had a TR change only more than 46 days since the first implemented measure. Estimate TR changes occurred first in capitals, followed by inland cities (Supplementary Table 4).

The dynamic behaviour post intervention can be classified into DDD, DDI, DID, IDI and III, corresponding to decrease or increase in TR in the state as a whole, capital and inland, respectively. Around 62% of the states presented a DDD behavior, that is a decrease on the TR in the state, capital and inland cities. These states had an average TR decrease higher in the capitals (−49.67%) compared to the inland cities (−40.43%). The states of AP, RO, AL and RS (14.8%) displayed a DDI pattern, with an increase in cases observed for inland cities in May, 2020. On the other hand, the states of PI and SC showed an increase of the TR in capitals and an observed decrease in both state-level and inland cities (DID pattern). AC was the only state with an IDI pattern, while the remaining states (TO, SE and MS) presented with an III pattern. However, TO and SE later resembled a DDD pattern with a second change in TR. Furthermore, all states with an increase in TR in any of its regions (state, capital or inland cities) had either an ID or IS for the stringency index (as noted in the previous subsection).

In Figure 1B we show values for the basic reproduction number for each region. The lowest values of *ℛ*_0_ were for the regions that presented an increased TR, as showed before. For instance, TO and SE presented values of *ℛ*_0_ *<* 1. This indicates the lower transmission in the early stage of the disease spread in these states. The state of MS, that also had and III pattern, and the inland cities of SC all had the following lowest values of *ℛ*_0_ (less than 1.5). The inland cities of AC, AL, RO and RS showed values between 1.5 and 2. The remaining states that presented with a DDD pattern for the TR had *ℛ*_0_ *>* 2. Additionally, we can see that states that implemented the measures before the first reported case usually experienced delayed changes in the TR (after 8 April) compared to the remaining states and, with exception of AP, these states were the ones with an average lower value of *ℛ*_0_ as well.

To conclude, changes in the TR also impacted on the effective reproduction number causing an average reduction of −56.44% in capitals and of −46.43% in inland cities, after the first TR change In Figure 4 we show this impact for each state, their capitals and inland cities. Although a decrease in the average *ℛ*_*t*_ after the first TR reduction was observed in most of the states, in none of the regions the values of *ℛ*_*t*_ fell below one. Moreover, after the first TR change, the average level of *ℛ*_*t*_ was equal to 1.86 in the inland cities compared to 1.53 in capitals.

**Figure 4:**
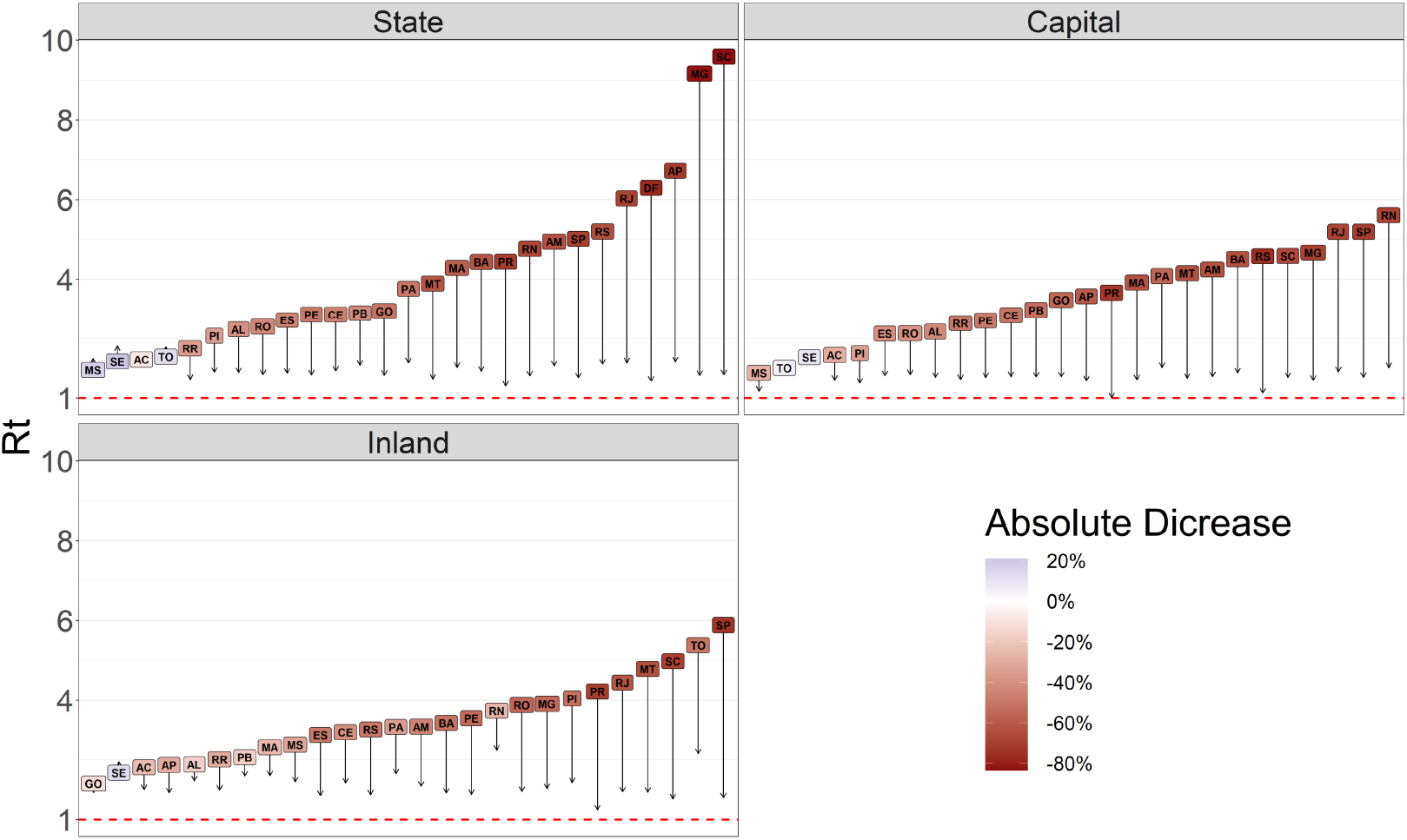
Impact of measures and popular adherence on the effective reproduction number in each Brazilian state. The plot depicts the decrease of the average of *ℛ*_*t*_ before and after the first TR change.

More details regarding the fitting of the data to the SEIR model produced in this work can be found in the supplementary web-page, for both capitals and inland cities of each state, as well as the entire state (Supplementary Figure 4). We highlight, in each plot, as vertical dashed red lines, the dates of transition from *β*_0_ to *β*_1_ (and *β*_1_ to *β*_2_, when applicable) (Supplementary Figure 4B). The blue dashed and full lines represent the evolution of the epidemic with a fixed transmission rate *β*_0_ and with both *β*_0_ and *β*_1_ (where *β*_2_ is included when suitable), respectively (Supplementary Figure 4B). The effective reproductive number is also presented for each state, capitals and inland cities (Supplementary Figure 4C). The black line represents the *ℛ*_*t*_ calculated with reported number of new cases; the blue dashed line represents the *ℛ*_*t*_ calculated with the new number of simulated cases obtained from the model. The variation of the TR highlights the variations of the trends of the effective reproductive number. Also shown are the estimates for stringency for each region and state (Supplementary Figure 4D), the moments of changes in *β* (Supplementary Figure 4D) and the goodness-of-fit (Supplementary Figure 4F), along with tables describing all these results (Supplementary Figure 4G). Finally, post assessment for all fit performances are available to evaluate reliability of the forecasts for a 15 day-period beyond May 22.

## 4. Discussion

In this work we evaluated the effects of non-pharmaceutical interventions and social mobility reduction patterns on the spread dynamics of SARS-CoV-2 throughout the 27 Brazilian states, by employing an underlying SEIR model to estimate the transmission rate of SARS-CoV-2. Our results show that the measures adopted, combined with the population adherence to restrict circulation (and therefore decrease contacts), contributed to the lowering of the TR in almost all states, an effect that was perceived in both capitals and inland cities. However, in spite of the continued maintenance of governmental restrictions in most regions, population adherence to isolation recommendations gradually decreased over time, even with the expansion of cases throughout the country. This was reflected in the *ℛ*(*t*) values, which we observed to have decreased in all states, but still insufficiently to consider SARS-CoV-2 transmission controlled in the country, as it remained above 1 for all Brazilian states, capital and inland cities in the studied period. Thus, public cooperation constitutes a particularly important challenge for tackling COVID-19 in low- and middle-income countries.

The analysis performed on the sub-measures implemented reveal that the Brazilian states lacked to endure measures restricting transportation (or did so in a very lax manner), strict lock-downs (as adopted by other countries), and even the mandatory use of masks at some places, the latter a low cost measure that plays an important rule to reduce the spread of the disease. Such restrictive policies have shown to significantly decrease the number of cases, deaths, and viral transmission in other countries [47, 48]. On the other hand, the economical costs imposed by harsher interventions is even more burdensome to developing countries, where large economical segments rely on consumption and services, usually involving physical contact, such as informal workers, tourism, service and retail businesses.

Once the entry of the virus was confirmed within each state, the capitals were the most affected initially, as shown by others as well [19, 18]. Then, viral spread continued at different rates, with most of inland cities presenting an average effective reproduction number higher than the capital after the first TR change. This is a concerning trend considering the large inequalities in the access to health services as well as their distribution in Brazil [49, 50], which tend to concentrate near state capitals. We also observed that downward changes on TRs occurred first in the capitals, followed by the remaining cities. This effect can also be due to the flux between cities, which was not accounted in our modeling approach. Accordingly, the TR observed in capitals should also affect that of inland cities, as suggested by a meta-population compartmental model [51], but the possibility of further waves of COVID-19 in these smaller cities, particularly with the lifting of measures, should not be ruled out. These results highlight the major role of state capitals as seeding spots, followed by the entry of cases towards smaller, inland cities. Capitals also tend to centralize international airports, ports, population density and industries [52, 53, 18, 54].

We identified common trends in the stringency index that allowed the disclosure of three patterns, with the majority of states conforming to an increase-and-steady pattern, in which the set of governmental policies adopted remained unaltered over time. States that enforced and maintained mitigation measures were likely to observe a less pronounced relaxation of stay-at-home advises by their population, as measured by the SMRI. These results suggest the intimate relationship between the magnitude of governmental measures and the population adherence to such measures, particularly since higher values of stringency implicate in decreased opportunities of public activities. However, individuals throughout all states, in both capitals and inland cities, still reduced their adherence to social isolation in the course of time. The politicization of COVID-19 in Brazil [55] could have had an impact on people’s behavior and compliance with sanitary recommendations, particularly when individuals downplay the health risks imposed by SARS-CoV-2, as has been suggested for other countries [56].

We also observed that even in states that conformed to an increase-and-decrease stringency index pattern, at least a part of the population still maintained adherence to isolation. More studies are warranted to evaluate if this trend associates with specific age-groups, such as the elderly, employment status, such as individuals that have the possibility to continue working from home, education level or perception of risks around COVID-19.

The present study confirmed the findings about early implementation of interventions to mitigate the disease spread [28, 39]. In fact, the states that anticipated applied restriction of the population were the ones with lower value of *ℛ*_0_, representing a more controlled situation if compared to the other states. Nevertheless, with few exceptions, they were states that had an ID or IS pattern of mobility, and therefore the efforts were not sufficient to bring the transmission rate to control levels, as assessed by the *ℛ*_*t*_ values. However, even with late TR change detection in these states, overall delays must be noticed. In fact, in the scenario of an unfolding epidemic, the delay in notification systems impacts the perception of the usefulness of implemented measures. Our results reveal the magnitude of this perception, where only 26% of the states showed a change on the TR with a maximum delay of 15 days for the perception of the first peak of enforced policies, with the remaining states having a significantly larger delay. These results call attention to this problem and highlight the importance of improving the surveillance system to better evaluate the impact of implemented measures and optimise their strictness specially in countries with very low resource levels [57].

Our work has some limitations. First, in order to estimate TRs (and changes thereof) we relied upon a generalized form of the SEIR model which explicitly considers asymptomatic/non-detected infections. Thus, albeit the estimates of model parameters (or their respective search intervals) were informed by the literature for other countries, they could be different from the reality of the ongoing epidemic in Brazil. However, while the true extent of SARS-CoV-2 transmission by asymptomatic/non-detected and pre-symptomatic individuals is still debated, current reports conclude that it is an important route of transmission [5, 58, 33]. It is important to note that levels of COVID-19 detection may vary over time due to the differing testing capacity of each region, a data that to our knowledge is currently unavailable for the country. Our reflects this issue by the parameter *p*. However, the availability of data about the number of tests performed over time is a limitation. Also, there are noticeable notification delays that also present with different magnitudes throughout the regions in Brazil. In fact, Brazil has a total of 5,570 municipalities, this generates heterogeneities on the reporting date of the case notification. A systematic analysis to infer the reporting delay would require availability of data to at least measure a mean report delay interval, as others have done [28]. Additionally, this delay may change over time due to improvement or deterioration in the healthcare system. These are key points that should be included to overcome this type of bias and correctly estimate its uncertainties. This limitation may impact on the perception of the implemented measures as well as compromise the planning of new ones. However, our work further motivates the importance of an improved notification system for Brazil and other countries, which will contribute to better mitigate the current and future pandemics.

We used mobility data from mobile phones as proxies of social isolation as measured by the SMRI. In particular, the sample space of devices monitored using this technology cannot be considered a representative population sample, as state/cities with superior economic status will probably exhibit increased technology adoption by their populations, leading to better accuracy of the mobility patterns in these regions. This is in contrast to rural areas, for instance, where mobile phone usage is limited [59]. However, considering the general widespread use of mobile phones in the country (with estimates that 60% of adults report owning a smartphone [60]), the general trends observed in our work should not be drastically altered by more accurate measurements of social mobility reduction. Lastly, the evaluation of all measures enforced is not trivial, and in our work we have only included the use of mask as health etiquette policies, with other NPIs such as sanitary cleaning of public and private places, the obligatory availability of 70% alcohol in stores, as well as economic measures such as emergency payments for the self-employed, reduction of taxes and others were not evaluated. Still, proper evaluation of these categories should be assessed.

In sum, our results point to the importance of timely deployment of interventions in curbing the first-wave of the COVID-19 epidemic in Brazil. Yet, population adherence represents a crucial factor for the success of this effort and represents a major challenge in low- and middle-income countries.

## Data Availability

Codes used to produce the results presented herein, and related datasets, are available as supplementary material and in a public GitHub repository.

https://github.com/cidacslab/Mathematical-and-Statistical-Modeling-of-COVID19-in-Brazil.git

## Acknowledgements

This study was financed in part by the Coordenação de Aperfeiçoamento de Pessoal de Nível Superior – Brazil (CAPES) – Finance Code 001. STRP was supported by an International Cooperation grant (process number INT0002/2016) from Bahia Research Foundation (FAPESB). STRP and RFSA were supported by the National Institute of Science and Technology - Complex Systems from CNPq, Brazil. JFO was supported by the Center of Data and Knowledge Integration for Health (CIDACS) through the Zika Platform - a long-term surveillance platform for Zika virus and microcephaly (Unified Health System (SUS), Brazilian Ministry of Health). DCJ acknowledges a Scientific Initiation scholarship from CNPq (process number 117568/2019-8).MSS acknowledges a Scientific Initiation scholarship from CNPq (process number 116731/2019-2). The authors acknowledge the helpful suggestions from members of the CoVida Network (http://covid19br.org), in special to contributors to the Rede CoVida Modelling Task-force: Aureliano S. S. Paiva, Caio P. Castro, Gabriela L. Borges, Gervásio F. Santos, José G. B. Castro, José G. V. Miranda, Maira L. Souza, Maria Yury Ichihara, Matheus F. Torquato, Maurício L. Barreto, Raphael S. do Rosário, Rafael V. Veiga and Rosemeire L. Fiaconne. We acknowledge the InLoco team, namely José Luciano Melo, Raíza Oliveira, Afonso Delgado, Abel Borges, André De’Carli, Hector Pinheiro, Lucas Rufino, Gabriel Teotonio and Luiza Botelho for providing raw files of the Social mobility reduction data used here. We also acknowledge the open data initiative of Brasil.io (https://brasil.io/datasets/).

## Authors’ contributions

Conceptualization: LLC, NBS, FACP, PIPR, JFO Data curation: DCPJ, MSR, MSS, VAFS, FACP, ARA, PIPR, JFO Formal analysis: DCPJ, MSR, MSS, LLC, NBS, IHS, VAFS, FACP, ARA, ASSA, STRP, RFSA, PIPR, JFO Investigation: LLC, NBS, FACP, PIPR, JFO Methodology: DCPJ, MSR, MSS, LLC, NBS, VAFS, FACP, ASSA, STRP, RFSA, PIPR, JFO Project administration: STRP, RFSA, PIPR, JFO Software: DCPJ, MSR, MSS, FACP, ARA, ASA, JFO Validation: DCPJ, MSR, MSS, LLC, NBS, VAFS, FACP, ARA, ASSA, STRP, RFSA, PIPR, JFO Visualization: MSR, MSS, ARA, PIPR, JFO Writing - original draft: LLC, STRP, PIPR, JFO Writing - review and editing: DCPJ, MSR, MSS, LLC, NBS, VAFS, FACP, ARA, ASSA, STRP, RFSA, PIPR, JFO

## Supplementary Figures

**Supplementary Figure 1:**
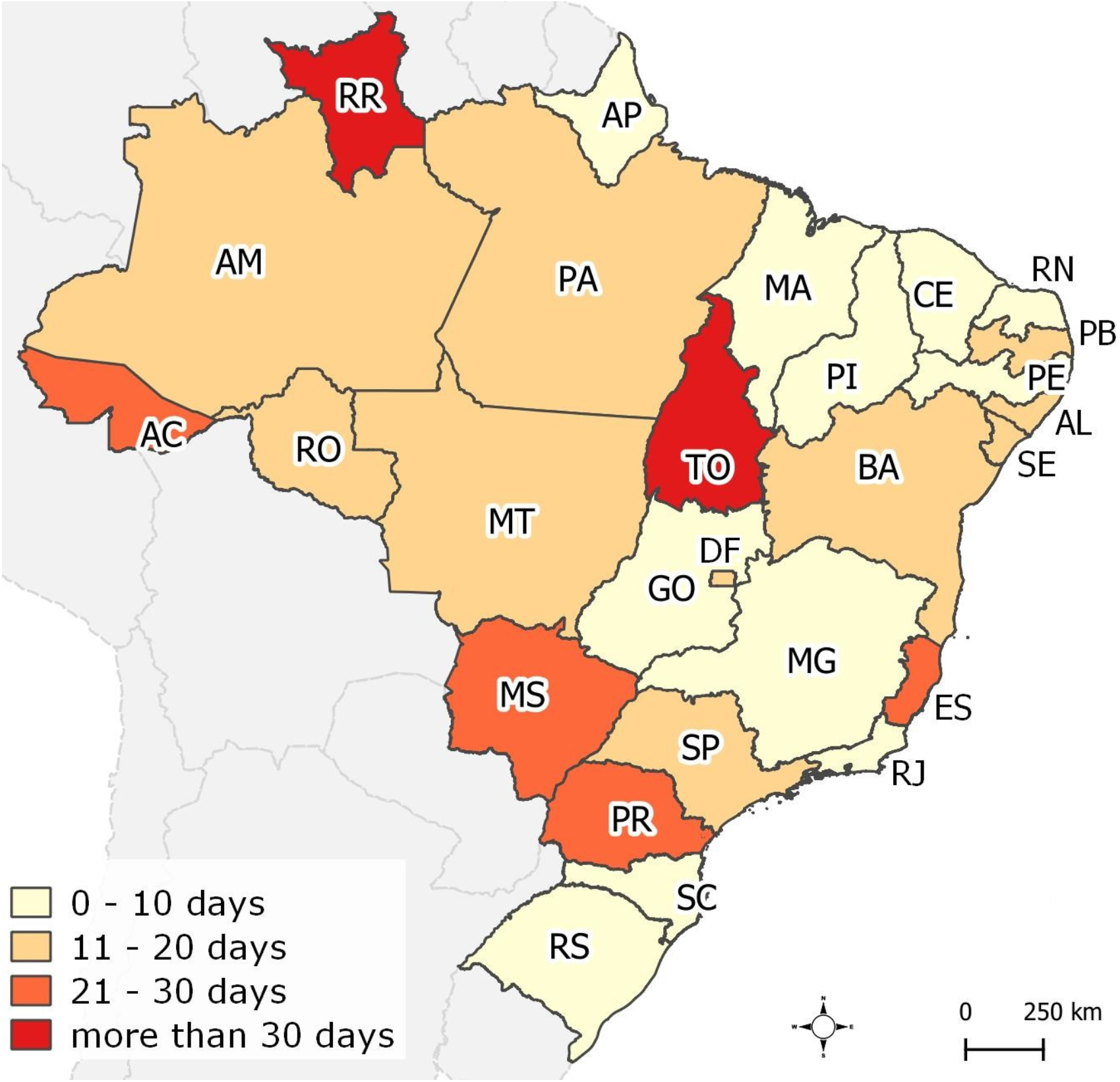
Dissemination of COVID-19 in Brazil. (a) Date of first confirmed COVID-19 case per state and (b) number of days elapsed from the first identified case to the establishment of community transmission in the state. Two-letter state abbreviations are as follows: AC, Acre; AL, Alagoas; AP, Amapá; AM, Amazonas; BA, Bahia; CE, Ceará; DF, Distrito Federal; ES, Espírito Santo; GO, Goiás; MA, Maranhão; MT, Mato Grosso; MS, Mato Grosso do Sul; MG, Minas Gerais; PA, Pará; PB, Paraíba; PR, Paraná; PE, Pernambuco; PI, Piauí; RJ, Rio de Janeiro; RN, Rio Grande do Norte; RS, Rio Grande do Sul; RO, Rondônia; RR, Roraima; SC, Santa Catarina; SP, São Paulo; SE, Sergipe; TO, Tocantins.

**Supplementary Figure 2:**
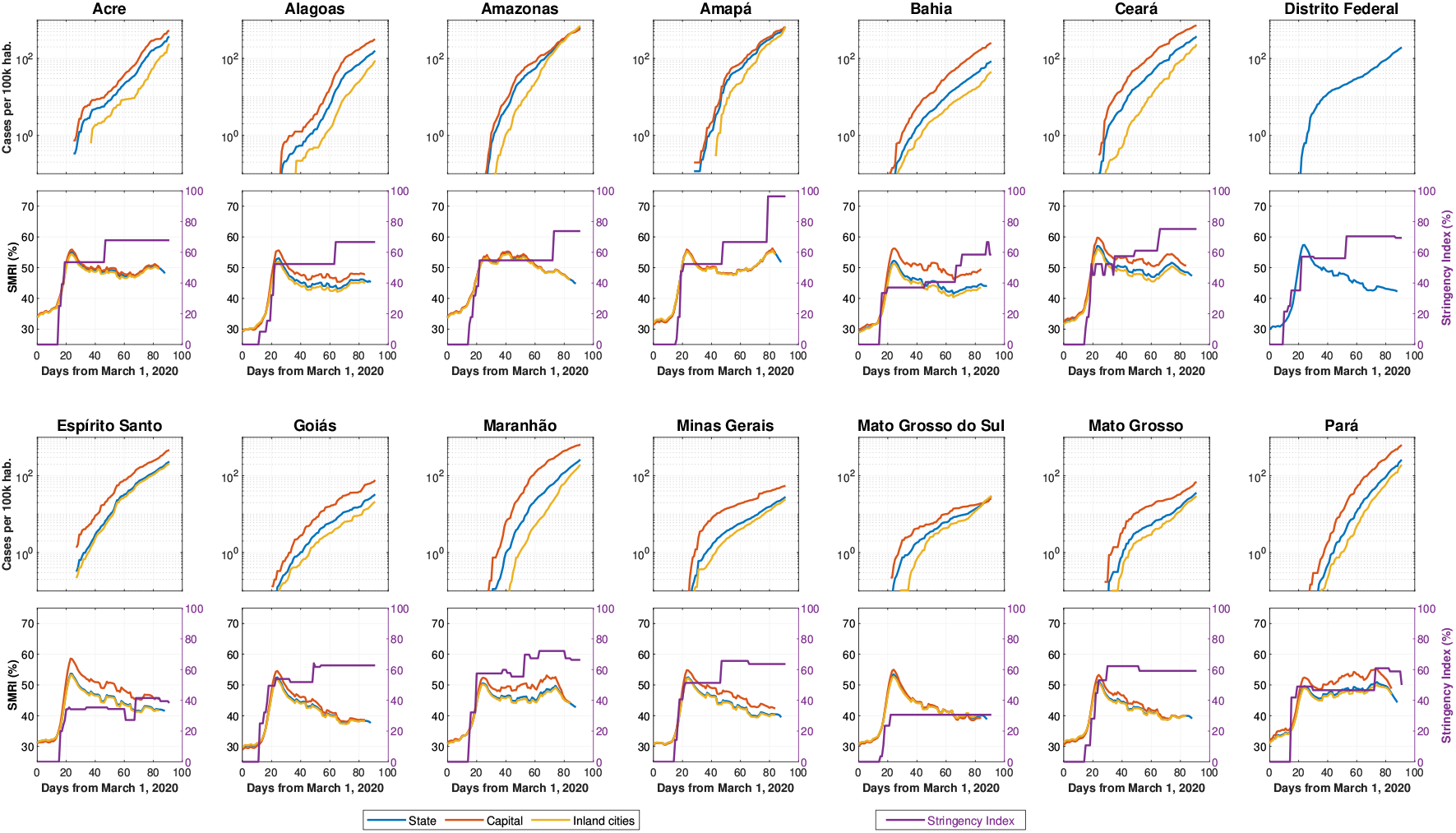
COVID-19 incidence per state, their capitals and remaining inland cities (upper plots). The bottom subplots exhibit the Social mobility reduction index, considered separately for capitals, inland cities and the whole state, and the stringency index, for the state measures, over time.

**Supplementary Figure 3:**
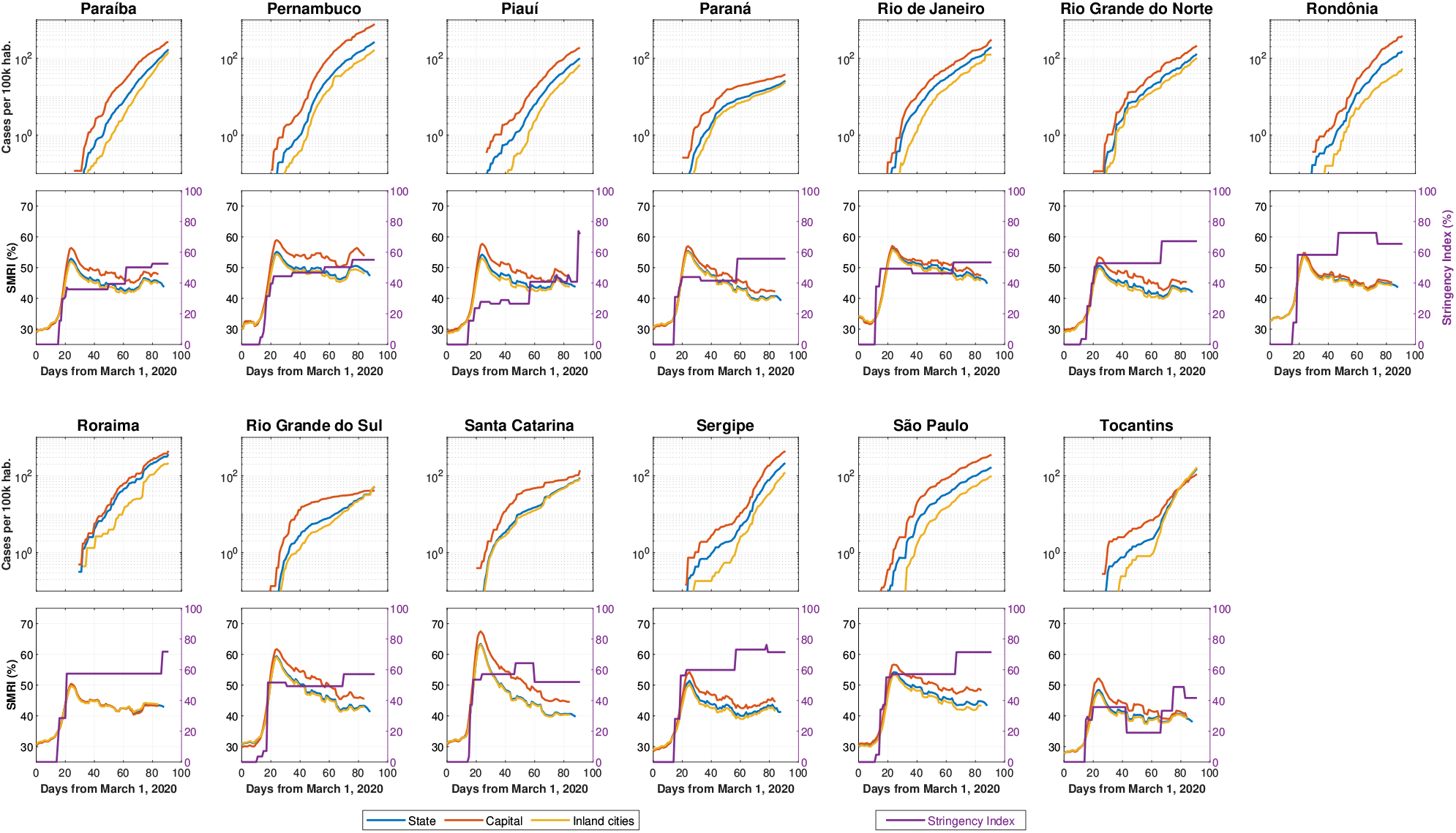
COVID-19 incidence per state, their capitals and remaining inland cities (upper plots). The bottom subplots exhibit the Social mobility reduction index, considered separately for capitals, inland cities and the whole state, and the stringency index, for the state measures, over time.

**Supplementary Figure 4.**
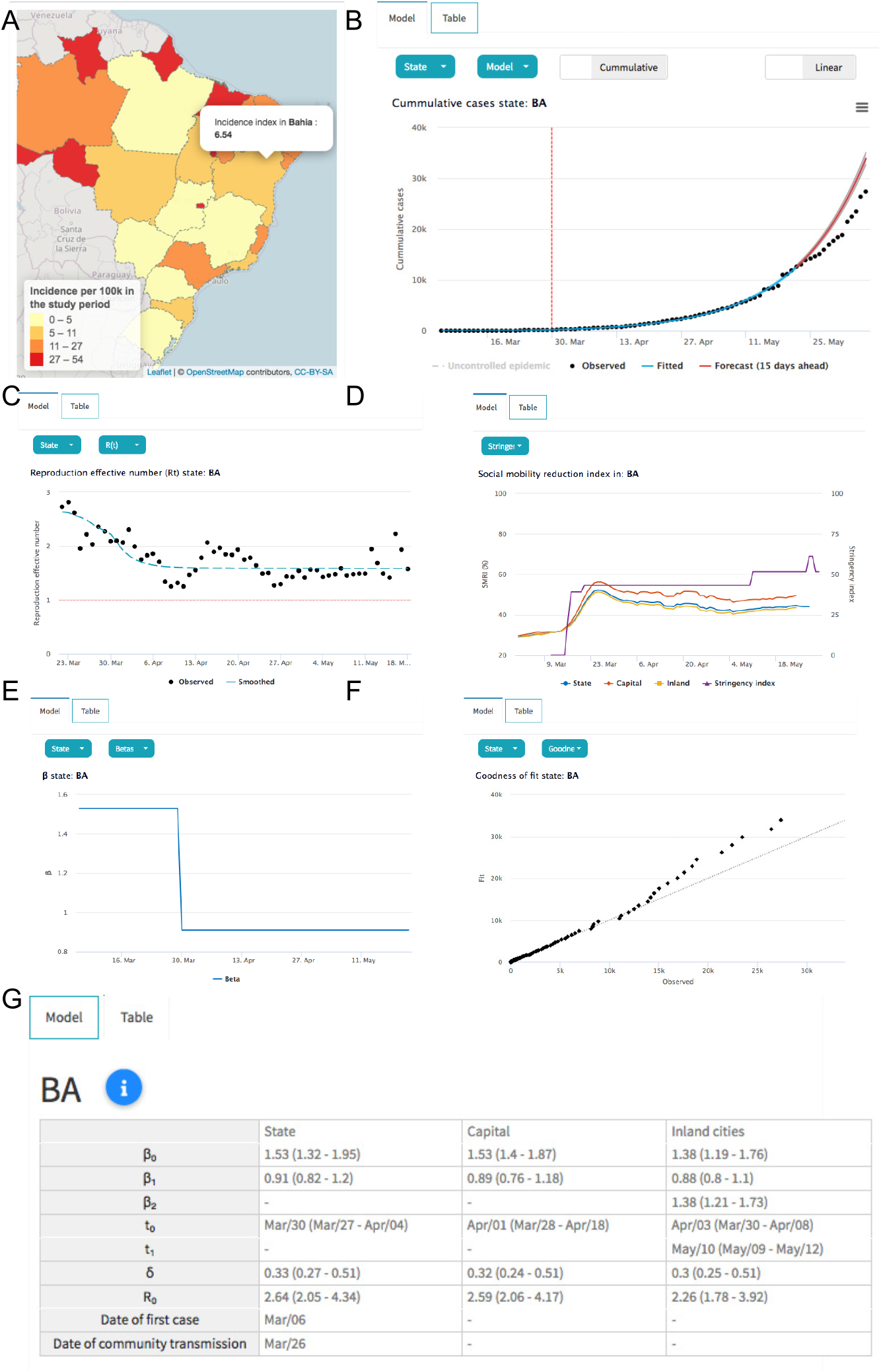

## Supplementary Tables

**Supplementary Table 1:**
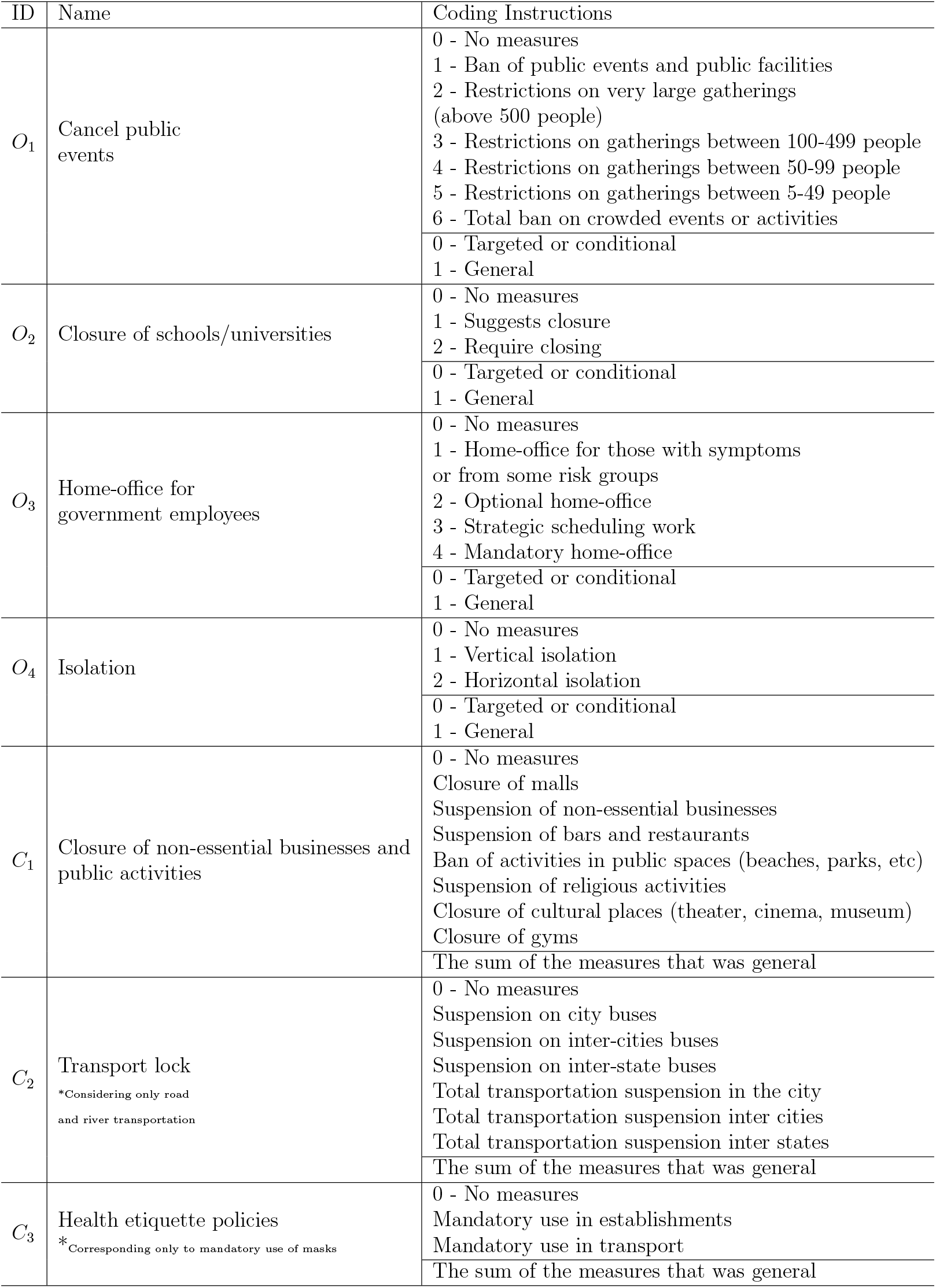
Classification of the measures implemented in Brazilian states as a response to the COVID-19 spread.

| ID | Name | Coding Instructions |
| --- | --- | --- |
| $O_1$ | Cancel public events | 0 - No measures<br>1 - Ban of public events and public facilities<br>2 - Restrictions on very large gatherings (above 500 people)<br>3 - Restrictions on gatherings between 100-499 people<br>4 - Restrictions on gatherings between 50-99 people<br>5 - Restrictions on gatherings between 5-49 people<br>6 - Total ban on crowded events or activities |
|  |  | 0 - Targeted or conditional<br>1 - General |
| $O_2$ | Closure of schools/universities | 0 - No measures<br>1 - Suggests closure<br>2 - Require closing |
|  |  | 0 - Targeted or conditional<br>1 - General |
| $O_3$ | Home-office for government employees | 0 - No measures<br>1 - Home-office for those with symptoms or from some risk groups<br>2 - Optional home-office<br>3 - Strategic scheduling work<br>4 - Mandatory home-office |
|  |  | 0 - Targeted or conditional<br>1 - General |
| $O_4$ | Isolation | 0 - No measures<br>1 - Vertical isolation<br>2 - Horizontal isolation |
|  |  | 0 - Targeted or conditional<br>1 - General |
| $C_1$ | Closure of non-essential businesses and public activities | 0 - No measures<br>Closure of malls<br>Suspension of non-essential businesses<br>Suspension of bars and restaurants<br>Ban of activities in public spaces (beaches, parks, etc)<br>Suspension of religious activities<br>Closure of cultural places (theater, cinema, museum)<br>Closure of gyms |
|  |  | The sum of the measures that was general |
| $C_2$ | Transport lock<br>*Considering only road and river transportation | 0 - No measures<br>Suspension on city buses<br>Suspension on inter-cities buses<br>Suspension on inter-state buses<br>Total transportation suspension in the city<br>Total transportation suspension inter cities<br>Total transportation suspension inter states |
|  |  | The sum of the measures that was general |
| $C_3$ | Health etiquette policies<br>*Corresponding only to mandatory use of masks | 0 - No measures<br>Mandatory use in establishments<br>Mandatory use in transport |
|  |  | The sum of the measures that was general |

**Supplementary Table 2:** SARS-CoV-2 entry in Brazil and association with NPIs and peak stringency

| State | Date of first case | Date of community transmission | Interval between initial NPIs and first stringency peak (days)<sup>†</sup> |
| --- | --- | --- | --- |
| Acre | Mar/17 | Apr/09 [61] | Mar 16-20 (4 days) |
| Amazonas | Mar/13 | Mar/28 [62] | Mar 16-23 (7 days) |
| Amapá | Mar/20 | Mar/28 [63] | Mar 17-22 (5 days) |
| Pará | Mar/18 | Mar/30 [64] | Mar 16-20 (4 days) |
| Rondonia | Mar/20 | Mar/31 [65] | Mar 17-20 (3 days) |
| Roraima | Mar/21 | May/02 [45] | Mar 16-22 (6 days) |
| Tocantins | Mar/18 | Apr/24 [66] | Mar 16-21 (5 days) |
| Alagoas | Mar/08 | Mar/28 [67] | Mar 13-23 (10 days) |
| Bahia | Mar/06 | Mar/19 [68] | Mar 16-21 (5 days) |
| Ceará | Mar/16 | Mar/20 [69] | Mar 16-20 (4 days) |
| Maranhão | Mar/20 | Mar/30 [70] | Mar 16-21 (5 days) |
| Paraíba | Mar/18 | Apr/04 [71] | Mar 17-22 (5 days) |
| Pernambuco | Mar/12 | Mar/17 [72] | Mar 14-23 (9 days) |
| Piauí | Mar/19 | Mar/20 [2] | Mar 16-24 (8 days) |
| Sergipe | Mar/14 | Mar/25 [73] | Mar 16-24 (8 days) |
| Rio Grande do Norte | Mar/12 | Mar/20 [2] | Mar 13-24 (11 days) |
| Goiás | Mar/12 | Mar/15 [74] | Mar 13-24 (11 days) |
| Distrito Federal | Mar/07 | Mar/26 [75] | Mar 11-21 (10 days) |
| Mato Grosso | Mar/20 | Mar/31 [76] | Mar 16-31 (15 days) |
| Mato Grosso do Sul | Mar/14 | Apr/04 [77] | Mar 16-23 (7 days) |
| Espírito Santo | Mar/06 | Mar/30 [78] | Mar 17-21 (4 days) |
| Minas Gerais | Mar/08 | Mar/18 [79] | Mar 16-23 (7 days) |
| Rio de Janeiro | Mar/05 | Mar/13 [80] | Mar 13-16 (3 days) |
| São Paulo | Feb/26 | Mar/13 [80] | Mar 13-20 (7 days) |
| Paraná | Mar/12 | Apr/09 [44] | Mar 16-23 (7 days) |
| Rio Grande do Sul | Mar/10 | Mar/20 [81] | Mar 16-21 (5 days) |
| Santa Catarina | Mar/12 | Mar/18 [82] | Mar 16-19 (3 days) |

All dates refer to the year 2020; †, interval between the first NPIs enacted by states and the first peak of the stringency index.

**Supplementary Table 3:** Summary of the stringency index and subindexes for each state.

| State | $\iota$ | | | $O_1$ | | | $O_2$ | | | $O_3$ | | | $O_4$ | | | $C_1$ | | | $C_2$ | | | $C_3$ | | |
| --- | --- | --- | --- | --- | --- | --- | --- | --- | --- | --- | --- | --- | --- | --- | --- | --- | --- | --- | --- | --- | --- | --- | --- | --- |
|  | Mean | Max | SD | Mean | Max | SD | Mean | Max | SD | Mean | Max | SD | Mean | Max | SD | Mean | Max | SD | Mean | Max | SD | Mean | Max | SD |
| AC | 0.52 | 0.68 | 0.23 | 0.68 | 1.00 | 0.36 | 0.83 | 1.00 | 0.37 | 0.86 | 1.00 | 0.35 | 0.0 | 0.0 | 0.0 | 0.81 | 1.00 | 0.39 | 0.21 | 0.25 | 0.09 | 0.25 | 0.50 | 0.25 |
| AM | 0.49 | 0.74 | 0.23 | 0.49 | 0.83 | 0.25 | 0.84 | 1.00 | 0.36 | 0.80 | 1.00 | 0.39 | 0.0 | 0.0 | 0.0 | 0.79 | 1.00 | 0.40 | 0.27 | 0.33 | 0.13 | 0.21 | 1.00 | 0.41 |
| AP | 0.59 | 0.96 | 0.29 | 0.28 | 1.00 | 0.31 | 0.91 | 1.00 | 0.37 | 0.90 | 1.00 | 0.38 | 0.16 | 1.00 | 0.35 | 0.89 | 1.00 | 0.39 | 0.48 | 0.75 | 0.20 | 0.54 | 1.00 | 0.50 |
| PA | 0.47 | 0.61 | 0.13 | 0.78 | 0.83 | 0.20 | 0.94 | 1.00 | 0.24 | 0.47 | 0.50 | 0.12 | 0.0 | 0.0 | 0.0 | 0.83 | 0.93 | 0.24 | 0.03 | 0.17 | 0.06 | 0.23 | 1.00 | 0.42 |
| RO | 0.59 | 0.73 | 0.20 | 0.74 | 0.83 | 0.26 | 0.93 | 1.00 | 0.26 | 0.89 | 1.00 | 0.31 | 0.0 | 0.0 | 0.0 | 0.78 | 1.00 | 0.34 | 0.22 | 0.25 | 0.08 | 0.55 | 1.00 | 0.50 |
| RR | 0.53 | 0.57 | 0.16 | 0.94 | 1.00 | 0.24 | 0.93 | 1.00 | 0.26 | 0.87 | 1.00 | 0.34 | 0.0 | 0.0 | 0.0 | 0.74 | 0.86 | 0.29 | 0.14 | 0.17 | 0.06 | 0.06 | 1.00 | 0.24 |
| TO | 0.30 | 0.49 | 0.12 | 0.29 | 1.00 | 0.45 | 0.94 | 1.00 | 0.24 | 0.27 | 0.75 | 0.14 | 0.1 | 0.5 | 0.2 | 0.05 | 0.50 | 0.15 | 0.14 | 0.25 | 0.09 | 0.30 | 1.00 | 0.46 |
| AL | 0.47 | 0.67 | 0.22 | 0.83 | 1.00 | 0.35 | 0.78 | 1.00 | 0.42 | 0.42 | 0.50 | 0.17 | 0.0 | 0.0 | 0.0 | 0.82 | 1.00 | 0.38 | 0.13 | 0.17 | 0.07 | 0.31 | 1.00 | 0.47 |
| BA | 0.39 | 0.67 | 0.19 | 0.57 | 0.67 | 0.24 | 0.85 | 1.00 | 0.36 | 0.21 | 0.25 | 0.09 | 0.12 | 0.50 | 0.21 | 0.38 | 1.00 | 0.18 | 0.20 | 0.25 | 0.10 | 0.42 | 1.00 | 0.47 |
| CE | 0.52 | 0.75 | 0.25 | 0.83 | 1.00 | 0.36 | 0.83 | 1.00 | 0.37 | 0.71 | 1.00 | 0.41 | 0.15 | 0.50 | 0.23 | 0.74 | 1.00 | 0.36 | 0.26 | 0.58 | 0.22 | 0.12 | 0.25 | 0.13 |
| MA | 0.58 | 0.72 | 0.18 | 0.94 | 1.00 | 0.24 | 0.93 | 1.00 | 0.26 | 0.89 | 1.00 | 0.29 | 0.0 | 0.0 | 0.0 | 0.66 | 0.86 | 0.26 | 0.16 | 0.33 | 0.13 | 0.48 | 1.00 | 0.50 |
| PB | 0.39 | 0.53 | 0.14 | 0.31 | 0.33 | 0.09 | 0.9 | 1.00 | 0.3 | 0.70 | 0.75 | 0.20 | 0.0 | 0.0 | 0.0 | 0.37 | 0.43 | 0.15 | 0.03 | 0.17 | 0.06 | 0.40 | 1.00 | 0.46 |
| PE | 0.45 | 0.55 | 0.13 | 0.91 | 1.00 | 0.23 | 0.91 | 1.00 | 0.28 | 0.23 | 0.25 | 0.06 | 0.0 | 0.0 | 0.0 | 0.77 | 0.86 | 0.25 | 0.18 | 0.50 | 0.17 | 0.11 | 0.25 | 0.12 |
| PI | 0.31 | 0.74 | 0.13 | 0.04 | 1.00 | 0.16 | 0.94 | 1.00 | 0.24 | 0.02 | 1.00 | 0.16 | 0.0 | 0.0 | 0.0 | 0.74 | 1.00 | 0.29 | 0.03 | 0.17 | 0.06 | 0.43 | 1.00 | 0.50 |
| SE | 0.60 | 0.76 | 0.18 | 0.91 | 1.00 | 0.26 | 0.94 | 1.00 | 0.24 | 0.89 | 1.00 | 0.28 | 0.0 | 0.0 | 0.0 | 0.78 | 1.00 | 0.25 | 0.24 | 0.33 | 0.15 | 0.43 | 1.00 | 0.50 |
| RN | 0.52 | 0.67 | 0.17 | 0.75 | 0.83 | 0.22 | 0.93 | 1.00 | 0.26 | 0.89 | 1.00 | 0.28 | 0.0 | 0.0 | 0.0 | 0.70 | 0.79 | 0.25 | 0.07 | 0.08 | 0.03 | 0.30 | 1.00 | 0.46 |
| GO | 0.55 | 0.64 | 0.13 | 0.98 | 1.00 | 0.16 | 0.93 | 1.00 | 0.25 | 0.73 | 0.75 | 0.12 | 0.0 | 0.0 | 0.0 | 0.62 | 0.86 | 0.22 | 0.05 | 0.17 | 0.08 | 0.52 | 1.00 | 0.50 |
| DF | 0.54 | 0.70 | 0.22 | 0.84 | 1.00 | 0.31 | 0.91 | 1.00 | 0.29 | 0.80 | 1.00 | 0.38 | 0.0 | 0.0 | 0.0 | 0.78 | 1.00 | 0.33 | 0.0 | 0.0 | 0.0 | 0.43 | 1.00 | 0.50 |
| MT | 0.52 | 0.62 | 0.18 | 0.91 | 1.00 | 0.26 | 0.85 | 1.00 | 0.36 | 0.68 | 0.75 | 0.21 | 0.17 | 0.50 | 0.24 | 0.33 | 0.86 | 0.33 | 0.21 | 0.25 | 0.09 | 0.49 | 1.00 | 0.50 |
| MS | 0.27 | 0.31 | 0.09 | 0.0 | 0.0 | 0.0 | 0.88 | 1.00 | 0.30 | 0.91 | 1.00 | 0.27 | 0.0 | 0.0 | 0.0 | 0.13 | 0.14 | 0.04 | 0.0 | 0.0 | 0.0 | 0.0 | 0.0 | 0.0 |
| ES | 0.30 | 0.41 | 0.14 | 0.25 | 0.50 | 0.25 | 0.82 | 1.00 | 0.37 | 0.21 | 0.25 | 0.09 | 0.0 | 0.0 | 0.0 | 0.48 | 0.64 | 0.23 | 0.05 | 0.08 | 0.04 | 0.27 | 1.00 | 0.44 |
| MG | 0.54 | 0.66 | 0.17 | 0.74 | 0.83 | 0.26 | 0.91 | 1.00 | 0.28 | 0.94 | 1.00 | 0.24 | 0.0 | 0.0 | 0.0 | 0.34 | 0.43 | 0.13 | 0.29 | 0.33 | 0.11 | 0.55 | 1.00 | 0.50 |
| RJ | 0.48 | 0.53 | 0.08 | 0.98 | 1.00 | 0.16 | 0.98 | 1.00 | 0.16 | 0.49 | 0.50 | 0.08 | 0.0 | 0.0 | 0.0 | 0.60 | 0.79 | 0.16 | 0.16 | 0.17 | 0.04 | 0.16 | 0.50 | 0.24 |
| SP | 0.57 | 0.71 | 0.16 | 0.95 | 1.00 | 0.20 | 0.94 | 1.00 | 0.24 | 0.90 | 1.00 | 0.28 | 0.0 | 0.0 | 0.0 | 0.90 | 1.00 | 0.28 | 0.0 | 0.0 | 0.0 | 0.30 | 1.00 | 0.46 |
| PR | 0.45 | 0.56 | 0.14 | 0.63 | 0.67 | 0.16 | 0.94 | 1.00 | 0.24 | 0.47 | 0.50 | 0.12 | 0.0 | 0.0 | 0.0 | 0.50 | 0.57 | 0.19 | 0.17 | 0.33 | 0.09 | 0.41 | 1.00 | 0.50 |
| RS | 0.47 | 0.57 | 0.15 | 0.78 | 1.00 | 0.26 | 0.9 | 1.0 | 0.3 | 0.48 | 0.50 | 0.08 | 0.0 | 0.0 | 0.0 | 0.63 | 0.79 | 0.24 | 0.26 | 0.33 | 0.11 | 0.27 | 1.00 | 0.45 |
| SC | 0.51 | 0.64 | 0.15 | 0.93 | 1.00 | 0.26 | 0.9 | 1.0 | 0.3 | 0.44 | 0.50 | 0.14 | 0.0 | 0.0 | 0.0 | 0.59 | 1.00 | 0.44 | 0.46 | 0.50 | 0.13 | 0.27 | 0.50 | 0.25 |

**Supplementary Table 4:**
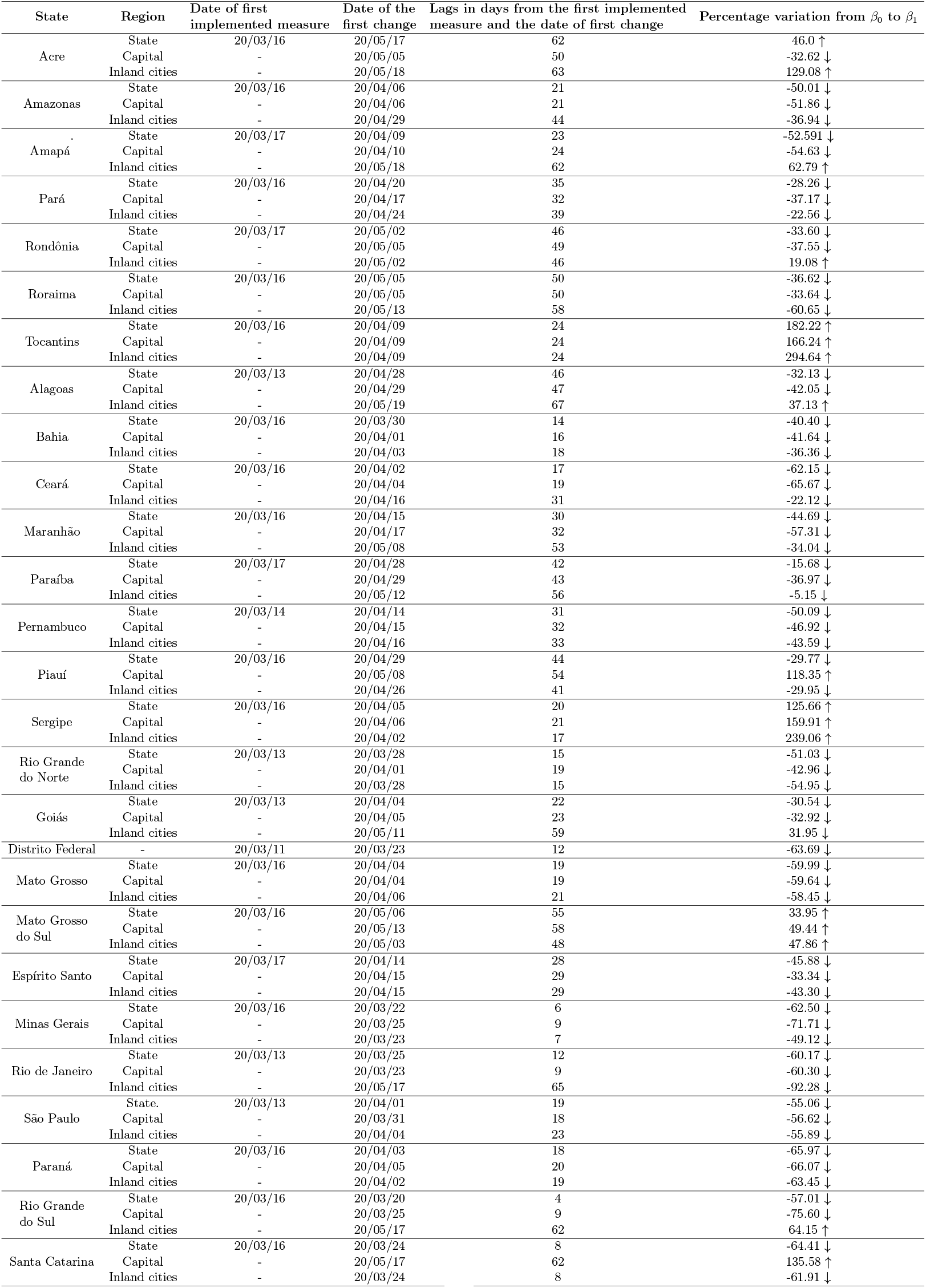
SARS-CoV-2 spread throughout 27 Brazilian states and effects on the transmission rate

**Supplementary Table 5.**
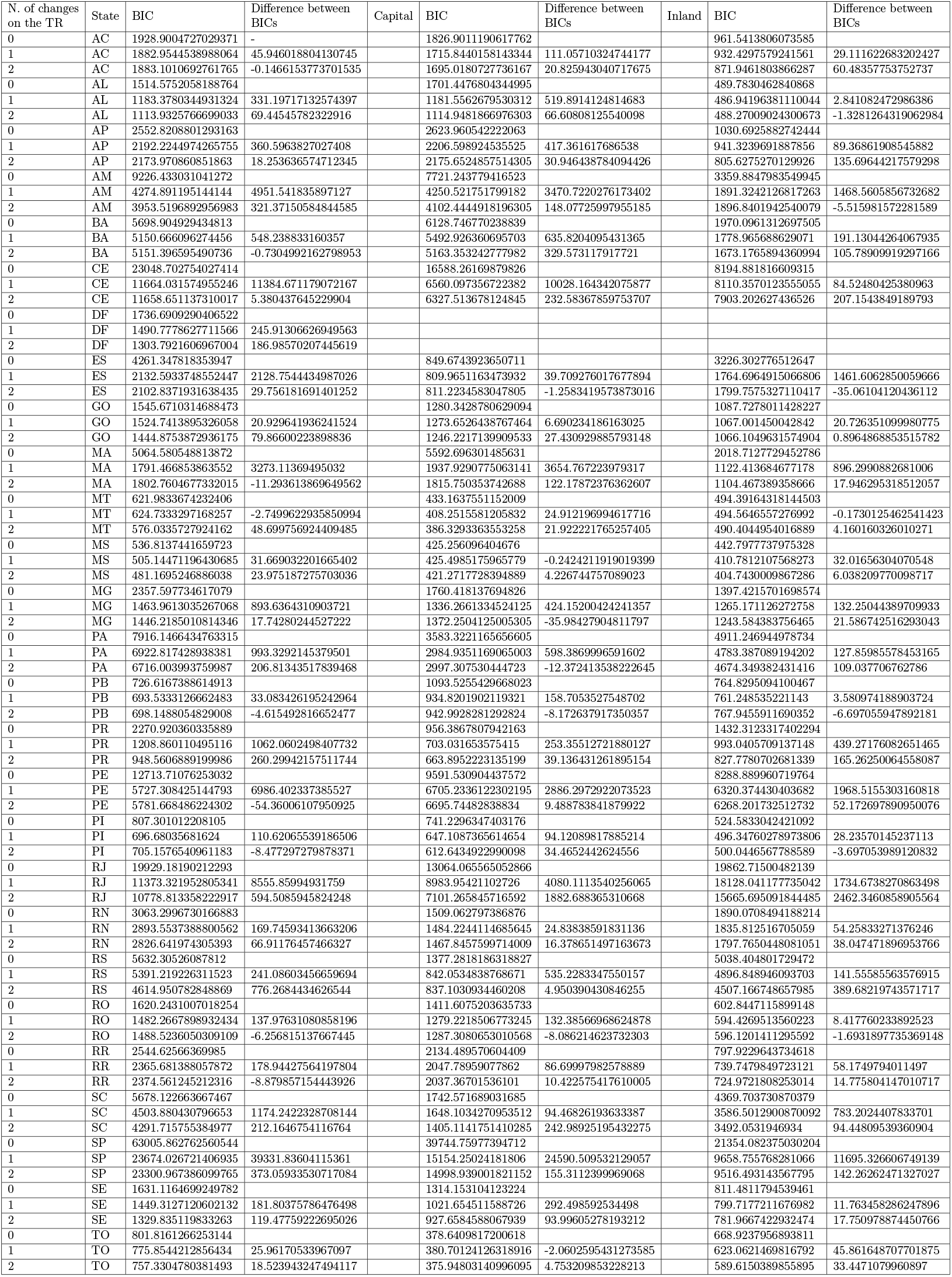

## Supplementary Note 1

### Stringency index calculation: examples

### Ordered measure

In this type of measure there is a well-defined order, it is a simple calculation. Scenarios for an ordered measure with N = 3 is shown in Supplementary Table 6.

A more restrictive measure applied in only in few regions has less value than a slightly less restrictive measure applied across the state, as we see in scenarios 3 and 4 above.

**Supplementary Table 6.**

| Scenario 1 |  | Scenario 2 |  | Scenario 3 |  | Scenario 4 |  |
| --- | --- | --- | --- | --- | --- | --- | --- |
| j | $G_O$ | j | $G_O$ | j | $G_O$ | j | $G_O$ |
| 0 | 1 | 1 | 1 | 2 | 1 | 3 | 0 |
| $\iota_O = 0/3 * (2 - 1) = 0$ | | $\iota_O = 1/3 * (2 - 1) = 0.33$ | | $\iota_O = 2/3 * (2 - 1) = 0.66$ | | $\iota_O = 3/3 * (2 - 0) = 0.5$ | |

### Cumulative measure

In this case, the evaluation of the cumulative measure *C*_*i*_ is the sum of the restrictions applied, and 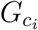 is the sum of the targets, N is the number of possible restrictions, counting with no restrictions. An example for a cumulative measure is shown in Supplementary Table 7.

**Supplementary Table 7.**

| Restriction to: | Restricted (1) / Open (0) | General (1) /<br>Target to a specific region (0) |
| --- | --- | --- |
| Shoppings | 1 | 0 |
| Open markets | 1 | 1 |
| Bar and restaurants | 1 | 1 |
| Gymns | 0 | 0 |
| Parks and beaches | 1 | 0 |
| Religious Temples | 1 | 1 |
| Theaters and cinemas | 1 | 0 |
| $N = 7$ | $s_C = 6$ | $s_{G_c} = 3$ |
| $\iota_C = (3 + 1.5)/(7 - 1) = 0.75$ | | |

## Supplementary Note 2

### Parameter sensitivity analysis

In this section, a sensitivity analysis is performed to evaluate the effects of the mathematical model parameters in the dynamics of *S, E, I*_*a*_, *I*_*s*_ and *R* compartments over time. By using an statistical variance-based method, described by Sobol (2001) [83], the sensitivity analysis of the SEIR model considers the following parameter vector

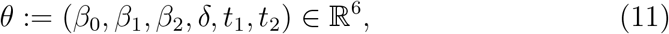

assuming that its elements are uniformly distributed (𝒰) in proper intervals:

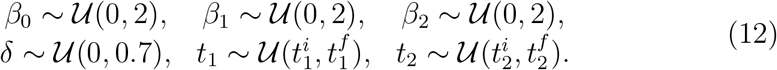

Note that the range of uniform distribution with respect to *t*_1_ and *t*_2_ varies according to the Brazilian states. In Fig. 5, the analysis is performed considering the Brazilian state of *Distrito Federal* as an example. For this example, the intervals for *t*_1_ and *t*_2_ are given as follows: *t*_1_ ∼ 𝒰(0, 50) and *t*_2_ ∼ 𝒰(30, 57). Despite the different dates of community transmission be-tween the Brazilian states, the influence of the analyzed parameters presented in Fig. 5, remains equivalent in relation to the other states.

### The method

To apply the statistical variance-based method, sample values is generated for the input factors shown in Eq. (11) by creating matrices *A* and *B*, each with size *N* × *n*, where *N* is the number of samples and *n* = 6 is the number of parameters being analyzed, given by

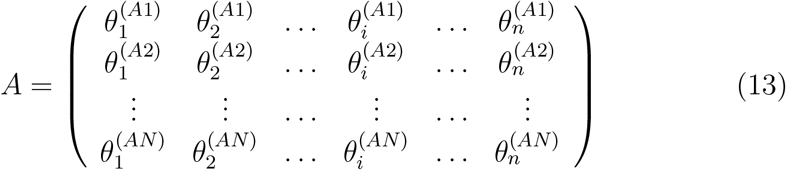

and

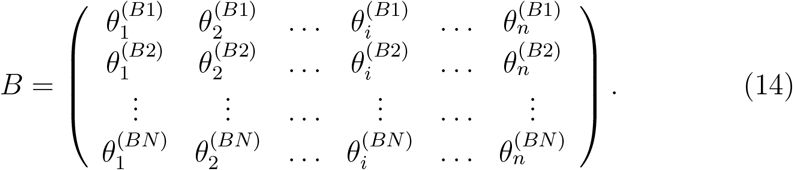

We then create *n* matrices 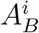, where column *i* comes from matrix *B* and all other *n* − 1 columns come from matrix *A*:

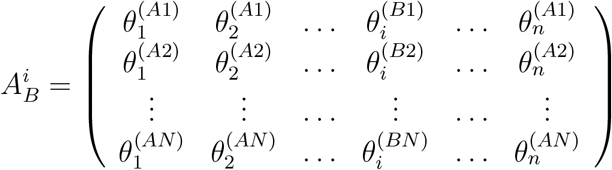

In the matrices *A, B* and 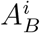, each row represents a set of parameter to be used as an input for the model. Numerical simulations are performed, and the output of the sample matrices *A, B* and 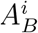 are stored as the vectors

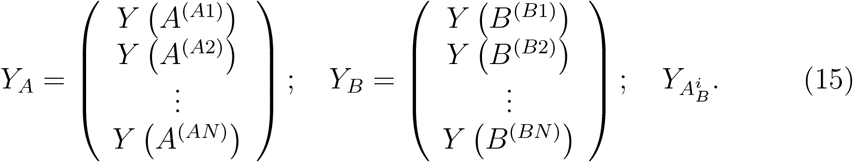

where *Y*_*A*_, *Y*_*B*_ and 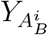 are output vectors.

The final step involves the calculation of the sensitivity indices, using the samples generated during the sampling scheme. We computed the total effect indices, given by

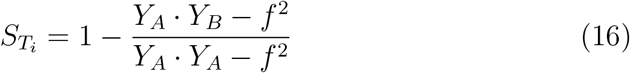

where *f* is defined as

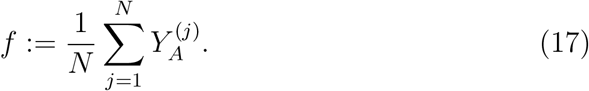

This index indicates the contribution of the parameter to the output of the model. The importance of each parameter *i* is proportional to the value of 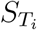, meaning that higher 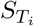 leads to a higher contribution to the model output [84]. Parameters with higher *S*_*T*_ need a more carefully calibration, as small error during the calibration can lead to larger errors to the predictions generated by the model. The total effect takes into account higher-order interactions among model variables; thus, correlation between variables can also be identified using this method. In addition, we also evaluated the influence of first-order effects, which do not consider interactions among variables, to the model output.

### Parameter sensitivity analysis: discussion

In Figure 5, the result of the sensitivity analysis over time is presented. The numerical simulations were performed using SALib library [85]. The experiment was conducted generating *N* = 15,000 parameter combinations, totaling 120,000 simulations of the model, and the result shows the evolution of the parameters according to *S, E, I*_*a*_, *I*_*s*_ and *R* compartments.

**Supplementary Figure 5:**
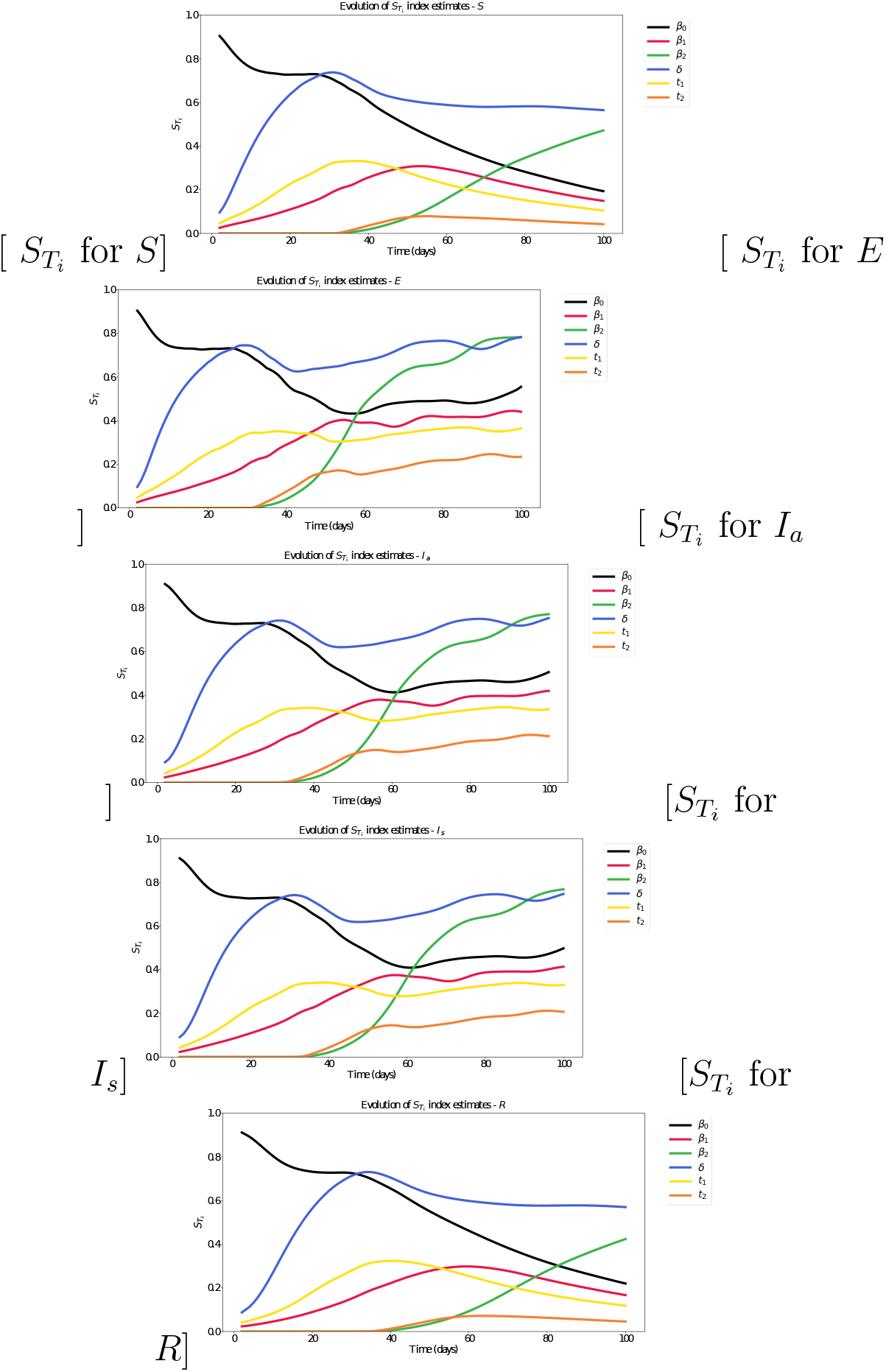
Sensitivity analysis study for *S, E, I*_*a*_, *I*_*s*_ and *R* compartments over time. The total effect index 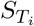 is shown for each evaluated parameter in each compartment of the SEIR model.

## Footnotes

10 Two-letter state abbreviations are as follows: AC, Acre; AL, Alagoas; AP, Amapá; AM, Amazonas; BA, Bahia; CE, Ceará; DF, Distrito Federal; ES, Espírito Santo; GO, Goiás; MA, Maranhão; MT, Mato Grosso; MS, Mato Grosso do Sul; MG, Minas Gerais; PA, Pará; PB, Paraíba; PR, Paraná; PE, Pernambuco; PI, Piauí; RJ, Rio de Janeiro; RN, Rio Grande do Norte; RS, Rio Grande do Sul; RO, Rondônia; RR, Roraima; SC, Santa Catarina; SP, São Paulo; SE, Sergipe; TO, Tocantins.

## References

[1] Organization WH. WHO Director-General’s opening remarks at the media briefing on COVID-19-11 March 2020. 2020;.

[2] Ministério da Saúde declara transmissão comunitária nacional;. Available from: https://www.saude.gov.br/noticias/agencia-saude/46568-ministerio-da-saude-declara-transmissao-comunitaria-nacional.

[3] Aquino EM, Silveira IH, Pescarini JM, Aquino R, Souza-Filho JAd. Social distancing measures to control the COVID-19 pandemic: potential impacts and challenges in Brazil. Ciência & Saúde Coletiva. 2020;25:2423–2446.

[4] Panovska-Griffiths J. Can mathematical modelling solve the current Covid-19 crisis? BMC Public Health. 2020;.

[5] Li R, Pei S, Chen B, Song Y, Zhang T, Yang W, et al. Substantial un-documented infection facilitates the rapid dissemination of novel coronavirus (SARS-CoV-2). Science. 2020;368(6490):489–493.

[6] Berger DW, Herkenhoff KF, Huang C, Mongey S. Testing and reopening in an SEIR model. Review of Economic Dynamics. 2020;in press.

[7] Roche B, Garchitorena A, Roiz D. The impact of lockdown strategies targeting age groups on the burden of COVID-19 in France. Epidemics. 2020;33:100424.

[8] Oliveira JF, Jorge DC, Veiga RV, Rodrigues MS, Torquato MF, da Silva NB, et al. Evaluating the burden of COVID-19 on hospital resources in Bahia, Brazil: A modelling-based analysis of 14.8 million individuals. Nature Communications. 2021;12:333.

[9] Ferguson N, Laydon D, Nedjati Gilani G, Imai N, Ainslie K, Baguelin M, et al. Impact of non-pharmaceutical interventions (NPIs) to reduce COVID19 mortality and healthcare demand; 2020.

[10] Moghadas SM, Shoukat A, Fitzpatrick MC, Wells CR, Sah P, Pandey A, et al. Projecting hospital utilization during the COVID-19 outbreaks in the United States. Proceedings of the National Academy of Sciences. 2020;117(16):9122–9126.

[11] Prem K, Liu Y, Russell TW, Kucharski AJ, Eggo RM, Davies N, et al. The effect of control strategies to reduce social mixing on outcomes of the COVID-19 epidemic in Wuhan, China: a modelling study. The Lancet Public Health. 2020;5:e261–e270.

[12] Leung K, Wu JT, Liu D, Leung GM. First-wave COVID-19 transmissibility and severity in China outside Hubei after control measures, and second-wave scenario planning: a modelling impact assessment. The Lancet. 2020;.

[13] Weitz JS, Beckett SJ, Coenen AR, Demory D, Dominguez-Mirazo M, Dushoff J, et al. Modeling shield immunity to reduce COVID-19 epidemic spread. Nature Medicine. 2020;26:849–854.

[14] Kissler SM, Tedijanto C, Goldstein E, Grad YH, Lipsitch M. Projecting the transmission dynamics of SARS-CoV-2 through the postpandemic period. Science. 2020;3(21):433. doi:10.1126/science.abb5793.

[15] Keeling MJ, Rohani P. Modeling infectious diseases in humans and animals. Princeton University Press; 2011.

[16] Coronavírus P. Dataset of cases of COVID-19 in Brazil; 2020. https://covid.saude.gov.br/.

[17] Io B. Dataset of cases of COVID-19 in Brazil; 2020. https://brasil.io/datasets/.

[18] Candido DS, Claro IM, de Jesus JG, Souza WM, Moreira FRR, Dellicou S, et al. Evolution and epidemic spread of SARS-CoV-2 in Brazil. Science;369:1255–1260.

[19] Candido DDS, Watts A, Abade L, Kraemer MU, Pybus OG, Croda J, et al. Routes for COVID-19 importation in Brazil. Journal of Travel Medicine. 2020;27(3):taaa042.

[20] Bolsonaro JM. Federal level decrees to overcome COVID-19 pandemic; 2020. https://www.planalto.gov.br/ccivil_03/_ato2019-2022/2020/decreto/d10282.htm.

[21] System BJ; 2020. http://www.stf.jus.br/arquivo/cms/noticiaNoticiaStf/anexo/ADPF672liminar.pdf.

[22] InLoco; 2020. https://mapabrasileirodacovid.inloco.com.br/pt/.

[23] Peixoto PS, Marcondes D, Peixoto C, Oliva SM. Modeling future spread of infections via mobile geolocation data and population dynamics. An application to COVID-19 in Brazil. PLOS One. 2020;15(7):e0235732.

[24] Ajzenman N, Cavalcanti T, Da Mata D. More than words: Leaders’ speech and risky behavior during a pandemic; 2020.

[25] Thomas Hale APTP Sam Webster, Kira B. Oxford COVID-19 Government Response Tracker. 2020;.

[26] Lin Q, Zhao S, Gao D, Lou Y, Yang S, Musa SS, et al. A conceptual model for the coronavirus disease 2019 (COVID-19) outbreak in Wuhan, China with individual reaction and governmental action. International journal of infectious diseases. 2020;93:211–216.

[27] Chowell G, Hengartner NW, Castillo-Chavez C, Fenimore PW, Hyman JM. The basic reproductive number of Ebola and the effects of public health measures: the cases of Congo and Uganda. Journal of theoretical biology. 2004;229(1):119–126.

[28] Dehning J, Zierenberg J, Spitzner FP, Wibral M, Neto JP, Wilczek M, et al. Inferring change points in the spread of COVID-19 reveals the effectiveness of interventions. Science. 2020;369(6500).

[29] Kissler SM, Tedijanto C, Goldstein E, Grad YH, Lipsitch M. Projecting the transmission dynamics of SARS-CoV-2 through the postpandemic period. Science. 2020;.

[30] Ferretti L, Wymant C, Kendall M, Zhao L, Nurtay A, Abeler-Döorner L, et al. Quantifying SARS-CoV-2 transmission suggests epidemic control with digital contact tracing. Science. 2020;368(6491).

[31] Mizumoto K, Kagaya K, Zarebski A, Chowell G. Estimating the asymptomatic proportion of coronavirus disease 2019 (COVID-19) cases on board the Diamond Princess cruise ship, Yokohama, Japan, 2020. Eurosurveillance. 2020;25(10):2000180.

[32] Cheng HY, Jian SW, Liu DP, Ng TC, Huang WT, Lin HH. Contact tracing assessment of COVID-19 transmission dynamics in Taiwan and risk at different exposure periods before and after symptom onset. JAMA Internal Medicine. 2020;180:1156–1163.

[33] Nishiura H, Kobayashi T, Miyama T, Suzuki A, Jung Sm, Hayashi K, et al. Estimation of the asymptomatic ratio of novel coronavirus infections (COVID-19). International Journal of Infectious Diseases. 2020;94:154.

[34] Sanche S, Lin YT, Xu C, Romero-Severson E, Hengartner N, Ke R. High Contagiousness and Rapid Spread of Severe Acute Respiratory Syndrome Coronavirus 2. Emerging Infectious Diseases. 2020;26(7).

[35] Li Q, Guan X, Wu P, Wang X, Zhou L, Tong Y, et al. Early transmission dynamics in Wuhan, China, of novel coronavirus–infected pneumonia. New England Journal of Medicine. 2020;382:1199–1207.

[36] Linton NM, Kobayashi T, Yang Y, Hayashi K, Akhmetzhanov AR, Jung Sm, et al. Incubation period and other epidemiological characteristics of 2019 novel coronavirus infections with right truncation: a statistical analysis of publicly available case data. Journal of clinical medicine. 2020;9(2):538.

[37] Kissler SM, Tedijanto C, Goldstein E, Grad YH, Lipsitch M. Projecting the transmission dynamics of SARS-CoV-2 through the postpandemic period. Science. 2020;368(6493):860–868.

[38] Miranda L. PySwarms: a research toolkit for particle swarm optimization in Python. Journal of Open Source Software. 2018;3(21):433.

[39] Flaxman S, Mishra S, Gandy A, Unwin HJT, Mellan TA, Coupland H, et al. Estimating the effects of non-pharmaceutical interventions on COVID-19 in Europe. Nature. 2020;584(7820):257–261.

[40] Wallinga J, Lipsitch M. How generation intervals shape the relationship between growth rates and reproductive numbers. Proceedings of the Royal Society B: Biological Sciences. 2007;274(1609):599–604.

[41] Van den Driessche P, Watmough J. Reproduction numbers and subthreshold endemic equilibria for compartmental models of disease transmission. Mathematical biosciences. 2002;180(1-2):29–48.

[42] Fraser C. Estimating individual and household reproduction numbers in an emerging epidemic. PloS one. 2007;2(8):e758.

[43] Jorge DC, Rodrigues MS, Silva MS, Cardim LL, da Silva NB, Silveira IH, et al.. Assessing the nationwide impact of COVID-19 mitigation policies on the transmission rate of SARS-CoV-2 in Brazil; 2020. https://github.com/Julian-sun/Assessing-the-nationwide-impact-of-COVID-19-mitigation-policies-on-the-transmgit.

[44] Bittar W. Paraná já registra casos de transmissão comunitária do novo coronavírus; 2020. https://cbncuritiba.com/parana-ja-registra-casos-de-transmissao-comunitaria-do-novo-coronavirus/.

[45] Araújo R. Roraima tem transmissão comunitária de coronavírus, diz secretária-adjunta de Saúde; 2020. https://g1.globo.com/rr/roraima/noticia/2020/05/02/roraima-tem-transmissao-comunitaria-de-coronavirus-diz-secretaria-adjunta-de-saude.ghtml.

[46] PAULO FDS. Maioria dos estados já tem mais de 70% de ocupação de UTI para Covid-19; 2020. https://www1.folha.uol.com.br/cotidiano/2020/05/maioria-dos-estados-ja-tem-mais-de-70-de-ocupacao-de-uti-para-covid-19.shtml.

[47] Tobías A. Evaluation of the lockdowns for the SARS-CoV-2 epidemic in Italy and Spain after one month follow up. Science of the Total Environment. 2020; p. 138539.

[48] Alfano V, Ercolano S. The Efficacy of Lockdown Against COVID-19: A Cross-Country Panel Analysis. Applied Health Economics and Health Policy. 2020; p. 1.

[49] Travassos C, Oliveira EXG, Viacava F. Desigualdades geográficas e sociais no acesso aos serviços de saúde no Brasil: 1998 e 2003. Ciência Saúde Coletiva. 2006;11:975 – 986.

[50] Garcia-Subirats I, Vargas I, Mogollón-Pérez AS, De Paepe P, Da Silva Mrf, Unger JP, et al. Inequities in access to health care in different health systems: a study in municipalities of central Colombia and north-eastern Brazil. International Journal for Equity in Health. 2014;13(1):10.

[51] Costa G, Cota W, Ferreira SC. Outbreak diversity in epidemic waves propagating through distinct geographical scales. Physical Review Research. 2020;2:043306.

[52] Jesus JGd, Sacchi C, Candido DdS, Claro IM, Sales FCS, Manuli ER, et al. Importation and early local transmission of COVID-19 in Brazil, 2020. Revista do Instituto de Medicina Tropical de São Paulo. 2020;62.

[53] Xavier J, Giovanetti M, Adelino T, Fonseca V, Costa AVBd, Ribeiro AA, et al. The ongoing COVID-19 epidemic in Minas Gerais, Brazil: insights from epidemiological data and SARS-CoV-2 whole genome sequencing. Emerging Microbes Infections. 2020;9(1):1824–1834.

[54] Forster P, Forster L, Renfrew C, Forster M. Phylogenetic network analysis of SARS-CoV-2 genomes. Proceedings of the National Academy of Sciences. 2020;117(17):9241–9243.

[55] Lancet T. COVID-19 in Brazil:”So what?”. Lancet (London, England). 2020;395(10235):1461.

[56] Rothgerber H, Wilson T, Whaley D, Rosenfeld DL, Humphrey M, Moore A, et al. Politicizing the Covid-19 pandemic: Ideological differences in adherence to social distancing; 2020. Available from: doi10.31234/ osf.io/k23cv.

[57] Pataro IM, Oliveira JF, Morato MM, Amad AA, Ramos PI, Pereira FA, et al. A control framework to optimize public health policies in the course of the COVID-19 pandemic. medRxiv. 2021;.

[58] He X, Lau EH, Wu P, Deng X, Wang J, Hao X, et al. Temporal dynamics in viral shedding and transmissibility of COVID-19. Nature Medicine. 2020;26(5):672–675.

[59] Malaquias RF, Silva AF. Understanding the use of mobile banking in rural areas of Brazil. Technology in Society. 2020; p. 101260.

[60] Pew Research Center. Smartphone Ownership Is Growing Rapidly Around the World, but Not Always Equally; 2019.

[61] Valle L. Transmissão comunitária de coronavírus no Acre já é uma realidade e proteção a grupos de riscos deve ser reforçada, alerta médico; 2020. https://agencia.ac.gov.br/transmissao-comunitaria-de-coronavirus-no-acre-ja-e-uma-realidade-e-protecao-a-grupos-de-riscos-deve-ser-reforcada-alerta-medico/.

[62] de Estado de Saúde do Amazonas S. Amazonas registra 111 casos do novo coronavírus e passa a ter transmissão comunitária; 2020. http://www.saude.am.gov.br/visualizar-noticia.php?id=4381.

[63] Coutinho C. Amapá confirma 1ª transmissão local de coronavírus; pacientes não têm sintomas graves; 2020. https://g1.globo.com/ap/amapa/noticia/2020/03/28/amapa-confirma-1a-transmissao-local-de-coronavirus-pacientes-nao-tem-sintomas-graves.ghtml.

[64] Lira M. Pará já tem transmissão comunitária do novo Coronavírus e 26 casos confirmados; 2020. http://www.saude.pa.gov.br/para-ja-tem-transmissao-comunitaria-do-novo-coronavirus-e-26-casos-confirmados/.

[65] Redação. Morte de idosa é o primeiro caso de trans-missão comunitária do Coronavírus em Porto Velho; 2020. https://www.rondoniagora.com/geral/morte-de-idosa-e-o-primeiro-caso-de-transmissao-comunitaria-do-coronavirus-em-porto-velho.

[66] de Saúde de Palmas-Tocantins SM; 2020. https://coronavirus.palmas.to.gov.br/storage/reports/LUbPAx3BlBu5Jz0FPysEvGKvG3BevrpnalDmBgfo.pdf.

[67] de Saúde de Alagoas S. Informe Epidemiológico; 2020. https://www.saude.al.gov.br/wp-content/uploads/2020/07/Informe-COVID-19-n°-41-16-DE-ABRIL-14h49.pdf.pdf.

[68] Brandão J. Bahia tem primeiros casos de transmissão comunitária de coronavírus; 2020. https://www.metro1.com.br/noticias/bahia/89207,bahia-tem-primeiros-casos-de-transmissao-comunitaria-de-coronavirus.

[69] do Estado do Ceará G. Nota Técnica n 1; 2020. https://www.saude.ce.gov.br/wp-content/uploads/sites/9/2020/02/NOTA_TECNICA_COVID19_21_03_20.pdf.

[70] Rafaelle Fróes AB. Maranhão tem casos de transmissão comunitária por coronavírus, diz secretário de Saúde; 2020. https://g1.globo.com/ma/maranhao/noticia/2020/03/30/maranhao-tem-casos-de-transmissao-comunitaria-por-coronavirus-diz-secretario-de-saude.ghtml.

[71] do Estado da Paraíba G. Governo reforça necessidade de no-tificação imediata de casos suspeitos Covid-19 pelos municípios; 2020. https://paraiba.pb.gov.br/noticias/governo-reforca-necessidade-de-notificacao-imediata-de-casos-suspeitos-covid-19-pelos-municipios.

[72] Amanda Azevedo CL. Pernambuco tem primeiro caso de transmissão comunitária de coronavírus; 2020. https://jc.ne10.uol.com.br/colunas/saude-e-bem-estar/2020/03/5602589-pernambuco-tem-primeiro-caso-de-transmissao-comunitaria-de-coronavirus.html.

[73] de Estado da Saúde Sergipe S. Sergipe tem mais um caso confirmado de coronavírus; 2020. https://www.saude.se.gov.br/sergipe-tem-mais-um-caso-confirmado-de-coronavirus/.

[74] de Estado da Saúde Goiás S. Governo de Goiás destaca que união de esforços pode conter novo coronavírus; 2020. https://www.saude.go.gov.br/coronavirus/noticias-coronavirus/10578-governo-de-goias-destaca-que-uniao-de-esforcos-pode-conter-novo-coronavirus.

[75] de Saúde do Distrito Federal S. Boletim Epidemiológico; 2020.http://www.saude.df.gov.br/wp-conteudo/uploads/2020/03/26-Boletim-COVID_DF-26.03.2020_2vers∼ao-1.pdf.

[76] MT G. Governo reconhece que há transmissão comunitária de coronavírus em três municípios de MT; 2020. https://g1.globo.com/mt/mato-grosso/noticia/2020/04/01/governo-reconhece-que-ha-transmissao-comunitaria-de-coronavirus-em-tres-municipios-de-mt.ghtml.

[77] Ângela Kempfer. Com transmissão comunitária, MS já tem 113 casos e 4 mortes por coronavírus; 2020. https://www.campograndenews.com.br/cidades/capital/com-transmissao-comunitaria-ms-ja-tem-113-casos-e-4-mortes-por-coronavirus.

[78] do Estado do Espírito Santo G. NOTA TÉCNICA COVID-19 N° 04/2020 – GEVS/SESA/ES; 2020. https://coronavirus.es.gov.br/Media/Coronavirus/NotasTecnicas.

[79] do Estado de Minas Gerais G. BOLETIM INFORMATIVO DIÁRIO; 2020. https://coronavirus.saude.mg.gov.br/images/boletim/03-marco/18032020_Boletim_epidemiologico_COVID-19_MG.pdf.

[80] Amorim F. Brasil confirma transmissão comunitária de coronavírus; entenda o que é; 2020. https://noticias.uol.com.br/saude/ultimas-noticias/redacao/2020/03/13/brasil-confirma-transmissao-comunitaria-de-coronavirus-entenda-o-que-e.htm.

[81] Secom R. RS confirma transmissão comunitária do novo coro-navírus; 2020. Available from: https://estado.rs.gov.br/rs-confirma-transmissao-comunitaria-do-novo-coronavirus.

[82] do Estado de Saúde Santa Catarina S. CORO-NAVíRUS EM SC: GOVERNO CONFIRMA 14 CA-SOS E TRANSMISSÕ ES COMUNITÁRIAS; 2020. https://www.saude.sc.gov.br/coronavirus/CORONAVIRUS-EM-SC-GOVERNO-CONFIRMA-14-CASOS-E-TRANSMISSOES-COMUNITARIAS.html.

[83] Sobol IM. Global sensitivity indices for nonlinear mathematical models and their Monte–Carlo estimates. Mathematics and Computers in Simulation. 2001;55(1-3):271–280.

[84] Saltelli A, Ratto M, Andres T, Campolongo F, Cariboni J, Gatelli D, et al. Global Sensitivity Analysis: The Primer. John Wiley & Sons; 2008.

[85] Herman J, Usher W. SALib: An open-source Python library for Sensitivity Analysis. The Journal of Open Source Software. 2017;2(9). doi:10.21105/joss.00097.

