## Supplementary material for "Assessing the nationwide impact of COVID-19 mitigation policies on the transmission rate of SARS-CoV-2 in Brazil": Code analysis.ipynb

```
# %% [code]
import numpy as np
import matplotlib.pyplot as plt
from scipy.integrate import odeint
from scipy.optimize import least_squares
import math
import pandas as pd
import csv
import datetime
from scipy import stats
from scipy import optimize
from datetime import date, timedelta
import random
import requests
from scipy.stats import truncnorm
import datetime

import statsmodels as sm
#import statsmodels.api as sm

from PIL import Image
from scipy.stats import norm

from statsmodels.tsa.vector_ar.var_model import VAR

import functools

# %% [markdown]
# Read data

# %% [markdown]
## Cases

# %% [code]
df = pd.read_csv('/Users/julianeoliveira/Desktop/dropbox_15_11_2019/Artigos_preprints/COVID19/Paper_submissions/Paper2/Assessing-the-nationwide-impact-of-COVID-19-mitigation-policies-on-the-transmission-rate-of-SARS-CoV/data/covid.csv')

# %% [markdown]
## Population

# %% [code]
pop = pd.read_csv('/Users/julianeoliveira/Desktop/dropbox_15_11_2019/Artigos_preprints/COVID19/Paper_submissions/Paper2/Assessing-the-nationwide-impact-of-COVID-19-mitigation-policies-on-the-transmission-rate-of-SARS-CoV/data/populacao_municipio.csv')

# %% [markdown]
## Mobility data

# %% [code]
mob = pd.read_csv('/Users/julianeoliveira/Desktop/dropbox_15_11_2019/Artigos_preprints/COVID19/Paper_submissions/Paper2/Assessing-the-nationwide-impact-of-COVID-19-mitigation-policies-on-the-transmission-rate-of-SARS-CoV/data/distancing_series.csv')

# %% [markdown]
## Stringency

# %% [code]
strg = pd.read_csv('/Users/julianeoliveira/Desktop/dropbox_15_11_2019/Artigos_preprints/COVID19/Paper_submissions/Paper2/Assessing-the-nationwide-impact-of-COVID-19-mitigation-policies-on-the-transmission-rate-of-SARS-CoV/data/stringency_series.csv',sep= ',')

# %% [code]
strg

# %% [markdown]
# Create dataframe to analyse the following time series:

    * Daily number of cases
    * Acumulated number of cases
    * SMRI
    * Stringency
    * Rt

# %% [code]
df = df[(df.date >= '2020-03-01') & (df.date <= '2020-05-22')]

# %% [code]
mob['date2'] = pd.to_datetime(mob.date)

# %% [code]
mob.head(2)

# %% [code]
mob = mob[(mob.date2 <= '2020-05-22')]

# %% [code]
series_state = df.groupby(['state', 'date'])['new_confirmed'].sum().reset_index()

# %% [code]
pop.head()

# %% [code]
capital_dic={
    'AC': 'Rio Branco',
    'AL': 'Maceió',
    'AP': 'Macapá',
    'AM': 'Manaus',
    'BA': 'Salvador',
    'CE': 'Fortaleza',
    'DF': 'Brasília',
    'ES': 'Vitória',
    'GO': 'Goiânia',
    'MA': 'São Luís',
    'MT': 'Cuiabá',
    'MS': 'Campo Grande',
    'MG': 'Belo Horizonte',
    'PA': 'Belém',
    'PB': 'João Pessoa',
    'PR': 'Curitiba',
    'PE': 'Recife',
    'PI': 'Teresina',
    'RJ': 'Rio de Janeiro',
    'RN': 'Natal',
    'RS': 'Porto Alegre',
    'RO': 'Porto Velho',
    'RR': 'Boa Vista',
    'SC': 'Florianópolis',
    'SP': 'São Paulo',
    'SE': 'Aracaju',
    'TO': 'Palmas'
}

pop_dic={}

for state in pop['ARmaior'].unique():
    pop_state=[]

    Sigla=pop.loc[pop['ARmaior'] == state].NomeMunic.values[0][-2]+pop.loc[pop['ARmaior'] == state].NomeMunic.values[0][-1]
    
    pop_state.append(sum(pop.loc[pop['ARmaior'] == state].Total.values))
    
    Sigla=pop.loc[pop['ARmaior'] == state].NomeMunic.values[0][-2]+pop.loc[pop['ARmaior'] == state].NomeMunic.values[0][-1]
    
    ind1=list(pop.loc[(pop['ARmaior'] == state)& (pop['Sexo'] =='f' )].NomeMunic.values).index(capital_dic[Sigla]+" - "+Sigla)
    ind2=list(pop.loc[(pop['ARmaior'] == state)& (pop['Sexo'] =='m' )].NomeMunic.values).index(capital_dic[Sigla]+" - "+Sigla)

    
    
    pop_state.append(pop.loc[(pop['ARmaior'] == state)& (pop['Sexo'] =='f' )].Total.values[ind1]+pop.loc[(pop['ARmaior'] == state)& (pop['Sexo'] =='m' )].Total.values[ind2])
    
    
    pop_state.append(pop_state[0]-pop_state[1])
    
    pop_dic[Sigla]=pop_state

# %% [code]
#conv_list={'estado':0, 'capital':1, 'interior':2}

np.round(pop_dic['TO'],decimals=0)

# %% [markdown]
# Descritive analysis of the incidence in the regions of study

# %% [code]
names = ['Region','Type','Date','cum_cases','Pop','New']

dta = pd.read_excel('/Users/julianeoliveira/Desktop/dropbox_15_11_2019/Artigos_preprints/COVID19/Paper_submissions/Paper2/Assessing-the-nationwide-impact-of-COVID-19-mitigation-policies-on-the-transmission-rate-of-SARS-CoV/data/allcases.xlsx',header=None,names=names)

dta =dta.drop([0, 2])

# %% [code]
dta['state'] = dta.Region.str[-2:]

# %% [code]
dta = dta.drop([1])

# %% [code]
dta

# %% [code]
dta['inc'] = dta['New']*100000/dta['Pop']
dta

# %% [code]
#dado = dta[(dta.state =='AC') | (dta.state =='RO') | (dta.state =='AM') | (dta.state =='RR') | (dta.state =='AP') | (dta.state =='PA') | (dta.state =='TO')]

dado = dta[(dta.state =='SE') | (dta.state =='RN') | (dta.state =='AL') | (dta.state =='PI') | (dta.state =='BA') | (dta.state =='CE') | (dta.state =='PE') | (dta.state =='PB') | (dta.state =='MA')]

# %% [code]
dado.head()

# %% [code]
dado.Type.unique()

# %% [code]
dado = dado[dado.Type != 'Estado']

# %% [code]
dado.head()

# %% [code]
dado =dado.pivot(index='Region', columns='Date', values='inc').fillna(0)

# %% [code]
import seaborn as sns
sns.set_theme()

sns.set(font_scale=42)

sns.set(rc={'figure.figsize':(20,10)})

sns_plot = sns.heatmap(round(dado,0), cmap="Reds",annot=True)


plt.tight_layout()

#sns_plot.figure.savefig("inc_capi_int.png")

# %% [code]
dado2 = dta[dta.Type == 'Estado']

# %% [code]
dado2 =dado2.pivot(index='Region', columns='Date', values='inc').fillna(0)

# %% [code]
import seaborn as sns
sns.set_theme()

sns.set(font_scale=42)

sns.set(rc={'figure.figsize':(20,10)})

sns_plot = sns.heatmap(round(dado2,0), cmap="Reds",annot=True)


plt.tight_layout()

#sns_plot.figure.savefig("inc_capi.png")

# %% [markdown]
# Stringency descriptive

# %% [code]
strg = pd.read_csv('/Users/julianeoliveira/Desktop/dropbox_15_11_2019/Artigos_preprints/COVID19/Paper_submissions/Paper2/Assessing-the-nationwide-impact-of-COVID-19-mitigation-policies-on-the-transmission-rate-of-SARS-CoV/data/stringency_series.csv',sep= ',')

strg['date'] = pd.to_datetime(strg.Date)

#strg = strg[(strg.date >= '2020-03-11') & (strg.date <= '2020-03-31')]

# %% [code]
strg

# %% [code]
strg['indGeneral'] = strg['indGeneral']/100

# %% [code]
for value in strg.UF.unique():
    print(value, round(strg[strg.UF == value].describe(),2))

# %% [markdown]
# Correlation mobility and cases

# %% [code]
names = ['Region','Type','Date','cum_cases','Pop','New']

dta = pd.read_excel('/Users/julianeoliveira/Desktop/dropbox_15_11_2019/Artigos_preprints/COVID19/Paper_submissions/Paper2/Assessing-the-nationwide-impact-of-COVID-19-mitigation-policies-on-the-transmission-rate-of-SARS-CoV/data/allcases.xlsx',header=None,names=names)

dta =dta.drop([0, 1])

dta.head()

# %% [code]
dta['state'] = dta.Region.str[0:2]

# %% [code]
dta.info()

# %% [code]
state = dta[dta.state== 'AL']

capState = state[state.Type == 'Interior']

# %% [code]
capState['new_SMA_3'] = capState.New.astype(int).rolling(window=3).mean()
capState['new_SMA_7'] = capState.New.astype(int).rolling(window=7).mean()

# %% [code]
capState['GR'] = np.log(round(capState['new_SMA_3'],3))/np.log(round(capState['new_SMA_7'],3))
#GR = GR.fillna(0).replace(np.inf, 0, inplace=True)

capState['GR'].replace(np.inf, 0, inplace=True)

capState['GR'][capState['GR'] < 0] = 0

capState['GR'] = capState['GR'].fillna(0)

# %% [code]
lst = []
for i in range(0,len(capState['New'])):
    c = (capState['New'].iloc[i]/capState['new_SMA_3'].iloc[i] ) if capState['new_SMA_3'].iloc[i] != 0 else 0
    lst.append(c)
    
capState['GR2'] = lst


capState['GR2'].replace(np.inf, 0, inplace=True)

#capState['GR2'][capState['GR2'] < 0] = 0

capState['GR2'] = capState['GR2'].fillna(0)

# %% [code]
capState.head(3)

# %% [code]
mob.head(2)

# %% [code]
mobs = mob[mob.name == 'AL'][(mob[mob.name == 'AL'].date2 >= capState.iloc[0,2])]
mobs

mobs['indInland'];

# %% [code]
def crosscorr(datax, datay, lag=0):
    """ Lag-N cross correlation. 
    Parameters
    ----------
    lag : int, default 0
    datax, datay : pandas.Series objects of equal length

    Returns
    ----------
    crosscorr : float
    """
    return datax.corr(datay.shift(lag))

# %% [code]
new = pd.DataFrame([capState['GR'],capState['GR2']]).transpose().reset_index()

new['indInland'] = mobs['indInland'].values
new = new.filter(['GR','GR2','indInland'])

# %% [code]
new.head()

# %% [code]
xcov = [crosscorr(new['indInland'], new['GR2'], lag=i) for i in range(40)]
print(xcov)

# %% [code]
plt.figure(figsize=(8,3))

plt.grid()
plt.plot(xcov, 'o')

n =len(xcov)
l1 = (-1/n) + (2/math.sqrt(n))

plt.axhline(l1, color='blue', linestyle='dashed', lw=1)

plt.axhline(-l1, color='blue', linestyle='dashed', lw=1)


# %% [code]


# %% [code]
x = mobs[s1]
y = capState[s2]


fig, ax1 = plt.subplots(1, 1, sharex=True)
ax1.xcorr(x, y, usevlines=True, maxlags=50, normed=True, lw=2)
ax1.grid(True)

#ax2.acorr(x, usevlines=True, normed=True, maxlags=50, lw=2)
#ax2.grid(True)

#ax3.acorr(y, usevlines=True, normed=True, maxlags=50, lw=2)
#ax3.grid(True)

plt.show()

# %% [code]
mobs.filter([s1]).apply(functools.partial(test_stationarity))

# %% [code]
capState.filter([s2]).apply(functools.partial(test_stationarity))

# %% [code]
mobs[s1]

# %% [code]
new = pd.DataFrame([capState['GR']]).transpose().reset_index()

new[s1] = mobs[s1].values
new = new.filter(['GR',s1])

# %% [code]
new.head()

# %% [code]
model = VAR(new)
    
model1 = model.select_order()
print(model1.summary())

# %% [code]
xcov = [crosscorr(new[s1], new[s2], lag=i) for i in range(40)]
print(xcov)

# %% [code]
plt.figure(figsize=(8,3))

plt.grid()
plt.plot(xcov, 'o')

n =len(xcov)
l1 = (-1/n) + (2/math.sqrt(n))

plt.axhline(l1, color='blue', linestyle='dashed', lw=1)

plt.axhline(-l1, color='blue', linestyle='dashed', lw=1)


# %% [code]
# Differencing to get the data stationary
    
casesDiff = cases.filter([s1,s2]).diff().dropna()
    
casesDiff = casesDiff.apply(pd.to_numeric)
    
    print('\x1b[0;30;43m'+ 'stationarity of the series {}'.format(s1) + ' and {}'.format(s2) +'\x1b[0m')
    
    cases.filter([s1,s2]).apply(functools.partial(test_stationarity))
    
    print('\x1b[0;30;43m'+ 'stationarity of the differentiated series {}'.format(s1) + ' and {}'.format(s2)+'\x1b[0m')
    
    casesDiff.apply(functools.partial(test_stationarity))

# %% [code]
lst= []
for s in mob.name.unique():
    
    print(s)
    
    #filtering the state in the datasets
    #cases = series_state[series_state.state == s]
    state = dta[dta.state== 'AL']
    capState = state[state.Type == 'Estado']
    
    mobs = mob[mob.name == s][(mob[mob.name == s].date2 >= state.iloc[0,2])]
    
    #strg = strg[strg.UF == s]
    
    # creating series of cummulative, log and etc
    
    capState['indUF'] = np.array(mobs.indUF)    
    
    lst.append(state)

# %% [code]
data_corr = pd.concat(lst,sort=True)
data_corr.head()

# %% [code]
#data_corr.to_csv('data_corr.csv')

# %% [code]
lst

data_corr = pd.concat(lst,sort=True)
data_corr.head()

#data_corr.to_csv('data_corr.csv')

# %% [code]
plt.figure(figsize=(20,10))
plt.plot(capState['GR'])
#plt.plot(mob[mob.name == 'AL'][s1]/100)

# %% [code]
# croscorrelation with diferent lags 
    
s1 = 'indUF'
s2 = 'GR'
    
print('\x1b[0;30;43m'+ 'Crosscorrelation with lags between the series {}'.format(s1) + ' and {}'.format(s2) +'\x1b[0m')
    
xcov = [crosscorr(mob[mob.name == 'AL'][s1], capState[s2], lag=i) for i in range(40)]
print(xcov)

# %% [code]
plt.figure(figsize=(10,8))

plt.grid()
plt.plot(xcov, 'o')

n =len(xcov)
l1 = (-1/n) + (2/math.sqrt(n))

    #fig, [ax1,ax2] = plt.subplots(3, 1)
    #ax1.xcorr(x, y, usevlines=True, maxlags=None, normed=True, lw=2)

plt.axhline(l1, color='blue', linestyle='dashed', lw=1)

    #ax1.grid(True)

    #ax2.acorr(x, usevlines=True, normed=True, maxlags=None, lw=2)
    #ax2.grid(True)
    
    #ax2.scatter(cases.new_confirmedLog,cases.indUF, marker='o')
    #ax2.legend('new_confirmedLog','indUF')
    
    #ax3.plot(xcov, marker='o')
    #ax3.axhline(l1, color='blue', linestyle='dashed', lw=1)
plt.axhline(-l1, color='blue', linestyle='dashed', lw=1)
    #ax3.axis(ymin=-1,ymax=1)

    #plt.show()

# %% [code]
for s in mob.name.unique():
    
    print(s)
    
    #filtering the state in the datasets
    state = dta[dta.state== s]
    
    onlyState = state[state.Type == 'Estado']
    capState = state[state.Type == 'Capital']
    intState = state[state.Type == 'Interior']
    
    
    mobs = mob[mob.name == s][(mob[mob.name == s].date2 >= onlyState.iloc[0,2])]
    
    #mobUF = mob[mob.name == s]['indUF']
    #mobUF = mob[mob.name == s]['indCAP']
    #mobUF = mob[mob.name == s]['indInland']
    
    
    # croscorrelation with diferent lags 
    
    s1 = 'indUF'
    s2 = 'GR'
    
    print('\x1b[0;30;43m'+ 'Crosscorrelation with lags between the series {}'.format(s1) + ' and {}'.format(s2) +'\x1b[0m')
    
    xcov = [crosscorr(mobs[s1], onlyState[s2], lag=i) for i in range(40)]
    print(xcov)


# %% [code]
dta.head()

# %% [markdown]
# Functions

# %% [code]
from statsmodels.tsa.stattools import adfuller
def test_stationarity(timeseries):
    
    #Perform Dickey-Fuller test:
    print ('Results of Dickey-Fuller Test:')
    dftest = adfuller(timeseries, autolag='AIC')
    dfoutput = pd.Series(dftest[0:4], index=['Test Statistic','p-value','#Lags Used','Number of Observations Used'])
    for key,value in dftest[4].items():
        dfoutput['Critical Value (%s)'%key] = value
    print (dfoutput)

# %% [code]
def crosscorr(datax, datay, lag=0):
    """ Lag-N cross correlation. 
    Parameters
    ----------
    lag : int, default 0
    datax, datay : pandas.Series objects of equal length

    Returns
    ----------
    crosscorr : float
    """
    return datax.corr(datay.shift(lag))

# %% [code]
lst= []
for s in mob.name.unique():
    
    print(s)
    
    #filtering the state in the datasets
    cases = series_state[series_state.state == s]
    
    mobs = mob[mob.name == s][(mob[mob.name == s].date2 >= cases.iloc[0,1])]
    
    strg = strg[strg.UF == s]
    
    # creating series of cummulative, log and etc
    
    cases['indUF'] = np.array(mobs.indUF)    
    
    lst.append(cases)

# %% [code]
lst

# %% [code]
data_corr = pd.concat(lst,sort=True)
data_corr.head()

# %% [code]
#data_corr.to_csv('data_corr.csv')

# %% [code]
for s in mob.name.unique():
    
    print(s)
    
    #filtering the state in the datasets
    cases = series_state[series_state.state == s]
    
    mobs = mob[mob.name == s][(mob[mob.name == s].date2 >= cases.iloc[0,1])]
    
    # creating series of cummulative, log and etc
    
    cases['cumulative'] = cases.new_confirmed.cumsum()
    
    cases['cumLog'] = np.log(cases.filter(['cumulative']))
    
    cases['indUF'] = np.array(mobs.indUF)    
    
    cases['indUFLog'] = np.log(cases.filter(['indUF']))
    
    cases['new_confirmedLog'] = np.log(cases.filter(['new_confirmed']))
    
    cases = cases.replace([np.inf, -np.inf], np.nan).fillna(0)
    
    # test stationarity
    
    s1 = 'indUF'
    s2 = 'new_confirmed'
    
    # Differencing to get the data stationary
    
    casesDiff = cases.filter([s1,s2]).diff().dropna()
    
    casesDiff = casesDiff.apply(pd.to_numeric)
    
    print('\x1b[0;30;43m'+ 'stationarity of the series {}'.format(s1) + ' and {}'.format(s2) +'\x1b[0m')
    
    cases.filter([s1,s2]).apply(functools.partial(test_stationarity))
    
    print('\x1b[0;30;43m'+ 'stationarity of the differentiated series {}'.format(s1) + ' and {}'.format(s2)+'\x1b[0m')
    
    casesDiff.apply(functools.partial(test_stationarity))
    

# %% [code]
for s in mob.name.unique():
    
    print(s)
    
    #filtering the state in the datasets
    cases = series_state[series_state.state == s]
    
    mobs = mob[mob.name == s][(mob[mob.name == s].date2 >= cases.iloc[0,1])]
    
    # creating series of cummulative, log and etc
    
    cases['cumulative'] = cases.new_confirmed.cumsum()
    
    cases['cumLog'] = np.log(cases.filter(['cumulative']))
    
    cases['indUF'] = np.array(mobs.indUF)    
    
    cases['indUFLog'] = np.log(cases.filter(['indUF']))
    
    cases['new_confirmedLog'] = np.log(cases.filter(['new_confirmed']))
    
    cases = cases.replace([np.inf, -np.inf], np.nan).fillna(0)
    
    
    # Analyse the lag dependence using Var
    
    s1 = 'indUF'
    s2 = 'new_confirmed'
    
    # Differencing to get the data stationary
    
    casesDiff = cases.filter([s1,s2]).diff().dropna()
    
    casesDiff = casesDiff.apply(pd.to_numeric)
    
    print('\x1b[0;30;43m'+ 'Var of the series {}'.format(s1) + ' and {}'.format(s2) +'\x1b[0m')

    model = VAR(cases.filter([s1,s2]))
    
    model1 = model.select_order()
    print(model1.summary())
    

# %% [code]
for s in mob.name.unique():
    
    print(s)
    
    #filtering the state in the datasets
    cases = series_state[series_state.state == s]
    
    mobs = mob[mob.name == s][(mob[mob.name == s].date2 >= cases.iloc[0,1])]
    
    # creating series of cummulative, log and etc
    
    cases['cumulative'] = cases.new_confirmed.cumsum()
    
    cases['cumLog'] = np.log(cases.filter(['cumulative']))
    
    cases['indUF'] = np.array(mobs.indUF)    
    
    cases['indUFLog'] = np.log(cases.filter(['indUF']))
    
    cases['new_confirmedLog'] = np.log(cases.filter(['new_confirmed']))
    
    cases = cases.replace([np.inf, -np.inf], np.nan).fillna(0)
    
    
    # calculate correlation between series without lag and any previous analysis
    
    s1 = 'indUF'
    s2 = 'new_confirmedLog'
    
    print('\x1b[0;30;43m'+ 'Crosscorrelation between the series {}'.format(s1) + ' and {}'.format(s2) +'\x1b[0m')

    
    r, p = stats.pearsonr(cases.dropna()[s1], cases.dropna()[s2])
    print(f"Scipy computed Pearson r: {r} and p-value: {p}")
    
    r, p = stats.spearmanr(cases.dropna()[s1], cases.dropna()[s2])
    print(f"Scipy computed Spearman r: {r} and p-value: {p}")
    
    r, p = stats.kendalltau(cases.dropna()[s1], cases.dropna()[s2])
    print(f"Scipy computed Kendall's tau: {r} and p-value: {p}")

# %% [code]
for s in mob.name.unique():
    
    print(s)
    
    #filtering the state in the datasets
    cases = series_state[series_state.state == s]
    
    mobs = mob[mob.name == s][(mob[mob.name == s].date2 >= cases.iloc[0,1])]
    
    # creating series of cummulative, log and etc
    
    cases['cumulative'] = cases.new_confirmed.cumsum()
    
    cases['cumLog'] = np.log(cases.filter(['cumulative']))
    
    cases['indUF'] = np.array(mobs.indUF)    
    
    cases['indUFLog'] = np.log(cases.filter(['indUF']))
    
    cases['new_confirmedLog'] = np.log(cases.filter(['new_confirmed']))
    
    cases = cases.replace([np.inf, -np.inf], np.nan).fillna(0)
    
    
    # croscorrelation with diferent lags 
    
    s1 = 'indUF'
    s2 = 'new_confirmedLog'
    
    print('\x1b[0;30;43m'+ 'Crosscorrelation with lags between the series {}'.format(s1) + ' and {}'.format(s2) +'\x1b[0m')
    
    xcov = [crosscorr(cases[s1], cases[s2], lag=i) for i in range(40)]
    print(xcov)
    
    
    #x= cases.indUF
    #y= cases.new_confirmedLog

    #n =len(x)
    #l1 = (-1/n) + (2/math.sqrt(n))

    #fig, [ax1,ax2] = plt.subplots(3, 1)
    #ax1.xcorr(x, y, usevlines=True, maxlags=None, normed=True, lw=2)

    #ax1.axhline(l1, color='blue', linestyle='dashed', lw=1)

    #ax1.grid(True)

    #ax2.acorr(x, usevlines=True, normed=True, maxlags=None, lw=2)
    #ax2.grid(True)
    
    #ax2.scatter(cases.new_confirmedLog,cases.indUF, marker='o')
    #ax2.legend('new_confirmedLog','indUF')
    
    #ax3.plot(xcov, marker='o')
    #ax3.axhline(l1, color='blue', linestyle='dashed', lw=1)
    #ax3.axhline(-l1, color='blue', linestyle='dashed', lw=1)
    #ax3.axis(ymin=-1,ymax=1)

    #plt.show()

# %% [markdown]
# Analysis of the fit results

# %% [markdown]
## BIC

# %% [code]
bic = pd.read_csv('/Users/julianeoliveira/Desktop/seiir_fits_bic.csv')
bic

# %% [code]
bic[bic.state == 'AC']

# %% [code]
lst = []
for i in range(0, len(bic)-1):
    value =  bic['BIC'][i] - bic['BIC'][i+1]
    lst.append(value)
lst.append(0)


# %% [code]
bic['dif'] = lst

# %% [code]
bic.filter(['state','type','BIC','dif']).to_csv('/Users/julianeoliveira/Desktop/seiir_fits_bic2.csv')

# %% [markdown]
## Data results

# %% [code]
import sys
import glob

path = r'/Users/julianeoliveira/Desktop/dropbox_15_11_2019/Artigos_preprints/COVID19/Paper_submissions/Paper2/Assessing-the-nationwide-impact-of-COVID-19-mitigation-policies-on-the-transmission-rate-of-SARS-CoV/results/seiir_bootstrap'
filenames = glob.glob(path + "/*.csv")
filenames

# %% [code]
lst_df = []
for f in filenames:
    print(f)
    df = pd.read_csv(f, low_memory = False, encoding="iso-8859-1")     
    #df.columns= df.columns.str.lower()
    lst_df.append(df)

# %% [code]
df = pd.concat(lst_df,sort=True)
df.head()

# %% [markdown]
## Calculation of the confidence intervals

# %% [code]
#df = pd.read_csv('/Users/julianeoliveira/Downloads/seiir_fits_boots_SE_new.csv')

# %% [code]
import scipy.stats

def mean_confidence_interval(data, confidence=0.95):
    a = 1.0 * np.array(data)
    n = len(a)
    m, se = np.median(a), scipy.stats.sem(a)
    h = se * scipy.stats.skewnorm.ppf((1 + confidence) / 2., n-1)
    return m, m-h, m+h

def day_date(t):
    t_date = []
    for days in t:
        days = np.nan_to_num(days)
        date = datetime.datetime(2020, 1, 1) + datetime.timedelta(int(days) - 1)
        t_date.append(date)

    return(t_date)

def r0(b,delta,gammaA=1/3.5,gammaS=1/4,p=0.2):

    R0a = b*delta/gammaA

    R0s = b/gammaS

    R0_geral = p*R0s + (1-p)*R0a
    
    return(R0_geral)

# %% [code]
df.head()

# %% [code]
dfs = []
for uf in df.state.unique():
    for area in df.type.unique():
    
        frame = df[(df.state == uf) & (df.type == area)]


        beta0= np.array(mean_confidence_interval(frame['beta0'], confidence=0.95))

        beta0Round = np.round(mean_confidence_interval(frame['beta0'], confidence=0.95), decimals=2)

        beta1 = np.round(mean_confidence_interval(frame['beta1'], confidence=0.95), decimals=2)

        beta2 = np.round(mean_confidence_interval(frame['beta2'], confidence=0.95), decimals=2)

        delta = np.array(mean_confidence_interval(frame['delta'], confidence=0.95))

        deltaRound = np.round(mean_confidence_interval(frame['delta'], confidence=0.95), decimals=2)

        tcut0 = np.round(mean_confidence_interval(frame['tcut0'], confidence=0.95), decimals=0)

        tcut1 = np.round(mean_confidence_interval(frame['tcut1'], confidence=0.95), decimals=0)

        tcut0_date = np.array(day_date(tcut0))

        tcut1_date = np.array(day_date(tcut1))

        R0 = np.round(r0(beta0,delta),2)
    
        data = np.array([beta0, beta1,beta2,delta,tcut0,tcut0_date,tcut1,tcut1_date,R0])
        df1 = pd.DataFrame(data=data, index=['beta0','beta1','beta2','delta','tcut0','tcut0_date','tcut1','tcut1_date','R0'],columns=['median','lower','upper']).transpose()
        df1['state'] = uf
        df1['type'] = area
        dfs.append(df1)

# %% [code]
dfs

# %% [code]
dfs_result = pd.concat(dfs,sort=True)
dfs_result

# %% [code]
dfs_result['decrease'] = (dfs_result['beta1'] - dfs_result['beta0'])*100/dfs_result['beta0']

dfs_result

# %% [code]
dado = dfs_result[(dfs_result['type'] == 'capital')].reset_index()

dado[dado['index'] == 'median']['R0'].mean()

# %% [code]
dfs_result[(dfs_result['R0'] > 1.5) & (dfs_result['R0'] < 2)]

# %% [code]
#dfs_result.filter(['state','type','decrease']).to_csv('decrease.csv')

# %% [code]
#dfs_result.to_csv('/Users/julianeoliveira/Desktop/dropbox_15_11_2019/Artigos_preprints/COVID19/Paper_submissions/Paper2/Assessing-the-nationwide-impact-of-COVID-19-mitigation-policies-on-the-transmission-rate-of-SARS-CoV/results/ic_par.csv')

# %% [markdown]
# Comparing dates 

# %% [code]
pars = pd.read_csv('/Users/julianeoliveira/Desktop/dropbox_15_11_2019/Artigos_preprints/COVID19/Paper_submissions/Paper2/Assessing-the-nationwide-impact-of-COVID-19-mitigation-policies-on-the-transmission-rate-of-SARS-CoV/results/ic_par.csv')

# %% [code]
pars.head()

# %% [code]
pars.tcut0_date =  pd.to_datetime(pars.tcut0_date)

# %% [code]
series_state.head()

# %% [code]
series_state.date =  pd.to_datetime(series_state.date)

# %% [code]
for s in series_state.state.unique():

    #filtering the state in the datasets
    cases = series_state[series_state.state == s]
    
    p = pars[pars.state == s]
    
    mobs = mob[mob.name == s]
    mobs = mobs[(mobs[mobs.name == s].date2 >= cases.iloc[0,1])]
    
    mobs[mobs.indUF == max(mobs.indUF)]    
    
    d1 = cases.iloc[0,1]
    
    d2 = p.iloc[0,7]
    
    
    print(s, d1, d2, 'tcut1 and first case ->', abs((d2 - d1).days))
    
    #mobs = mob[mob.name == s][(mob[mob.name == s].date2 >= cases.iloc[0,1])]

# %% [code]
mob

# %% [code]
s = 'SC'
mobs = mob[mob.name == s][(mob[mob.name == s].date2 >= cases.iloc[0,1])]
    
mobs[mobs.indUF == max(mobs.indUF)]

# %% [code]
series_state[series_state.state == 'SC']

# %% [code]
pars[pars.state == 'SC']

# %% [markdown]
# ... Others


# %% [code]
sig_uf = []
for value in mob.state_name:
    if value == 'Minas Gerais': value = 'MG'
    elif value == 'Ceará': value = 'CE'
    elif value == 'Pernambuco': value = 'PE'
    elif value == 'Mato Grosso do Sul': value = 'MS'
    elif value == 'Rio Grande do Sul': value = 'RS'
    elif value == 'Sergipe': value = 'SE'
    elif value == 'Rio Grande do Norte': value = 'RN'
    elif value == 'Acre': value = 'AC'
    elif value == 'Alagoas': value = 'AL'
    elif value == 'Bahia': value = 'BA'
    elif value == 'Rondônia': value = 'RO'
    elif value == 'Rio de Janeiro': value = 'RJ'
    elif value == 'Piauí': value = 'PI'
    elif value == 'Paraná': value = 'PR'
    elif value == 'Espírito Santo': value = 'ES'
    elif value == 'Pará': value = 'PA'
    elif value == 'Amapá': value = 'AP'
    elif value == 'Mato Grosso': value = 'MT'
    elif value == 'Paraíba': value = 'PB'
    elif value == 'Roraima': value = 'RR'
    elif value == 'Goiás': value = 'GO'
    elif value == 'São Paulo': value = 'SP'
    elif value == 'Distrito Federal': value = 'DF'
    elif value == 'Maranhão': value = 'MA'
    elif value == 'Santa Catarina': value = 'SC'
    elif value == 'Tocantins': value = 'TO'
    elif value == 'Amazonas': value = 'AM'
    sig_uf.extend([value])

# %% [code]


import seaborn as sns
import matplotlib.pyplot as plt
sns.set_theme(style="ticks", color_codes=True)

titanic = pd.read_csv('/Users/julianeoliveira/Desktop/cat.csv',sep=';')

sns.catplot(x="per_ocu", y="max_str", #hue="State",
            #palette={"male": "g", "female": "m"},
            markers=["o"], linestyles=["-"],
            kind="point", data=titanic)

sns.stripplot(x="per_ocu", y="max_str", data=titanic,size=10, jitter=True)
sns.despine()

sns.regplot(x="per_ocu",
            y="max_str", fit_reg=True, 
            data=titanic);

plt.ylim(0,1)
plt.xlim(0,100)

plt.xlabel("Percentage of ICU occupation",fontsize=16)
plt.ylabel("Maximum value of instrigency ",fontsize=16)
plt.tight_layout()

plt.savefig('max_ocu.png')

r, p = stats.pearsonr(titanic['per_ocu'], titanic['max_str'])
print(f"Scipy computed Pearson r: {r} and p-value: {p}")
    
r, p = stats.spearmanr(titanic['per_ocu'], titanic['max_str'])
print(f"Scipy computed Spearman r: {r} and p-value: {p}")
    
r, p = stats.kendalltau(titanic['per_ocu'], titanic['max_str'])
print(f"Scipy computed Kendall's tau: {r} and p-value: {p}")

# %% [code]
from datetime import datetime

def days_between(d1, d2):
    d1 = datetime.strptime(d1, "%Y-%m-%d")
    d2 = datetime.strptime(d2, "%Y-%m-%d")
    return abs((d2 - d1).days)


days_between('2020-03-24','2020-03-16')

# %% [code]
(-51.8589299847771 +
-65.66849846868327+
-41.6398921062905+
-36.96693770512808+
-60.30378963041908+
-37.16536165653501+
-46.91524786632097+
-66.0688577604271+
-33.337061177696675+
-42.95801698436636+
-32.91754186916487+
-56.61889858290541+
-33.644507168513144+
-59.6383596476075+
-57.30855145414773+
-71.70986350980179)/16

# %% [code]
(-36.94299319081954+
-22.117896238751758+
-36.3552418800142+
-5.153094362413334+
-92.27562567052559+
-22.563799205119476+
-43.59383818617082+
-63.452288176749725+
-43.29753097383823+
-54.95044831657731+
31.949105248456345+
-55.89308017291118+
-60.64575470909814+
-58.45045075341578+
-34.03573387860449+
-49.11857034352659)/16

# %% [code]
names = ['Region','R1','R2','R3']


rt_mean = pd.read_csv('/Users/julianeoliveira/Downloads/Rt_m.csv')
rt_mean = rt_mean.transpose().reset_index()
rt_mean = rt_mean.drop([0])

# %% [code]
rt_mean

# %% [code]
rt_mean.columns

# %% [code]
rt_mean['state'] = rt_mean['index'].astype(str).str[0:2]
rt_mean

# %% [code]
rt_mean['type'] = rt_mean['index'].astype(str).str[3:6]


# %% [code]
rt_mean['dicrease'] = (rt_mean[1] - rt_mean[0])*100/rt_mean[0]

# %% [code]
rt_mean

# %% [code]
rt_meancap = rt_mean[rt_mean.type == 'cap']
rt_meanint = rt_mean[rt_mean.type == 'int']

# %% [code]
rt_meancap[0].mean()

# %% [code]
rt_meanint[0].mean()

# %% [code]
#rt_mean.to_csv('rt_mean.csv')

# %% [code]
rt_mean[(rt_mean['dicrease']> -90)& (rt_mean['dicrease']< -80)]

# %% [code]
print('mean before change in DDD capitals' , rt_meancap[0].mean())

print('mean after change in DDD capitals' , rt_meancap[1].mean(), 'decrease of ', (rt_meancap[1].mean() - rt_meancap[0].mean())*100/rt_meancap[0].mean())

print('mean before change in DDD inland' , rt_meanint[0].mean())

print('mean after change in DDD inland' , rt_meanint[1].mean(), 'decrease of ', (rt_meanint[1].mean() - rt_meanint[0].mean())*100/rt_meanint[0].mean())

# %% [code]
rtDDD = rt_mean[(rt_mean.state !='TO') & (rt_mean.state !='AC') & (rt_mean.state !='SC') & (rt_mean.state !='AP') & (rt_mean.state !='RO') & (rt_mean.state !='AL') & (rt_mean.state !='PI') &  (rt_mean.state !='SE') & (rt_mean.state !='RS') & (rt_mean.state !='MS')]
rtDDD

# %% [code]
rDDDcap = rtDDD[rtDDD.type == 'cap']
rDDDint = rtDDD[rtDDD.type == 'int']

# %% [code]
print('mean before change in DDD capitals' , rDDDcap[0].mean())

print('mean after change in DDD capitals' , rDDDcap[1].mean(), 'decrease of ', (rDDDcap[1].mean() - rDDDcap[0].mean())*100/rDDDcap[0].mean())

print('mean before change in DDD inland' , rDDDint[0].mean())

print('mean after change in DDD inland' , rDDDint[1].mean(), 'decrease of ', (rDDDint[1].mean() - rDDDint[0].mean())*100/rDDDint[0].mean())

# %% [code]
rtDDI = rt_mean[(rt_mean.state =='AP') | (rt_mean.state =='RO') | (rt_mean.state =='AL') | (rt_mean.state =='RS')]
rtDDI

# %% [code]
rDDIcap = rtDDI[rtDDI.type == 'cap']
rDDIint = rtDDI[rtDDI.type == 'int']

# %% [code]
print('mean before change in DDD capitals' , rDDIcap[0].mean())

print('mean after change in DDD capitals' , rDDIcap[1].mean(), 'decrease of ', (rDDIcap[1].mean() - rDDIcap[0].mean())*100/rDDIcap[0].mean())

print('mean before change in DDD inland' , rDDIint[0].mean())

print('mean after change in DDD inland' , rDDIint[1].mean(), 'decrease of ', (rDDIint[1].mean() - rDDIint[0].mean())*100/rDDIint[0].mean())

# %% [code]
rtint[1].mean()

# %% [code]
```
