## Supplementary material for "Assessing the nationwide impact of COVID-19 mitigation policies on the transmission rate of SARS-CoV-2 in Brazil": corssCorr.ipynb

```
# %% [code]
import numpy as np
import matplotlib.pyplot as plt
from scipy.integrate import odeint
from scipy.optimize import least_squares
import math
import pandas as pd
import csv
import datetime
from scipy import stats
from scipy import optimize
from datetime import date, timedelta
import random
import requests
from scipy.stats import truncnorm
import datetime

import statsmodels as sm
#import statsmodels.api as sm

from PIL import Image
from scipy.stats import norm

from statsmodels.tsa.vector_ar.var_model import VAR

import functools

# %% [markdown]
## Mobility data

# %% [code]
mob = pd.read_csv('/Users/julianeoliveira/Desktop/dropbox_15_11_2019/Artigos_preprints/COVID19/Paper_submissions/Paper2/Assessing-the-nationwide-impact-of-COVID-19-mitigation-policies-on-the-transmission-rate-of-SARS-CoV/data/distancing_series.csv')

# %% [code]
mob['date2'] = pd.to_datetime(mob.date)

# %% [code]
mob = mob[(mob.date2 <= '2020-05-22')]

# %% [code]


# %% [markdown]
# Correlation mobility and cases

# %% [code]
names = ['Region','Type','Date','cum_cases','Pop','New']

dta = pd.read_excel('/Users/julianeoliveira/Desktop/dropbox_15_11_2019/Artigos_preprints/COVID19/Paper_submissions/Paper2/Assessing-the-nationwide-impact-of-COVID-19-mitigation-policies-on-the-transmission-rate-of-SARS-CoV/data/allcases.xlsx',header=None,names=names)

dta =dta.drop([0, 2])

dta.head()

# %% [code]
dta =dta.drop([1])

# %% [code]
dta['state'] = dta.Region.str[-2:]

# %% [code]
dta

# %% [code]
dta['state'].unique()

# %% [code]
lista = ['AC', 'AL', 'AM', 'AP', 'BA', 'CE', 'ES', 'GO', 'MA', 'MG', 'MS', 'MT', 'PA', 'PB', 'PE', 'PI', 'PR', 'RJ', 'RN', 'RO', 'RR',
       'RS', 'SC', 'SE', 'SP', 'TO']

# %% [code]
for value in lista:
    est = value
    tipo = 'Interior'


    state = dta[dta.state== est]

    tipoState = state[state.Type == tipo]
    
    tipoState['new_SMA_3'] = tipoState.New.astype(int).rolling(window=3).mean()
    tipoState['new_SMA_7'] = tipoState.New.astype(int).rolling(window=7).mean()
    
    tipoState['GR'] = np.log(round(tipoState['new_SMA_3'],3))/np.log(round(tipoState['new_SMA_7'],3))

    tipoState['GR'].replace(np.inf, 0, inplace=True)

    tipoState['GR'][tipoState['GR'] < 0] = 0

    tipoState['GR'] = tipoState['GR'].fillna(0)
    
    lst = []
    for i in range(0,len(tipoState['New'])):
        c = (tipoState['New'].iloc[i]/tipoState['new_SMA_3'].iloc[i] ) if tipoState['new_SMA_3'].iloc[i] != 0 else 0
        lst.append(c)
    
    tipoState['GR2'] = lst

    tipoState['GR2'].replace(np.inf, 0, inplace=True)

    tipoState['GR2'] = tipoState['GR2'].fillna(0)
    
    mobs = mob[mob.name == est][(mob[mob.name == est].date2 >= tipoState.iloc[0,2])]

    indTipo = 'indInland'
    
    new = pd.DataFrame([tipoState['GR'],tipoState['GR2']]).transpose().reset_index()

    new[indTipo] = mobs[indTipo].values
    new = new.filter(['GR','GR2',indTipo])
    
    xcov = [crosscorr(new[indTipo], new['GR'], lag=i) for i in range(40)]
    print(est, xcov)
    
    plt.figure(figsize=(8,3))

    plt.grid()
    plt.plot(xcov, 'o')

    n =len(xcov)
    l1 = (-1/n) + (2/math.sqrt(n))

    plt.axhline(l1, color='blue', linestyle='dashed', lw=1)

    plt.axhline(-l1, color='blue', linestyle='dashed', lw=1)
    
    plt.show()

# %% [code]
est = 'AL'
tipo = 'Interior'


state = dta[dta.state== est]

tipoState = state[state.Type == tipo]

# %% [code]
tipoState['new_SMA_3'] = tipoState.New.astype(int).rolling(window=3).mean()
tipoState['new_SMA_7'] = tipoState.New.astype(int).rolling(window=7).mean()

# %% [code]
tipoState['GR'] = np.log(round(tipoState['new_SMA_3'],3))/np.log(round(tipoState['new_SMA_7'],3))
#GR = GR.fillna(0).replace(np.inf, 0, inplace=True)

tipoState['GR'].replace(np.inf, 0, inplace=True)

tipoState['GR'][tipoState['GR'] < 0] = 0

tipoState['GR'] = tipoState['GR'].fillna(0)

# %% [code]
lst = []
for i in range(0,len(tipoState['New'])):
    c = (tipoState['New'].iloc[i]/tipoState['new_SMA_3'].iloc[i] ) if tipoState['new_SMA_3'].iloc[i] != 0 else 0
    lst.append(c)
    
tipoState['GR2'] = lst

tipoState['GR2'].replace(np.inf, 0, inplace=True)

#capState['GR2'][capState['GR2'] < 0] = 0

tipoState['GR2'] = tipoState['GR2'].fillna(0)

# %% [code]
mobs = mob[mob.name == est][(mob[mob.name == est].date2 >= tipoState.iloc[0,2])]

indTipo = 'indInland'

# %% [code]
def crosscorr(datax, datay, lag=0):
    """ Lag-N cross correlation. 
    Parameters
    ----------
    lag : int, default 0
    datax, datay : pandas.Series objects of equal length

    Returns
    ----------
    crosscorr : float
    """
    return datax.corr(datay.shift(lag))

# %% [code]
new = pd.DataFrame([tipoState['GR'],tipoState['GR2']]).transpose().reset_index()

new[indTipo] = mobs[indTipo].values
new = new.filter(['GR','GR2',indTipo])

# %% [code]
xcov = [crosscorr(new[indTipo], new['GR2'], lag=i) for i in range(40)]
print(xcov)

# %% [code]
plt.figure(figsize=(8,3))

plt.grid()
plt.plot(xcov, 'o')

n =len(xcov)
l1 = (-1/n) + (2/math.sqrt(n))

plt.axhline(l1, color='blue', linestyle='dashed', lw=1)

plt.axhline(-l1, color='blue', linestyle='dashed', lw=1)


# %% [markdown]
## Stringency

# %% [code]
strg = pd.read_csv('/Users/julianeoliveira/Desktop/dropbox_15_11_2019/Artigos_preprints/COVID19/Paper_submissions/Paper2/Assessing-the-nationwide-impact-of-COVID-19-mitigation-policies-on-the-transmission-rate-of-SARS-CoV/data/stringency_series.csv',sep= ',')

# %% [code]
strg

# %% [code]
strg['date2'] = pd.to_datetime(strg.Date)

# %% [code]
strgs = strg[strg.UF == est][(strg[strg.UF == est].date2 >= tipoState.iloc[0,2])]

# %% [code]
new

# %% [code]
```
