## Supplementary material for "Assessing the nationwide impact of COVID-19 mitigation policies on the transmission rate of SARS-CoV-2 in Brazil": model_SEIIR.py

```
#!/usr/bin/env python3
# -*- coding: utf-8 -*-
"""
Created on Fri Jun 26 16:05:26 2020

@author: lhunlindeion
"""

import numpy as np
from functools import reduce
import scipy.integrate as spi
from scipy.optimize import least_squares, minimize
# from platypus import NSGAII, Problem, Real
from pyswarms.single.global_best import GlobalBestPSO
import pyswarms as ps
from pyswarms.backend.topology import Star
from pyswarms.utils.plotters import plot_cost_history
from itertools import repeat
import multiprocessing as mp
import copy
import joblib
from scipy.stats import poisson

class SEIIR_BETAS:
    ''' SEIIHURD Model'''
    def __init__(self,tamanhoPop,numeroProcessadores=None):
        self.N = tamanhoPop
        self.numeroProcessadores = numeroProcessadores
        self.pos = None

#pars dict betas, delta, kappa, p, gammaA, gammaS, h, epsilon, gammaH, gammaU, muU, muH, wU, wH
# seguindo a notação beta_12 é 2 infectando 1, onde 1 é a linha e 2 a coluna.
    def SEIIR_eq(self, X, t, pars):
        S, E, Ia, Is, R, Nw = X
        StE = S * pars['beta'] * (Is + pars['delta'] * Ia) 
        dS = - StE
        dE = StE - pars['kappa'] * E
        dIa = (1 - pars['p']) * pars['kappa'] * E - pars['gammaA'] * Ia
        dIs = pars['p'] * pars['kappa'] * E - pars['gammaS'] * Is
        dR = pars['gammaA'] * Ia + pars['gammaS'] * Is
        dNw = pars['p'] * pars['kappa'] * E
        return np.r_[dS, dE, dIa, dIs, dR, dNw]
    
    
    def call_ODE(self, ts, ppars):
        atol = min([1e-4/self.N, 1.e-9])
        betas = ppars['beta'].copy()
        pars = copy.deepcopy(ppars)
        if 'tcut' not in ppars.keys():
            tcorte = None
        else:
            tcorte = pars['tcut']
        if type(ts) in [int, float]:
            ts = np.arange(ts)
        if type(tcorte) == type(None) or len(tcorte) == 0:
            tcorte = [ts[-1]]
            if type(betas) != list:
                betas = [betas]
        if tcorte[-1] < ts[-1]:
            tcorte = np.r_[tcorte, ts[-1]]
        tcorte = np.r_[ts[0],  tcorte]
        tcorte.sort()
        Is0 = pars['x0']
        x0 = np.r_[1. - Is0.sum(), Is0, 0, Is0[-1]]
        saida = x0.reshape((1,-1))
        Y = saida.copy()
        for i in range(1, len(tcorte)):
            cut_last = False
            pars['beta'] = betas[i-1]
            t = ts[(ts >= tcorte[i-1]) * (ts<= tcorte[i])]
            if len(t) > 0:
                if t[0] > tcorte[i-1]:
                    t = np.r_[tcorte[i-1], t]
                if t[-1] < tcorte[i]:
                    t = np.r_[t, tcorte[i]]
                    cut_last = True
                Y = spi.odeint(self.SEIIR_eq, Y[-1], t, args=(pars,), atol=1e-4/self.N)
                if cut_last:
                    saida = np.r_[saida, Y[1:-1]]
                else:
                    saida = np.r_[saida, Y[1:]]
            elif not np.isclose(tcorte[i], tcorte[i-1], rtol=1e-9):
                Y = spi.odeint(self.SEIIR_eq, Y[-1], tcorte[i-1:i+1], args=(pars,), atol=atol)

        return ts, saida


    def _fill_paramPSO(self, paramPSO):
        if 'options' not in paramPSO.keys():
            paramPSO['options'] = {'c1': 0.1, 'c2': 0.3, 'w': 0.9,'k':5,'p':2}
        if 'n_particles' not in paramPSO.keys():
            paramPSO['n_particles'] = 100
        if 'iter' not in paramPSO.keys():
            paramPSO['iter'] = 300
        return paramPSO

                
    def _prepare_conversor(self, p2f, pothers, bound):
        padjus = list()
        if  bound != None:
            bound_new = [[], []]
        for i, par in enumerate(p2f):
            if '_ALL' in par:
                name = par.split('_')[0]
                for j in range(len(pothers[name])):
                    padjus.append('{}_{}'.format(name, j))
                    if  bound != None:
                        bound_new[0].append(bound[0][i])
                        bound_new[1].append(bound[1][i])
            else:
                padjus.append(par)
                if  bound != None:
                    bound_new[0].append(bound[0][i])
                    bound_new[1].append(bound[1][i])
        if  bound != None:
            bound_new[0] = np.array(bound_new[0])
            bound_new[1] = np.array(bound_new[1])
        return bound_new, padjus
    
    def conversor(self, coefs, pars0, padjus):
        pars = copy.deepcopy(pars0)
        for i, coef in enumerate(coefs):
            if '_' in padjus[i]:
                name, indx = padjus[i].split('_')
                pars[name][int(indx)] = coef                  
            else:
                pars[padjus[i]] = coef
        return pars
        
    def create_std_bounds(self):
        self.bound = [[1./self.N, 1./21] , [10*self.Y[0]/self.N, 0.2]]
        for i in range(self.nbetas):
            self.bound[0].append(0)
            self.bound[1].append(2)
        for i in range(self.nbetas-1):
            self.bound[0].append(self.t[0]+0.5)
            self.bound[1].append(self.t[-1]-9.5)
        self.bound[0] = np.array(self.bound[0])
        self.bound[1] = np.array(self.bound[1])

    def prepare_to_fit(self, data, t, pars, pars_to_fit, bound=None, stand_error=False):
        self.pars_init = copy.deepcopy(pars)
        self.Y = data
        if type(t) == type(None):
            self.t = np.arange(len(self.Y))
        else:
            self.t = t
        self.bound, self.padjus = self._prepare_conversor(pars_to_fit, pars, bound)
        self.n_to_fit = len(self.padjus)

    
    def negloglikehood(self, coefs):
        '''
        estimates a loglikehood supposing that the new cases follows a
        poisson process with the model new cases as the parameter
        '''
        ts, mY = self.call_ODE(self.t, self.conversor(coefs, self.pars_init, self.padjus))
        mus = np.diff(self.N * mY[:,-1])
        ks = np.diff(self.Y)
        return - (poisson.logpmf(ks, mus)).sum()

    
    def negloglikehood_PSO(self, coefs_list):
        saida = np.zeros(coefs_list.shape[0])
        for i, coefs in enumerate(coefs_list):
            saida[i] = self.negloglikehood(coefs)
        return saida
    
    def BIC(self, coefs='LS'):
        if type(coefs) == type(None):
            coefs = self.pos
        elif type(coefs) == str:
            if coefs  == 'LS':
                coefs = self.pos_ls
            elif coefs == 'ML':
                coefs = self.pos_ml
            elif coefs == 'MLP':
                coefs = self.pos_m
        return len(coefs) * np.log(len(self.Y)) + 2* self.negloglikehood(coefs)
    
    def objectiveFunction(self, coefs_list, stand_error=False):
        errsq = np.zeros(coefs_list.shape[0])
        for i, coefs in enumerate(coefs_list):
            errs = self.residuals(coefs, stand_error)
            errsq[i] = (errs*errs).mean()
        return errsq

    def residuals(self, coefs, stand_error=False):
        ts, mY = self.call_ODE(self.t, self.conversor(coefs, self.pars_init, self.padjus))
        errs = (self.Y- self.N *  mY[:,-1])
        if stand_error:
            errs = errs / np.sqrt(self.N * mY[:,-1] + 1)
        if np.isnan(errs).any():
            print(coefs)
        return errs
    


    def fit(self, data,  pars, pars_to_fit, t=None, bound=None, paramPSO=dict(),  stand_error=False):
        self.prepare_to_fit(data, t, pars, pars_to_fit, bound=bound, stand_error=stand_error)
        paramPSO = self._fill_paramPSO(paramPSO)
        optimizer = ps.single.LocalBestPSO(n_particles=paramPSO['n_particles'], dimensions=len(self.bound[0]), options=paramPSO['options'],bounds=self.bound)
        cost = pos = None
        cost, pos = optimizer.optimize(self.objectiveFunction,paramPSO['iter'],  stand_error=stand_error, n_processes=self.numeroProcessadores)
        self.pos = pos
        self.pars_opt = self.conversor(pos, self.pars_init, self.padjus)
        self.rmse = cost
        # self.optimize = optimizer

    def fit_ML(self, data, pars, pars_to_fit, t=None, bound=None, paramPSO=dict(),  stand_error=False):
        self.prepare_to_fit(data, t, pars, pars_to_fit, bound=bound, stand_error=stand_error)
        paramPSO = self._fill_paramPSO(paramPSO)
        optimizer = ps.single.LocalBestPSO(n_particles=paramPSO['n_particles'], dimensions=len(self.bound[0]), options=paramPSO['options'],bounds=self.bound)
        cost = pos = None
        cost, pos = optimizer.optimize(self.negloglikehood_PSO, paramPSO['iter'],  n_processes=self.numeroProcessadores)
        self.pos_m = pos
        self.pars_opt_m = self.conversor(pos, self.pars_init, self.padjus)
        self.rmse_m = cost
        # self.optimize_m = optimizer

    def fit_ML_min(self,  data, pars, pars_to_fit, t=None,  bound=None, nrand=20, init=None,  stand_error=False):
        self.prepare_to_fit(data, t, pars, pars_to_fit, bound=bound, stand_error=stand_error)
        if type(init) == type(None):
            cost_best = np.inf
            res_best = None
            if type(self.pos) != type(None) or self.numeroProcessadores == None or self.numeroProcessadores <= 1:
                for i in range(nrand):
                    print("{} / {}".format(i, nrand))
                    par0 = np.random.rand(len(self.bound[0]))
                    par0 = self.bound[0] + par0 * (self.bound[1] - self.bound[0])
                    res = minimize(self.negloglikehood, par0, bounds=np.array(self.bound).T)
                    if res.fun < cost_best:
                        cost_best = res.fun
                        res_best = res
            else:
                par0 = np.random.rand(nrand, len(self.bound[0]))
                par0 = self.bound[0].reshape((1,-1)) + par0 * (self.bound[1] - self.bound[0]).reshape((1,-1))
                f = lambda p0: minimize(self.negloglikehood, p0, bounds=np.array(self.bound).T)
                all_res = joblib.Parallel(n_jobs=self.numeroProcessadores)(joblib.delayed(f)(p0,) for p0 in par0)
                costs = np.array([res.fun for res in all_res])
                cost_best = all_res[costs.argmin()].fun
                res_best = all_res[costs.argmin()]
        else:
            res_best = minimize(self.negloglikehood, init, bounds=np.array(self.bound).T)
        self.pos_ml = res_best.x
        self.pars_opt_ml = self.conversor(res_best.x, self.pars_init, self.padjus)
        self.result_ml = res_best

    def fit_lsquares(self, data, pars, pars_to_fit, t=None,  bound=None, nrand=20, init=None,  stand_error=False):
        self.prepare_to_fit(data, t, pars, pars_to_fit, bound=bound, stand_error=stand_error)
        if type(init) == type(None):
            cost_best = np.inf
            res_best = None
            #BUG: the parallel code does not work if PSO code had run previously
            if type(self.pos) != type(None) or self.numeroProcessadores == None or self.numeroProcessadores <= 1:
                for i in range(nrand):
                    print("{} / {}".format(i, nrand))
                    par0 = np.random.rand(len(self.bound[0]))
                    par0 = self.bound[0] + par0 * (self.bound[1] - self.bound[0])
                    res = least_squares(self.residuals, par0, bounds=self.bound, args=(stand_error,))
                    if res.cost < cost_best:
                        cost_best = res.cost
                        res_best = res
            else:
                par0 = np.random.rand(nrand, len(self.bound[0]))
                par0 = self.bound[0].reshape((1,-1)) + par0 * (self.bound[1] - self.bound[0]).reshape((1,-1))
                f = lambda p0: least_squares(self.residuals, p0, bounds=self.bound, args=(stand_error,))
                all_res = joblib.Parallel(n_jobs=self.numeroProcessadores)(joblib.delayed(f)(p0,) for p0 in par0)
                costs = np.array([res.cost for res in all_res])
                cost_best = all_res[costs.argmin()].cost
                res_best = all_res[costs.argmin()]
        else:
            res_best = least_squares(self.residuals, init, bounds=self.bound, args=(stand_error,) )
        self.pos_ls = res_best.x
        self.pars_opt_ls = self.conversor(res_best.x, self.pars_init, self.padjus)
        self.rmse_ls = (res_best.fun**2).mean()
        self.result_ls = res_best
        
    def predict(self, t=None, coefs=None, model_output=False):
        if type(t) == type(None):
            t = self.t
        if type(coefs) == type(None):
            coefs = self.pos
        elif type(coefs) == str:
            if coefs  == 'LS':
                coefs = self.pos_ls
            elif coefs == 'ML':
                coefs = self.pos_ml
            elif coefs == 'MLP':
                coefs = self.pos_m
        ts, mY = self.call_ODE(t, self.conversor(coefs, self.pars_init, self.padjus))
        saida = self.N * mY[:,-1]
        
        if model_output:
            return ts, saida, mY
        else:
            return ts, saida
```
