## Supplementary material for "Assessing the nationwide impact of COVID-19 mitigation policies on the transmission rate of SARS-CoV-2 in Brazil": seiir_bootstrap.py

```
#!/usr/bin/env python3
# -*- coding: utf-8 -*-
"""
Bootstrap + Maximum Likehood + PSO pra todos os estados e separando 
@author: Felipe

"""

import numpy as np
import pandas as pd
from model_SEIIR import SEIIR_BETAS
from scipy.stats import poisson

def gen_bootstrap_serie(serie, iscumul=True):
    if iscumul:
        serie = np.r_[serie[0], np.diff(serie)]
    boots = poisson.rvs(mu=serie)
    if iscumul:
        boots = np.cumsum(boots)
    return boots

capital_dic={
    'AC': 'Rio Branco',
    'AL': 'Maceió',
    'AP': 'Macapá',
    'AM': 'Manaus',
    'BA': 'Salvador',
    'CE': 'Fortaleza',
    'DF': 'Brasília',
    'ES': 'Vitória',
    'GO': 'Goiânia',
    'MA': 'São Luís',
    'MT': 'Cuiabá',
    'MS': 'Campo Grande',
    'MG': 'Belo Horizonte',
    'PA': 'Belém',
    'PB': 'João Pessoa',
    'PR': 'Curitiba',
    'PE': 'Recife',
    'PI': 'Teresina',
    'RJ': 'Rio de Janeiro',
    'RN': 'Natal',
    'RS': 'Porto Alegre',
    'RO': 'Porto Velho',
    'RR': 'Boa Vista',
    'SC': 'Florianópolis',
    'SP': 'São Paulo',
    'SE': 'Aracaju',
    'TO': 'Palmas'
}


df3=pd.read_csv('populacao_municipio.csv')
pop_dic={}

    
for state in df3['ARmaior'].unique():
    pop_state=[]

    Sigla=df3.loc[df3['ARmaior'] == state].NomeMunic.values[0][-2]+df3.loc[df3['ARmaior'] == state].NomeMunic.values[0][-1]
    
    pop_state.append(sum(df3.loc[df3['ARmaior'] == state].Total.values))
    
    Sigla=df3.loc[df3['ARmaior'] == state].NomeMunic.values[0][-2]+df3.loc[df3['ARmaior'] == state].NomeMunic.values[0][-1]
    
    ind1=list(df3.loc[(df3['ARmaior'] == state)& (df3['Sexo'] =='f' )].NomeMunic.values).index(capital_dic[Sigla]+" - "+Sigla)
    ind2=list(df3.loc[(df3['ARmaior'] == state)& (df3['Sexo'] =='m' )].NomeMunic.values).index(capital_dic[Sigla]+" - "+Sigla)

    
    
    pop_state.append(df3.loc[(df3['ARmaior'] == state)& (df3['Sexo'] =='f' )].Total.values[ind1]+df3.loc[(df3['ARmaior'] == state)& (df3['Sexo'] =='m' )].Total.values[ind2])
    
    
    pop_state.append(pop_state[0]-pop_state[1])
    
    pop_dic[Sigla]=pop_state

conv_list={'estado':0, 'capital':1, 'interior':2}
#pop_dic['sigla'] = list with [state, capital, interior]
#%%

pars_names = ['beta0', 'beta1', 'beta2', 'tcut0', 'tcut1', 'delta', 'e0', 'ia0', 'is0']

nthreads = 32
path = "data/"
states = list(capital_dic.keys())
#sub_secs = ['estado', 'capital', 'interior']
sub_secs = ['estado']
ifig = 0
nboots = 100
paramPSO= {'nparticles':150, 'iter':500 }

states = [state for state in states if state == "DF"]
print(states)

for state in states:
    saida = {}
    saida['state'] = list()
    saida['type'] = list()
    for name in pars_names:
        saida[name] = list()
    for sub_sec in sub_secs:
        print(state, sub_sec)
        N = pop_dic[state][conv_list[sub_sec]]
        data = pd.read_csv('{}{}/{}.csv'.format(path, state, sub_sec), sep=',', decimal='.')
        dday = 'date'
        data[dday] = pd.to_datetime(data[dday], yearfirst=True)
        data['DayNum'] = data[dday].dt.dayofyear
        Y0 = data['cases'].to_numpy()
        nneg = np.flatnonzero(np.diff(Y0)<0)
        while len(nneg) > 0:
            Y0[nneg+1] = Y0[nneg]
            nneg = np.flatnonzero(np.diff(Y0)<0)
        t = data['DayNum'].to_numpy()
        bounds = {'x0_ALL': [0, 10./N],
                  'beta_ALL': [0, 2],
                  'p': [0.1, 0.35],
                  'delta': [0., .7],
                  'tcut_0': [t.min()+1, t.max()-1],
                  'tcut_1': [104, t.max()-1]
                  }
        
        I0 = 0.0001 * np.ones(1)
        #Initializing data
        Mb = 1
        param = {'delta': 0.62, #asymptomatic infection correction
            'kappa': 0.25, #exposed decay parameter
            'gammaA': 1./3.5, # asymptomatic decay parameter
            'gammaS': 0.25, # symptomatic decay parameter
            'p': 0.2, #fraction of exposed that becomes symptomatic
            'beta': [1.06 * Mb, 0.63 * Mb], #infectivity matrix
            'tcut': [30.], #time instants of change of the infectivity matrix
            'x0': np.r_[I0, I0, I0] #initial conditions
          }
        if (state in ['DF', 'MT', 'PR']) or (state=='BA' and sub_sec=='interior'):
            param['beta'] = np.array([1. * Mb, 1. * Mb, 1. * Mb])
            param['tcut'] = np.array([70., 80.])
            pars_to_fit = ['beta_ALL', 'tcut_0', 'tcut_1', 'delta', 'x0_ALL']
            pars_names_f = pars_names
            bounds['tcut_0'] = [bounds['tcut_0'][0], 104]
            is3beta = True
        else:
            pars_to_fit = ['beta_ALL', 'tcut_0', 'delta', 'x0_ALL']
            pars_names_f = ['beta0', 'beta1', 'tcut0', 'delta', 'e0', 'ia0', 'is0']
            is3beta = False

        bound = np.array([bounds[key] for key in pars_to_fit])
        bound = [bound[:,0], bound[:,1]]
        for b in range(nboots):
            Y = gen_bootstrap_serie(Y0)
            model = SEIIR_BETAS(N, nthreads)
            model.fit_ML(Y, param, pars_to_fit, t=t, stand_error=True, bound=bound, paramPSO=paramPSO)
            saida['state'].append(state)
            saida['type'].append(sub_sec)
            for i, name in enumerate(pars_names_f):
                saida[name].append(model.pos_m[i])
            if not is3beta:
                saida['tcut1'].append(np.nan)
                saida['beta2'].append(np.nan)


    S = pd.DataFrame(data=saida)
    S.to_csv('seiir_fits_boots_{}.csv'.format(state))  
```
