## Supplementary material for "Assessing the nationwide impact of COVID-19 mitigation policies on the transmission rate of SARS-CoV-2 in Brazil": Sup_Run.ipynb

```
# %% [code]
import numpy as np
import matplotlib.pyplot as pl
from scipy.integrate import odeint
from scipy.optimize import least_squares
import math
import pandas as pd
import csv
import matplotlib.gridspec as gridspec
from scipy import stats
from scipy import optimize
from datetime import date, timedelta
import random
from scipy.stats import truncnorm


df_con=pd.read_csv('data_casos_data_con.csv',sep=';')


state_dic={'BA': 'Bahia'}


capital_dic={
    'AC': 'Rio Branco',
    'AL': 'Maceió',
    'AP': 'Macapá',
    'AM': 'Manaus',
    'BA': 'Salvador',
    'CE': 'Fortaleza',
    'DF': 'Brasília',
    'ES': 'Vitória',
    'GO': 'Goiânia',
    'MA': 'São Luís',
    'MT': 'Cuiabá',
    'MS': 'Campo Grande',
    'MG': 'Belo Horizonte',
    'PA': 'Belém',
    'PB': 'João Pessoa',
    'PR': 'Curitiba',
    'PE': 'Recife',
    'PI': 'Teresina',
    'RJ': 'Rio de Janeiro',
    'RN': 'Natal',
    'RS': 'Porto Alegre',
    'RO': 'Porto Velho',
    'RR': 'Boa Vista',
    'SC': 'Florianópolis',
    'SP': 'São Paulo',
    'SE': 'Aracaju',
    'TO': 'Palmas'
}


df3=pd.read_csv('populacao_municipio.csv')
pop_dic={}

    
for state in df3['ARmaior'].unique():
    pop_state=[]

    Sigla=df3.loc[df3['ARmaior'] == state].NomeMunic.values[0][-2]+df3.loc[df3['ARmaior'] == state].NomeMunic.values[0][-1]
    
    pop_state.append(sum(df3.loc[df3['ARmaior'] == state].Total.values))
    
    Sigla=df3.loc[df3['ARmaior'] == state].NomeMunic.values[0][-2]+df3.loc[df3['ARmaior'] == state].NomeMunic.values[0][-1]
    
    ind1=list(df3.loc[(df3['ARmaior'] == state)& (df3['Sexo'] =='f' )].NomeMunic.values).index(capital_dic[Sigla]+" - "+Sigla)
    ind2=list(df3.loc[(df3['ARmaior'] == state)& (df3['Sexo'] =='m' )].NomeMunic.values).index(capital_dic[Sigla]+" - "+Sigla)

    
    
    pop_state.append(df3.loc[(df3['ARmaior'] == state)& (df3['Sexo'] =='f' )].Total.values[ind1]+df3.loc[(df3['ARmaior'] == state)& (df3['Sexo'] =='m' )].Total.values[ind2])
    
    
    pop_state.append(pop_state[0]-pop_state[1])
    
    pop_dic[Sigla]=pop_state


# %% [code]
#Defining the Steap Function 

 
kb,kappa,gammaA,gammaS=[50,0.25,0.285714286,0.24999375]

def H(t):
        h = 1.0/(1.0+ np.exp(-2.0*kb*t))
        return h


    #Defining the Beta(t)
def beta(t,t1,b,b1):
        beta = b*H(t1-t) + b1*H(t-t1) 
        return beta


#Defining the Model
def SEIIR(f,t,parametros):
    
        #parameters
        b, b1, delta, p, t1 = parametros
    
        #variables
        S = f[0]
        E = f[1]
        IA = f[2]
        IS = f[3]
        R = f[4]
        Nw = f[5]
    
    
        #equations
        dS_dt = - beta(t,t1,b,b1)*S*(IS + delta*IA) 
        dE_dt = beta(t,t1,b,b1)*S*(IS + delta*IA) - kappa * E
        dIA_dt = (1-p)*kappa*E - gammaA*IA
        dIS_dt = p*kappa*E - gammaS*IS      #+(1-(muH))*omega*gammaH*H -gammaU*U
        dR_dt = gammaA*IA +gammaS*IS
    
    
        #epidemic curve
        dNw_dt = p*kappa*E
    
        #Returning the derivatives
        return [dS_dt,dE_dt,dIA_dt,dIS_dt,dR_dt,dNw_dt]

    #Calculating the Error
def lq_SEIIR(pars, ts0):
    
        #assaing the Least square given parameters to the numerical integration for the error calculation
        b, b1, delta, p ,ia0, is0, e0, t1 = pars
    
        #initial conditions
        q0 = [1-ia0 - is0 -e0,e0,ia0,is0,0,0]
    
        #parameters to feed the E.D.Os
        parode = b, b1,delta, p ,t1
    
        #Integrating
        qs = odeint(SEIIR,q0, ts0,args=(parode,),mxstep=1000000)

        #sinf the epidemic curve
        #sdth the death curve

        sinf = qs[:,-1]
    
        #define the standardized residuals
        erri = (N*sinf - infec) / np.sqrt(N*sinf+1)
    
        return np.r_[erri] 


def Fit(s,ts0,dates):


        Comm_Contamin=df_con.loc[df_con['State'] == s]['Date of comunity contamination'].values[0]
        
        if not any(x == Comm_Contamin for x in dates):
            Comm_Contamin=dates[0]        

    #DEEFINE THE INTERVALS OF PARS b, b1, xi, delta, h, ia0, is0, e0, t1
        intervals = np.array(   [ [0., 2.],#b
                              [0., 2.],#b1
                              [0., 1.5],#delta
                              [0.1, 0.35],#p
                              [0.,10./N],#ia0
                              [0.,10./N],#is0
                              [0.,10./N],#e0
                              [list(dates).index(Comm_Contamin),list(dates).index('2020-05-22')]#t1
                              ])

    #NUMBER OF FITS
        n_tries = 30

    #best error starts at inft 
        best_err = np.inf
    #aj the number of best fists 
        j = 0

        for i in range(n_tries):
    

        #create a set of ramdom parameters
            par0 = np.random.rand(len(intervals))
        #Limit those parameters to the interval defined
            par0 = intervals[:,0] + par0 * (intervals[:,1] - intervals[:,0])
    
        #try to fit

            res = least_squares(lambda pars: lq_SEIIR(pars, ts0), par0, bounds=(intervals[:,0], \
                                                 intervals[:,1]))
            ier = res.status
        #if converges
            if ier >= 1 and ier <= 4:
                if res.cost < best_err:
                #best_err = erro
                    best_err = res.cost
                #best_pop = parameters
                    best_pop = res.x
                #+1 best fit
                    j = j +1
                    print('achou',j)


        return best_pop
    
    
    
    
#Defining the Beta(t)
def beta_2(t,t1,t2,b,b1,b2):
        beta = b*H(t1-t) + b1*H(t2-t)*H(t-t1) + b2*H(t-t2)
        return beta


#Defining the Model
def SEIIR_2(f,t,parametros):
    
        #parameters
        b, b1,b2, delta, p, t1,t2 = parametros
    
        #variables
        S = f[0]
        E = f[1]
        IA = f[2]
        IS = f[3]
        R = f[4]
        Nw = f[5]
    
    
        #equations
        dS_dt = - beta_2(t,t1,t2,b,b1,b2)*S*(IS + delta*IA) 
        dE_dt = beta_2(t,t1,t2,b,b1,b2)*S*(IS + delta*IA) - kappa * E
        dIA_dt = (1-p)*kappa*E - gammaA*IA
        dIS_dt = p*kappa*E - gammaS*IS
        dR_dt = gammaA*IA  + gammaS*IS

    
    
        #epidemic curve
        dNw_dt = p*kappa*E
    
        #Returning the derivatives
        return [dS_dt,dE_dt,dIA_dt,dIS_dt,dR_dt,dNw_dt]

    #Calculating the Error
def lq_SEIIR_2(pars, ts0):
    
        #assaing the Least square given parameters to the numerical integration for the error calculation
        b, b1,b2,delta, p ,ia0, is0, e0, t1,t2 = pars
    
        #initial conditions
        q0 = [1-ia0 - is0 -e0,e0,ia0,is0,0,0]
    
        #parameters to feed the E.D.Os
        parode = b, b1,b2,delta, p ,t1,t2
    
        #Integrating
        qs = odeint(SEIIR_2,q0, ts0,args=(parode,),mxstep=1000000)

        #sinf the epidemic curve
        #sdth the death curve

        sinf = qs[:,-1]
    
        #define the standardized residuals
        erri = (N*sinf - infec) / np.sqrt(N*sinf+1)
    
        return np.r_[erri] 

    
def Fit_2(s,ts0,dates):


        Comm_Contamin=df_con.loc[df_con['State'] == s]['Date of comunity contamination'].values[0]
        

        if not any(x == Comm_Contamin for x in dates):
            Comm_Contamin=dates[0]

    #DEEFINE THE INTERVALS OF PARS b, b1, xi, delta, h, ia0, is0, e0, t1
        intervals = np.array(   [ [0., 1.5],#b
                              [0., 2.],#b1
                              [0., 2.],#b2
                              [0., 1.5],#delta
                              [0.1, 0.35],#p
                              [0.,50./N],#ia0
                              [0.,50./N],#is0
                              [0.,50./N],#e0
                              [list(dates).index(Comm_Contamin),list(dates).index('2020-04-12')],#t1
                              [list(dates).index('2020-04-13'),list(dates).index('2020-05-15')]#t1
                              ])

    #NUMBER OF FITS
        n_tries = 30

    #best error starts at inft 
        best_err = np.inf
    #aj the number of best fists 
        j = 0

        for i in range(n_tries):
              

        #create a set of ramdom parameters
            par0 = np.random.rand(len(intervals))
        #Limit those parameters to the interval defined
            par0 = intervals[:,0] + par0 * (intervals[:,1] - intervals[:,0])
    
        #try to fit

            res = least_squares(lambda pars: lq_SEIIR_2(pars, ts0), par0, bounds=(intervals[:,0], \
                                                 intervals[:,1]))
            ier = res.status
        #if converges
            if ier >= 1 and ier <= 4:
                if res.cost < best_err:
                #best_err = erro
                    best_err = res.cost
                #best_pop = parameters
                    best_pop = res.x
                #+1 best fit
                    j = j +1
                    print('achou',j)


        return best_pop
    

# %% [code]
pops_base={'Source':[],'beta_0':[],'beta_1':[],'delta':[],'p':[],'kappa':[],'gammaA':[],'gammaS':[],'Ia0':[],'Is0':[],'E0':[],'t1':[],'t1_date':[],'beta_2':[],'t2':[],'t2_date':[]}
pops= pd.DataFrame(pops_base)
pops.to_csv('Best_Fits.csv',index=False)


for s in state_dic.keys():


    print(s)
      
    if s=='DF':
    #Criar um grid para as figuras usando GridSpec

        #Definir o tamanho do plot que sera usado para cada plot individula tem o mesmo efieto de quando passado para subplot

        fig, ax1 = pl.subplots(figsize=(15.4,10))

        ax=[]
        ax.append(ax1)

    
        k=0
    
    
        for Type_Fit in ['estado']:
        
            #Carrega os dados
            df = pd.read_csv (s +'\\'+Type_Fit +".csv")
        
            dates=df.date.values
        
            ts0=np.array(list(range(len(dates))))+1
        
            infec=df.cases.values
        
            if Type_Fit=='estado':
                y_lim=infec[-1]*1.07
            
            N=int(round(pop_dic[s][k]))
        
        
        
        
        
            #Fitta
        
            best_pop=Fit_2(s,ts0,dates)

        

        
            #Integra
        
        
            b, b1,b2, delta, p , ia0, is0, e0, t1,t2 = best_pop
            best_pop2=[b, b1, delta, p,kappa,gammaA,gammaS , ia0, is0, e0, t1,dates[int(round(t1))],b2,t2,dates[int(round(t2))]]
            
            parode =b, b1,b2,delta, p , t1,t2
            parode1 =b, b,b ,delta, p , t1,t2
            parode2 =b, b1,b1 ,delta, p , t1,t2

            q0 = [1-ia0 - is0 -e0,e0,ia0,is0,0,0]

            #plot the data
            qs = odeint(SEIIR_2,q0, ts0,args=(parode,),mxstep=1000000)
            qs1= odeint(SEIIR_2,q0, ts0,args=(parode1,),mxstep=1000000)
            qs2= odeint(SEIIR_2,q0, ts0,args=(parode2,),mxstep=1000000)

    
            qq =qs[:,-1]
            qq1 =qs1[:,-1]
            qq2 =qs2[:,-1]
        

        

        
        #Computa nos plots
        
            if Type_Fit=='estados':
                # Add x-axis and y-axis
                tft=len(qq)
                for y in range(len(qq)):
                    if qq1[y]>qq[len(qq)-1]:
                        tft=y+1
                        break
            else:
                
                tft=len(qq)
                
            if Type_Fit=='estados':
                # Add x-axis and y-axis
                tft2=len(qq)
                for y in range(len(qq)):
                    if qq1[y]>qq[len(qq)-1]:
                        tft2=y+1
                        break
            else:
                
                tft2=len(qq)
                
                
                

            ax[k].grid(which='major', axis='both', color='black',linewidth=1.,alpha=0.3)
      
        
            ax[k].plot(dates,N*qq,'b-', linewidth=2.5)
            ax[k].plot(dates[:tft], N*qq1[:tft],'--',color='blue', linewidth=2.5)
            ax[k].plot(dates[:tft2], N*qq2[:tft2],'--',color='darkviolet', linewidth=2.5)


            ax[k].scatter(dates,infec,  s=10,color='black',zorder=3)


            ax[k].axvline(t1, 0, 600,c='r',linestyle='--')
            ax[k].axvline(t2, 0, 600,c='r',linestyle='--')


            #ax.set_xlabel('(a)',fontsize=20)
            ax[k].set_ylabel('Cumulative cases',fontsize=18, fontname="Arial")

            
            
            
            
            
            #Save
            save_pop=[s+'_'+Type_Fit]
            for saves in best_pop2:
                save_pop.append(saves)
            
        
            pops.loc[len(pops)]=save_pop
        
            pops.to_csv('Best_Fits.csv',index=False)
        
    
            
 
    
    
            k+=1
    
    
    
    
    
        ax1.set_ylim(0.,y_lim)


        ax1.tick_params(labelsize=15)

        ax1.text(0, 1.10,'(a)', transform=ax1.transAxes,
              fontsize=16, fontweight='bold', va='top', ha='right')


        ax1.xaxis.set_major_locator(pl.MaxNLocator(7))
        pl.setp(ax1.get_xticklabels(), rotation=30)


    
        for tick in ax1.get_xticklabels():
            tick.set_fontname("Arial")
        for tick in ax1.get_yticklabels():
            tick.set_fontname("Arial")  


        
        pl.show()
                 
        fig.savefig(r"Fits\\"+s+".png",bbox_inches='tight')
    
    
    elif any(x == s for x in ['MT','PR','BA']):
        #Criar um grid para as figuras usando GridSpec
        gs = gridspec.GridSpec(nrows = 2, ncols = 4)

        #Definir o tamanho do plot que sera usado para cada plot individula tem o mesmo efieto de quando passado para subplot
        fig=pl.figure(figsize=(15.4,10))

        #Definir espaco em branco entre os plots
        gs.update(wspace = 0.55)
        gs.update(hspace = 0.55)

        ax=[]
        #Criar o layout onde os plots serao gerados. E nessa parte que se define o grid
        ax1 = pl.subplot(gs[0, 1:3]) #Ininicar um plot em branco no centro da primeira linha (0)
        ax2 = pl.subplot(gs[1, :2])  #Ininicar um plot em branco na primeira posicao da segunda linha
        ax3 = pl.subplot(gs[1, 2:])  #Ininicar um plot em branco no segunda posicao da seguna linha

        ax.append(ax1)
        ax.append(ax2)
        ax.append(ax3)
    
        k=0
    
    
        for Type_Fit in ['estado','capital','interior']:

            
            if s=='BA' and (Type_Fit=='estado' or Type_Fit=='capital'):
        
            
        
            #Carrega os dados
                df = pd.read_csv (s +'\\'+Type_Fit +".csv")
        
                dates=df.date.values
        
                ts0=np.array(list(range(len(dates))))+1
        
                infec=df.cases.values
        
                if Type_Fit=='estado':
                    y_lim=infec[-1]*1.07
            
                N=int(round(pop_dic[s][k]))
        
        
        
            #Fitta


                best_pop=Fit(s,ts0,dates)

        
            #Integra
        
        
                b, b1, delta, p , ia0, is0, e0, t1 = best_pop
                best_pop2=[b, b1, delta, p,kappa,gammaA,gammaS , ia0, is0, e0, t1,dates[int(round(t1))],'Null','Null','Null']
            
            
                parode =b, b1,delta, p , t1
                parode1 =b, b,delta, p , t1

                q0 = [1-ia0 - is0 -e0,e0,ia0,is0,0,0]

            #plot the data
                qs = odeint(SEIIR,q0, ts0,args=(parode,),mxstep=1000000)
                qs1= odeint(SEIIR,q0, ts0,args=(parode1,),mxstep=1000000)

    
                qq =qs[:,-1]
                qq1 =qs1[:,-1]
        

        

        
        #Computa nos plots
        
                if Type_Fit=='estados':
                # Add x-axis and y-axis
                    tft=len(qq)
                    for y in range(len(qq)):
                        if qq1[y]>qq[len(qq)-1]:
                            tft=y+1
                            break
                else:
                
                    tft=len(qq)

                ax[k].grid(which='major', axis='both', color='black',linewidth=1.,alpha=0.3)
      
        
                ax[k].plot(dates,N*qq,'b-', linewidth=2.5)
                ax[k].plot(dates[:tft], N*qq1[:tft],'--',color='blue', linewidth=2.5)


                ax[k].scatter(dates,infec,  s=10,color='black',zorder=3)


                ax[k].axvline(t1, 0, 600,c='r',linestyle='--')


                #ax.set_xlabel('(a)',fontsize=20)
                ax[k].set_ylabel('Cumulative cases',fontsize=18, fontname="Arial")

            
            
            
            
            
                #Save
                save_pop=[s+'_'+Type_Fit]
                for saves in best_pop2:
                    save_pop.append(saves)

        
                pops.loc[len(pops)]=save_pop
        
                pops.to_csv('Best_Fits.csv',index=False)
        
    
            
            
            
            
            else:
            
        
                #Carrega os dados
                df = pd.read_csv (s +'\\'+Type_Fit +".csv")
        
                dates=df.date.values
        
                ts0=np.array(list(range(len(dates))))+1
        
                infec=df.cases.values
        
                if Type_Fit=='estado':
                    y_lim=infec[-1]*1.07
            
                N=int(round(pop_dic[s][k]))
        
        
        
            #Fitta
    
                best_pop=Fit_2(s,ts0,dates)
            #Integra
        
        
                b, b1,b2, delta, p , ia0, is0, e0, t1,t2 = best_pop
                best_pop2=[b, b1, delta, p,kappa,gammaA,gammaS , ia0, is0, e0, t1,dates[int(round(t1))],b2,t2,dates[int(round(t2))]]
            
                parode =b, b1,b2,delta, p , t1,t2
                parode1 =b, b,b ,delta, p , t1,t2
                parode2 =b, b1,b1 ,delta, p , t1,t2

                q0 = [1-ia0 - is0 -e0,e0,ia0,is0,0,0]

            #plot the data
                qs = odeint(SEIIR_2,q0, ts0,args=(parode,),mxstep=1000000)
                qs1= odeint(SEIIR_2,q0, ts0,args=(parode1,),mxstep=1000000)
                qs2= odeint(SEIIR_2,q0, ts0,args=(parode2,),mxstep=1000000)

    
                qq =qs[:,-1]
                qq1 =qs1[:,-1]
                qq2 =qs2[:,-1]
        

        

        
        #Computa nos plots
        
                if Type_Fit=='estados':
                # Add x-axis and y-axis
                    tft=len(qq)
                    for y in range(len(qq)):
                        if qq1[y]>qq[len(qq)-1]:
                            tft=y+1
                            break
                else:
                
                    tft=len(qq)
                
                if Type_Fit=='estados':
                # Add x-axis and y-axis
                    tft2=len(qq)
                    for y in range(len(qq)):
                        if qq1[y]>qq[len(qq)-1]:
                            tft2=y+1
                            break
                else:
                
                    tft2=len(qq)
                
                
                

                ax[k].grid(which='major', axis='both', color='black',linewidth=1.,alpha=0.3)
      
        
                ax[k].plot(dates,N*qq,'b-', linewidth=2.5)
                ax[k].plot(dates[:tft], N*qq1[:tft],'--',color='blue', linewidth=2.5)
                ax[k].plot(dates[:tft2], N*qq2[:tft2],'--',color='darkviolet', linewidth=2.5)


                ax[k].scatter(dates,infec,  s=10,color='black',zorder=3)


                ax[k].axvline(t1, 0, 600,c='r',linestyle='--')
                ax[k].axvline(t2, 0, 600,c='r',linestyle='--')


                #ax.set_xlabel('(a)',fontsize=20)
                ax[k].set_ylabel('Cumulative cases',fontsize=18, fontname="Arial")

            
            
            
            
            
            #Save
                save_pop=[s+'_'+Type_Fit]
                for saves in best_pop2:
                    save_pop.append(saves)
            
        
                pops.loc[len(pops)]=save_pop
        
                pops.to_csv('Best_Fits.csv',index=False)
        
    
            
            
            
            
            
            
            
            k+=1
    
        ax1.set_ylim(0.,y_lim)
            
        ax2.set_ylim(0.,y_lim)
        ax3.set_ylim(0.,y_lim)
    
                 
                 


        ax2.tick_params(labelsize=15)
        ax3.tick_params(labelsize=15)
        ax1.tick_params(labelsize=15)

        ax1.text(0, 1.10,'(a)', transform=ax1.transAxes,
              fontsize=16, fontweight='bold', va='top', ha='right')

        ax2.text(0, 1.10,'(b)', transform=ax2.transAxes,
              fontsize=16, fontweight='bold', va='top', ha='right')

        ax3.text(0, 1.10,'(c)', transform=ax3.transAxes,
              fontsize=16, fontweight='bold', va='top', ha='right')


        ax1.xaxis.set_major_locator(pl.MaxNLocator(7))
        pl.setp(ax1.get_xticklabels(), rotation=30)

        ax2.xaxis.set_major_locator(pl.MaxNLocator(7))
        pl.setp(ax2.get_xticklabels(), rotation=30)

        ax3.xaxis.set_major_locator(pl.MaxNLocator(7))
        pl.setp(ax3.get_xticklabels(), rotation=30)


    
        for tick in ax1.get_xticklabels():
            tick.set_fontname("Arial")
        for tick in ax1.get_yticklabels():
            tick.set_fontname("Arial")  

        for tick in ax2.get_xticklabels():
            tick.set_fontname("Arial")
        for tick in ax2.get_yticklabels():
            tick.set_fontname("Arial")  
    
        for tick in ax3.get_xticklabels():
            tick.set_fontname("Arial")
        for tick in ax3.get_yticklabels():
            tick.set_fontname("Arial")  

        
        pl.show()
                 
        fig.savefig(r"Fits\\"+s+".png",bbox_inches='tight')
   
    
    
    
    
    
    else:

        #Criar um grid para as figuras usando GridSpec
        gs = gridspec.GridSpec(nrows = 2, ncols = 4)

        #Definir o tamanho do plot que sera usado para cada plot individula tem o mesmo efieto de quando passado para subplot
        fig=pl.figure(figsize=(15.4,10))

        #Definir espaco em branco entre os plots
        gs.update(wspace = 0.55)
        gs.update(hspace = 0.55)

        ax=[]
        #Criar o layout onde os plots serao gerados. E nessa parte que se define o grid
        ax1 = pl.subplot(gs[0, 1:3]) #Ininicar um plot em branco no centro da primeira linha (0)
        ax2 = pl.subplot(gs[1, :2])  #Ininicar um plot em branco na primeira posicao da segunda linha
        ax3 = pl.subplot(gs[1, 2:])  #Ininicar um plot em branco no segunda posicao da seguna linha

        ax.append(ax1)
        ax.append(ax2)
        ax.append(ax3)
    
        k=0
    
    
        for Type_Fit in ['estado','capital','interior']:


        
            #Carrega os dados
            df = pd.read_csv (s +'\\'+Type_Fit +".csv")
        
            dates=df.date.values
        
            ts0=np.array(list(range(len(dates))))+1
        
            infec=df.cases.values
        
            if Type_Fit=='estado':
                y_lim=infec[-1]*1.07
            
            N=int(round(pop_dic[s][k]))
        
        
        
            #Fitta


            best_pop=Fit(s,ts0,dates)

        
            #Integra
        
        
            b, b1, delta, p , ia0, is0, e0, t1 = best_pop
            best_pop2=[b, b1, delta, p,kappa,gammaA,gammaS , ia0, is0, e0, t1,dates[int(round(t1))],'Null','Null','Null']
            
            
            parode =b, b1,delta, p , t1
            parode1 =b, b,delta, p , t1

            q0 = [1-ia0 - is0 -e0,e0,ia0,is0,0,0]

            #plot the data
            qs = odeint(SEIIR,q0, ts0,args=(parode,),mxstep=1000000)
            qs1= odeint(SEIIR,q0, ts0,args=(parode1,),mxstep=1000000)

    
            qq =qs[:,-1]
            qq1 =qs1[:,-1]
        

        

        
        #Computa nos plots
        
            if Type_Fit=='estados':
                # Add x-axis and y-axis
                tft=len(qq)
                for y in range(len(qq)):
                    if qq1[y]>qq[len(qq)-1]:
                        tft=y+1
                        break
            else:
                
                tft=len(qq)

            ax[k].grid(which='major', axis='both', color='black',linewidth=1.,alpha=0.3)
      
        
            ax[k].plot(dates,N*qq,'b-', linewidth=2.5)
            ax[k].plot(dates[:tft], N*qq1[:tft],'--',color='blue', linewidth=2.5)


            ax[k].scatter(dates,infec,  s=10,color='black',zorder=3)


            ax[k].axvline(t1, 0, 600,c='r',linestyle='--')


            #ax.set_xlabel('(a)',fontsize=20)
            ax[k].set_ylabel('Cumulative cases',fontsize=18, fontname="Arial")

            
            
            
            
            
            #Save
            save_pop=[s+'_'+Type_Fit]
            for saves in best_pop2:
                save_pop.append(saves)

        
            pops.loc[len(pops)]=save_pop
        
            pops.to_csv('Best_Fits.csv',index=False)
        
    
            
            
            
            
            
            
            
            k+=1
    
        #ax1.set_ylim(0.,y_lim)
        #ax2.set_ylim(0.,y_lim)
        #ax3.set_ylim(0.,y_lim)
    
                 
                 


        ax2.tick_params(labelsize=15)
        ax3.tick_params(labelsize=15)
        ax1.tick_params(labelsize=15)

        ax1.text(0, 1.10,'(a)', transform=ax1.transAxes,
              fontsize=16, fontweight='bold', va='top', ha='right')

        ax2.text(0, 1.10,'(b)', transform=ax2.transAxes,
              fontsize=16, fontweight='bold', va='top', ha='right')

        ax3.text(0, 1.10,'(c)', transform=ax3.transAxes,
              fontsize=16, fontweight='bold', va='top', ha='right')


        ax1.xaxis.set_major_locator(pl.MaxNLocator(7))
        pl.setp(ax1.get_xticklabels(), rotation=30)

        ax2.xaxis.set_major_locator(pl.MaxNLocator(7))
        pl.setp(ax2.get_xticklabels(), rotation=30)

        ax3.xaxis.set_major_locator(pl.MaxNLocator(7))
        pl.setp(ax3.get_xticklabels(), rotation=30)


    
        for tick in ax1.get_xticklabels():
            tick.set_fontname("Arial")
        for tick in ax1.get_yticklabels():
            tick.set_fontname("Arial")  

        for tick in ax2.get_xticklabels():
            tick.set_fontname("Arial")
        for tick in ax2.get_yticklabels():
            tick.set_fontname("Arial")  
    
        for tick in ax3.get_xticklabels():
            tick.set_fontname("Arial")
        for tick in ax3.get_yticklabels():
            tick.set_fontname("Arial")  

        
        pl.show()
                 
        fig.savefig(r"Fits\\"+s+".png",bbox_inches='tight')
   
    
    

      

# %% [code]
#(Fjj - Fii)
def Prod(i,F):
    
    P=1
    
    for j in range(len(F)):
        if i!=j:
          
            P= P*(F[j]-F[i])
    
    return P

#g(x)
def g(x,F):

    g=0
    
    for i in range(len(F)):
        
        if 0!=Prod(i,F): 
            g += np.exp(-F[i]*x)/Prod(i,F)
        
    g=np.prod(F)*g
    
    return g
    

    
#Integral b(t-x)g(x) dx
def Int(b,t,F):

    Int=0
    for x in range(t+1):
        
        Int += b[t-x]*g(x,F)
        
    return Int

# R(t)
def Rt(b,par):


    
    #Defining the F matrix array
    F=np.array([par[4:7]])[0]
    
    #Data
    
    Rt=[]
    
    for t in range(len(b)):
        
        Rt.append(b[t]/Int(b,t,F))
        
    Rt=np.array(Rt)
    

    return Rt

def Rtdupl(b):
    
    Rt=[]
    
    Rt.append(b[0]/1)
    for t in range(len(b)-1):
        
        if b[t]==0:
            Rt.append(b[t+1]/1)
        else:
            Rt.append(b[t+1]/b[t])
        
    Rt=np.array(Rt)
    
    return Rt
    
    

# %% [code]
df_pops=pd.read_csv('Best_Fits.csv')

m=3

def cut(s,Type_Fit):
    
    
    if s=='AC':
        cut=50
    elif s=='AL' and (Type_Fit=='estado' or Type_Fit=='capital'):
        cut=80    
    elif s=='AL' and Type_Fit=='interior':
        cut=40
    elif s=='CE':
        cut=80
    elif s=='PA':
        cut=60
    elif s=='PE':
        cut=70
    elif s=='PI':
        cut=60
    elif s=='RJ':
        cut=50
    elif s=='RN':
        cut=50
    elif s=='SE':
        cut=70
    elif s=='TO'and (Type_Fit=='interior' or Type_Fit=='capital'):
        cut=40
    elif s=='TO' and Type_Fit=='estado':
        cut=60
    else:
        cut=30
        
    return cut


dataEstado={'date':[],'state':[],'cases':[],'population':[],'Infec':[]}
pops= pd.DataFrame(dataEstado)
pops.to_csv('dataestado.csv',index=False)

dataCapital={'date':[],'state':[],'cases':[],'population':[],'Infec':[]}
pops= pd.DataFrame(dataCapital)
pops.to_csv('datacapital.csv',index=False)

dataInterior={'date':[],'state':[],'cases':[],'population':[],'Infec':[]}
pops= pd.DataFrame(dataInterior)
pops.to_csv('datainterior.csv',index=False)


RtEstado={'date':[],'state':[],'population':[],'Rtdata':[],'Rtmod':[]}
pops= pd.DataFrame(RtEstado)
pops.to_csv('Rtestado.csv',index=False)

RtCapital={'date':[],'state':[],'population':[],'Rtdata':[],'Rtmod':[]}
pops= pd.DataFrame(RtCapital)
pops.to_csv('Rtcapital.csv',index=False)

RtInterior={'date':[],'state':[],'population':[],'Rtdata':[],'Rtmod':[]}
pops= pd.DataFrame(RtInterior)
pops.to_csv('Rtinterior.csv',index=False)


for s in state_dic.keys():
#for s in ['PR']:
    print(s)
    
    
    
    
    y_lim=[]
    
    if s=='DF':

            
    #Criar um grid para as figuras usando GridSpec

        #Definir o tamanho do plot que sera usado para cada plot individula tem o mesmo efieto de quando passado para subplot

        fig, ax1 = pl.subplots(figsize=(15.4,10))

        ax=[]
        ax.append(ax1)

    
        k=0
    
    
        for Type_Fit in ['estado']:
        

            #Load the Data
            df = pd.read_csv (s +'\\'+Type_Fit +".csv")
        
            par=df_pops.loc[df_pops['Source']==s +'_'+Type_Fit].values[0][1:]
            
            dates_t=df.date.values
        
            
        
            data_t=df.cases.values
            
            
            N=int(round(pop_dic[s][k]))
        

        
            dates=dates_t[m:]
            data_noise=np.diff(data_t)
            rangeof=np.arange(len(data_noise)- 2*m)+m
            data=[np.mean(data_noise[i-m:i+m]) for i in rangeof]
            
            ts0=np.array(list(range(len(dates_t))))+1
            

        

        
            
        
        
            

        

            b, b1,delta, p,kappa,gammaA,gammaS, ia0, is0, e0, t1,t1_date,b2,t2,t2_date = par
            
            b2=float(b2)
            t2=float(t2)
            
            parode =b, b1,b2,delta, p, t1,t2


            q0 = [1-ia0 - is0 -e0,e0,ia0,is0,0,0]


            qs = odeint(SEIIR_2,q0, ts0,args=(parode,),mxstep=1000000)


            qq=N*qs[:,-1]
    
            sim_data =N*np.diff(qs[:,-1])

        
        #R(t)
        
            Rt_data=Rt(data,par)
    
            Rt_sim=Rt(sim_data,par)

        
        

        
        #Plotting

      
            w=0
            for ij in range(len(data_t)):
                
                if data_t[ij] > cut(s,Type_Fit):
                    w=ij
                    break
        
        
            y_lim.append(max(Rt_data[w:]))
        
            


            
            
        
            tf=len(data)
        
            ax[k].plot(dates[w:tf], Rt_sim[w+m:tf+m],color='blue',linestyle='--', linewidth=2.5,label=r'$R(t)$ From the Model Simulations')
            ax[k].plot(dates[w:tf], Rt_data[w:tf],'k-', linewidth=2.5,label=r'$R(t)$ From the Data')
        
        
        
            ax[k].grid(which='major', axis='both', color='black',linewidth=1.,alpha=0.3)
      
            ax[k].axhline(1, 0, 600,color='r',linestyle='--',linewidth=2.)


            #ax.set_xlabel('(a)',fontsize=20)
            ax[k].set_ylabel(r'$ R(t)$',fontsize=20, fontname="Arial")

            
            
      
        
        
            pops=pd.read_csv('data'+Type_Fit+'.csv')
            
            
            for save in range(len(dates_t)):
                save_pop=[dates_t[save],s,data_t[save],N,qq[save]]


                pops.loc[len(pops)]=save_pop
        
            pops.to_csv('data'+Type_Fit+'.csv',index=False)
            
            
        
            pops=pd.read_csv('Rt'+Type_Fit+'.csv')
            
            
            for save2 in range(len(dates[w:tf])):

                save_pop=[dates[w:tf][save2],s,N,Rt_data[w:tf][save2],Rt_sim[w+m:tf+m][save2]]

                
                pops.loc[len(pops)]=save_pop
        
            pops.to_csv('Rt'+Type_Fit+'.csv',index=False)
        
        
        
        
        
        
        
        
        
        

            k+=1
    
    
    


        ax1.tick_params(labelsize=15)

        ax1.text(0, 1.10,'(a)', transform=ax1.transAxes,
              fontsize=16, fontweight='bold', va='top', ha='right')


        ax1.xaxis.set_major_locator(pl.MaxNLocator(7))
        pl.setp(ax1.get_xticklabels(), rotation=30)


    
        for tick in ax1.get_xticklabels():
            tick.set_fontname("Arial")
        for tick in ax1.get_yticklabels():
            tick.set_fontname("Arial")  


        
        pl.show()
                 
        fig.savefig(r"Figs\\"+s+".png",bbox_inches='tight')
        
        
        
        
    elif any(x == s for x in ['BA','MT','PR']):
        
    
        #Criar um grid para as figuras usando GridSpec
        gs = gridspec.GridSpec(nrows = 2, ncols = 4)

        #Definir o tamanho do plot que sera usado para cada plot individula tem o mesmo efieto de quando passado para subplot
        fig=pl.figure(figsize=(15.4,10))

        #Definir espaco em branco entre os plots
        gs.update(wspace = 0.55)
        gs.update(hspace = 0.55)

        ax=[]
        #Criar o layout onde os plots serao gerados. E nessa parte que se define o grid
        ax1 = pl.subplot(gs[0, 1:3]) #Ininicar um plot em branco no centro da primeira linha (0)
        ax2 = pl.subplot(gs[1, :2])  #Ininicar um plot em branco na primeira posicao da segunda linha
        ax3 = pl.subplot(gs[1, 2:])  #Ininicar um plot em branco no segunda posicao da seguna linha

        ax.append(ax1)
        ax.append(ax2)
        ax.append(ax3)
    
        k=0
    
    
        for Type_Fit in ['estado','capital','interior']:

        
        
            if s=='BA' and (Type_Fit=='estado' or Type_Fit=='capital'):
             
                            #Load the Data
                df = pd.read_csv (s +'\\'+Type_Fit +".csv")
        
                par=df_pops.loc[df_pops['Source']==s +'_'+Type_Fit].values[0][1:]
            
                dates_t=df.date.values
        
            
        
                data_t=df.cases.values
            
            
                N=int(round(pop_dic[s][k]))
        

        
                dates=dates_t[m:]
                data_noise=np.diff(data_t)
                rangeof=np.arange(len(data_noise)- 2*m)+m
                data=[np.mean(data_noise[i-m:i+m]) for i in rangeof]
            
                ts0=np.array(list(range(len(dates_t))))+1
        

        
            #Integrates
        
        
            
        
                b, b1,delta, p,kappa,gammaA,gammaS, ia0, is0, e0, t1,t1_date,b2,t2,t2_date = par
            
            
            
                parode =b, b1, delta, p, t1


                q0 = [1-ia0 - is0 -e0,e0,ia0,is0,0,0]


                qs = odeint(SEIIR,q0, ts0,args=(parode,),mxstep=1000000)


                qq=N*qs[:,-1]
                sim_data =N*np.diff(qs[:,-1])

        
        #R(t)
        
                Rt_data=Rt(data,par)
    
                Rt_sim=Rt(sim_data,par)

        
        

        
        #Plotting

        
        
                w=0
                for ij in range(len(data_t)):
                
                    if data_t[ij] >cut(s,Type_Fit):
                        w=ij
                        break
        
        
                y_lim.append(max(Rt_data[w:]))
        
            


            
            
        
                tf=len(data)
        
                ax[k].plot(dates[w:tf], Rt_sim[w+m:tf+m],color='blue',linestyle='--', linewidth=2.5,label=r'$R(t)$ From the Model Simulations')
                ax[k].plot(dates[w:tf], Rt_data[w:tf],'k-', linewidth=2.5,label=r'$R(t)$ From the Data')
        
        
        
                ax[k].grid(which='major', axis='both', color='black',linewidth=1.,alpha=0.3)
      
                ax[k].axhline(1, 0, 600,color='r',linestyle='--',linewidth=2.)


                #ax.set_xlabel('(a)',fontsize=20)
                ax[k].set_ylabel(r'$ R(t)$',fontsize=20, fontname="Arial")

         
        
        
        
        
        
        
        
        
        
        
                pops=pd.read_csv('data'+Type_Fit+'.csv')
            
            
                for save in range(len(dates_t)):
                    save_pop=[dates_t[save],s,data_t[save],N,qq[save]]


                    pops.loc[len(pops)]=save_pop
        
                pops.to_csv('data'+Type_Fit+'.csv',index=False)
            
            
        
                pops=pd.read_csv('Rt'+Type_Fit+'.csv')
            
            
                for save2 in range(len(dates[w:tf])):

                    save_pop=[dates[w:tf][save2],s,N,Rt_data[w:tf][save2],Rt_sim[w+m:tf+m][save2]]

                
                    pops.loc[len(pops)]=save_pop
        
                pops.to_csv('Rt'+Type_Fit+'.csv',index=False)
        
        
        
        
            
            
            
                
                
            else:
            #Load the Data
                df = pd.read_csv (s +'\\'+Type_Fit +".csv")
        
                par=df_pops.loc[df_pops['Source']==s +'_'+Type_Fit].values[0][1:]
            
                dates_t=df.date.values
        
            
        
                data_t=df.cases.values
            
            
                N=int(round(pop_dic[s][k]))
        

        
                dates=dates_t[m:]
                data_noise=np.diff(data_t)
                rangeof=np.arange(len(data_noise)- 2*m)+m
                data=[np.mean(data_noise[i-m:i+m]) for i in rangeof]
            
                ts0=np.array(list(range(len(dates_t))))+1
            
            

        

        
        

        
            #Integrates
        
        
                b, b1,delta, p,kappa,gammaA,gammaS, ia0, is0, e0, t1,t1_date,b2,t2,t2_date = par
            
                b2=float(b2)
                t2=float(t2)
            
                parode =b, b1,b2,delta, p, t1,t2


                q0 = [1-ia0 - is0 -e0,e0,ia0,is0,0,0]


                qs = odeint(SEIIR_2,q0, ts0,args=(parode,),mxstep=1000000)


                qq=N*qs[:,-1]
                sim_data =N*np.diff(qs[:,-1])

        
        #R(t)
        
                Rt_data=Rt(data,par)
    
                Rt_sim=Rt(sim_data,par)

        
        

        
        #Plotting

        
        
        
       
                w=0
                for ij in range(len(data_t)):
                
                    if data_t[ij] > cut(s,Type_Fit):
                        w=ij
                        break
        
        
                y_lim.append(max(Rt_data[w:]))
        
            


            
            
        
                tf=len(data)
        
                ax[k].plot(dates[w:tf], Rt_sim[w+m:tf+m],color='blue',linestyle='--', linewidth=2.5,label=r'$R(t)$ From the Model Simulations')
                ax[k].plot(dates[w:tf], Rt_data[w:tf],'k-', linewidth=2.5,label=r'$R(t)$ From the Data')
        
        
                ax[k].grid(which='major', axis='both', color='black',linewidth=1.,alpha=0.3)
      
                ax[k].axhline(1, 0, 600,color='r',linestyle='--',linewidth=2.)


            #ax.set_xlabel('(a)',fontsize=20)
                ax[k].set_ylabel(r'$ R(t)$',fontsize=20, fontname="Arial")

         
        
        
        
        
      
        
        
                pops=pd.read_csv('data'+Type_Fit+'.csv')
            
            
                for save in range(len(dates_t)):
                    save_pop=[dates_t[save],s,data_t[save],N,qq[save]]


                    pops.loc[len(pops)]=save_pop
        
                pops.to_csv('data'+Type_Fit+'.csv',index=False)
            
            
        
                pops=pd.read_csv('Rt'+Type_Fit+'.csv')
            
            
                for save2 in range(len(dates[w:tf])):

                    save_pop=[dates[w:tf][save2],s,N,Rt_data[w:tf][save2],Rt_sim[w+m:tf+m][save2]]

                
                    pops.loc[len(pops)]=save_pop
        
                pops.to_csv('Rt'+Type_Fit+'.csv',index=False)
        
        
        
        
        
        
        
        
        
        
        
        
        
         
            k+=1


        
        
        ax1.set_ylim(0., max(y_lim))
        ax2.set_ylim(0., max(y_lim))
        ax3.set_ylim(0., max(y_lim))

        
        ax2.tick_params(labelsize=15)
        ax3.tick_params(labelsize=15)
        ax1.tick_params(labelsize=15)

        ax1.text(0, 1.10,'(a)', transform=ax1.transAxes,
              fontsize=16, fontweight='bold', va='top', ha='right')

        ax2.text(0, 1.10,'(b)', transform=ax2.transAxes,
              fontsize=16, fontweight='bold', va='top', ha='right')

        ax3.text(0, 1.10,'(c)', transform=ax3.transAxes,
              fontsize=16, fontweight='bold', va='top', ha='right')


        ax1.xaxis.set_major_locator(pl.MaxNLocator(7))
        pl.setp(ax1.get_xticklabels(), rotation=30)

        ax2.xaxis.set_major_locator(pl.MaxNLocator(7))
        pl.setp(ax2.get_xticklabels(), rotation=30)

        ax3.xaxis.set_major_locator(pl.MaxNLocator(7))
        pl.setp(ax3.get_xticklabels(), rotation=30)


    
        for tick in ax1.get_xticklabels():
            tick.set_fontname("Arial")
        for tick in ax1.get_yticklabels():
            tick.set_fontname("Arial")  

        for tick in ax2.get_xticklabels():
            tick.set_fontname("Arial")
        for tick in ax2.get_yticklabels():
            tick.set_fontname("Arial")  
    
        for tick in ax3.get_xticklabels():
            tick.set_fontname("Arial")
        for tick in ax3.get_yticklabels():
            tick.set_fontname("Arial")  

        
        pl.show()
                 
        fig.savefig(r"Figs\\"+s+".png",bbox_inches='tight')
        
        
        
        
        
    else:
        
        #Criar um grid para as figuras usando GridSpec
        gs = gridspec.GridSpec(nrows = 2, ncols = 4)

        #Definir o tamanho do plot que sera usado para cada plot individula tem o mesmo efieto de quando passado para subplot
        fig=pl.figure(figsize=(15.4,10))

        #Definir espaco em branco entre os plots
        gs.update(wspace = 0.55)
        gs.update(hspace = 0.55)

        ax=[]
        #Criar o layout onde os plots serao gerados. E nessa parte que se define o grid
        ax1 = pl.subplot(gs[0, 1:3]) #Ininicar um plot em branco no centro da primeira linha (0)
        ax2 = pl.subplot(gs[1, :2])  #Ininicar um plot em branco na primeira posicao da segunda linha
        ax3 = pl.subplot(gs[1, 2:])  #Ininicar um plot em branco no segunda posicao da seguna linha

        ax.append(ax1)
        ax.append(ax2)
        ax.append(ax3)
    
        k=0
    
    
        for Type_Fit in ['estado','capital','interior']:

            

            
            #Load the Data
            df = pd.read_csv (s +'\\'+Type_Fit +".csv")
        
            par=df_pops.loc[df_pops['Source']==s +'_'+Type_Fit].values[0][1:]
            
            dates_t=df.date.values
        
            
        
            data_t=df.cases.values
            
            
            N=int(round(pop_dic[s][k]))
        

        
            dates=dates_t[m:]
            data_noise=np.diff(data_t)
            rangeof=np.arange(len(data_noise)- 2*m)+m
            data=[np.mean(data_noise[i-m:i+m]) for i in rangeof]
            
            ts0=np.array(list(range(len(dates_t))))+1
        

        
            #Integrates
        
        
            
        
            b, b1,delta, p,kappa,gammaA,gammaS, ia0, is0, e0, t1,t1_date,b2,t2,t2_date = par
            
            
            
            parode =b, b1, delta, p, t1


            q0 = [1-ia0 - is0 -e0,e0,ia0,is0,0,0]


            qs = odeint(SEIIR,q0, ts0,args=(parode,),mxstep=1000000)


            qq=N*qs[:,-1]
            sim_data =N*np.diff(qs[:,-1])

        
        #R(t)
        
            Rt_data=Rt(data,par)
    
            Rt_sim=Rt(sim_data,par)

        
        

        
        #Plotting

        
        
            w=0
            for ij in range(len(data_t)):
                
                if data_t[ij] > cut(s,Type_Fit):
                    w=ij
                    break
        
        
            y_lim.append(max(Rt_data[w:]))
        
            


            
            
        
            tf=len(data)
        
            ax[k].plot(dates[w:tf], Rt_sim[w+m:tf+m],color='blue',linestyle='--', linewidth=2.5,label=r'$R(t)$ From the Model Simulations')
            ax[k].plot(dates[w:tf], Rt_data[w:tf],'k-', linewidth=2.5,label=r'$R(t)$ From the Data')
        
        
        
            ax[k].grid(which='major', axis='both', color='black',linewidth=1.,alpha=0.3)
      
            ax[k].axhline(1, 0, 600,color='r',linestyle='--',linewidth=2.)


            #ax.set_xlabel('(a)',fontsize=20)
            ax[k].set_ylabel(r'$ R(t)$',fontsize=20, fontname="Arial")

         
        
        
        
        
        
        
        
        
        
        
            pops=pd.read_csv('data'+Type_Fit+'.csv')
            
            
            for save in range(len(dates_t)):
                save_pop=[dates_t[save],s,data_t[save],N,qq[save]]


                pops.loc[len(pops)]=save_pop
        
            pops.to_csv('data'+Type_Fit+'.csv',index=False)
            
            
        
            pops=pd.read_csv('Rt'+Type_Fit+'.csv')
            
            
            for save2 in range(len(dates[w:tf])):

                save_pop=[dates[w:tf][save2],s,N,Rt_data[w:tf][save2],Rt_sim[w+m:tf+m][save2]]

                
                pops.loc[len(pops)]=save_pop
        
            pops.to_csv('Rt'+Type_Fit+'.csv',index=False)
        
        
        
        
        
        
        
        
        
        
        
        
        
            k+=1


        
        
        ax1.set_ylim(0., max(y_lim))
        ax2.set_ylim(0., max(y_lim))
        ax3.set_ylim(0., max(y_lim))

        
        ax2.tick_params(labelsize=15)
        ax3.tick_params(labelsize=15)
        ax1.tick_params(labelsize=15)

        ax1.text(0, 1.10,'(a)', transform=ax1.transAxes,
              fontsize=16, fontweight='bold', va='top', ha='right')

        ax2.text(0, 1.10,'(b)', transform=ax2.transAxes,
              fontsize=16, fontweight='bold', va='top', ha='right')

        ax3.text(0, 1.10,'(c)', transform=ax3.transAxes,
              fontsize=16, fontweight='bold', va='top', ha='right')


        ax1.xaxis.set_major_locator(pl.MaxNLocator(7))
        pl.setp(ax1.get_xticklabels(), rotation=30)

        ax2.xaxis.set_major_locator(pl.MaxNLocator(7))
        pl.setp(ax2.get_xticklabels(), rotation=30)

        ax3.xaxis.set_major_locator(pl.MaxNLocator(7))
        pl.setp(ax3.get_xticklabels(), rotation=30)


    
        for tick in ax1.get_xticklabels():
            tick.set_fontname("Arial")
        for tick in ax1.get_yticklabels():
            tick.set_fontname("Arial")  

        for tick in ax2.get_xticklabels():
            tick.set_fontname("Arial")
        for tick in ax2.get_yticklabels():
            tick.set_fontname("Arial")  
    
        for tick in ax3.get_xticklabels():
            tick.set_fontname("Arial")
        for tick in ax3.get_yticklabels():
            tick.set_fontname("Arial")  

        
        pl.show()
                 
        fig.savefig(r"Figs\\"+s+".png",bbox_inches='tight')
   
    
    
   

# %% [code]
```
