## Supplementary material for "Assessing the nationwide impact of COVID-19 mitigation policies on the transmission rate of SARS-CoV-2 in Brazil": Transmission rate.ipynb

```
# %% [markdown]
## Analysis of the trasnmission rate variation 

\begin{equation}\label{beta}  
\beta(t) =  \beta_{0}\mathcal{H}(t_{1} - t) + \sum_{i = 1}^{n-1}\beta_{i}\mathcal{H}(t_{i+1}-t)\mathcal{H}(t-t_{i}) + \beta_{n}\mathcal{H}(t-t_{n}),    
\end{equation}

# %% [markdown]
#### Defining the Steap Function

# %% [code]
 
k=50
def H(t):
    h = 1.0/(1.0+ np.exp(-2.0*k*t))
    return h


# %% [markdown]
#### Defining the $\beta(t)$ with one changing moment

# %% [code]
def beta(t,t1,b,b1):
    beta = b*H(t1-t) + b1*H(t-t1) 
    return beta

# %% [code]
import numpy as np
t = np.arange(1, 100, 1)

t1 = 15
b = 0.5 
b1 = 0.3

beta_function = beta(t, t1, b, b1)

beta_function

# %% [markdown]
#### Defining the $\beta(t)$ with two changing moment

# %% [code]
#Defining the Beta(t)
def beta(t,t1,t2,b,b1,b2):
    beta = b*H(t1-t) + b1*H(t2-t)*H(t-t1) + b2*H(t-t2) 
    return beta

#t3 = 300

# %% [code]
import numpy as np
t = np.arange(1, 100, 1)

t1 = 15
t2 =  60 
b = 0.5 
b1 = 0.3
b2 = 0.4

beta_function = beta(t, t1, t2, b, b1, b2)

beta_function

# %% [markdown]
#### Defining the $\beta(t)$ with three changing moment

# %% [code]
#Defining the Beta(t)
def beta(t,t1,t2,t3,b,b1,b2,b3):
    beta = b*H(t1-t) + b1*H(t2-t)*H(t-t1) + b2*H(t3-t)*H(t-t2) + b3*H(t-t3)  
    return beta

#t3 = 300

# %% [code]
t = np.arange(1, 150, 1)
t1 = 15
t2 =  60 
t3 = 100
b = 0.5 
b1 = 0.3
b2 = 0.4
b3 = 0.2

beta_function = beta(t, t1, t2,t3, b, b1, b2,b3)

beta_function

# %% [code]
```
